# Optimal vaccination with time-varying based on immunity barrier in Hunan Province, China

**DOI:** 10.1101/2022.03.02.22271610

**Authors:** Xiaohao Guo, Ziyan Liu, Shiting Yang, Zeyu Zhao, Yichao Guo, Guzainuer Abudurusuli, Jia Rui, Yao Wang, Shanlu Zhao, Ge Zeng, Shixiong Hu, Kaiwei Luo, Tianmu Chen

## Abstract

The current outbreak of novel coronavirus disease 2019 (COVID-19) is already causing a serious disease burden worldwide, this paper analyzed data of a delta variant Covid-19 outbreak in Hunan, China, and proposed an optimal dose-wise dynamical vaccinating process based on local contact pattern and vaccine coverage that minimize the accumulative cases in a certain future time interval. The optimized result requires an immediate vaccination to that none vaccinated at age group 30 to 39, which is coherent to the prevailing strategies. The dose-wise optimal vaccinating process can be directive for countries or regions where vaccines are not abundant. We recommend that vaccination should be further intensified to increase the coverage of booster shots, thus effectively reducing the spread of COVID-19.

## Introduction

At the end of 2019, the novel coronavirus disease 2019 (COVID-19) caused by severe acute respiratory syndrome coronavirus 2 (SARS-CoV-2) first broke out in Wuhan, China, and then swept across the world at an extremely rapid pace. As of November 17, 2021, a total of 253,640,693 confirmed cases globally with 5,104,899 deaths had been reported by WHO[1]. Although several targeted interventions have been implemented globally to contain the spread of the new crown epidemic, the situation remains challenging as the delta variant strain becomes dominant. To date, a total of 7,084,922,999 doses of vaccine have been administered worldwide, and a total of 2.338493 billion doses of new-coronavirus vaccine have been reported in China[2], representing 78% of the country’s total population[3]. A safe and effective new-coronavirus vaccine is essential to end the outbreak of novel coronavirus, and widespread vaccination has led to a milestone victory in the outbreak. However, the emergence of the Delta variant poses a challenge to the effectiveness of the new-coronavirus vaccine. The US Centers for Disease Control and Prevention began recommending that all people who receive the Johnson & Johnson vaccine receive a booster at least 2 months in advance[4]. Regular vaccinations help to maintain antibody levels in the body and a booster shot of the new-coronavirus vaccine may be the general trend.

SIR models have been constructed based on COVID-19 contact tracing information in Hunan Province to investigate the impact of age-mixing patterns on epidemic dynamics[5]; Zhang et al. collected contact survey data from four regions of China (Wuhan, Shanghai, Shenzhen and Changsha) during a pandemic and used the models to assess the impact of changes in age-stratified contact patterns on SARS-CoV-2 transmission[6]; and The mathematical model of the age structure of SARS-CoV-2 transmission in the study by Laura Matrajt et al. identified the optimal vaccine distribution strategy[7]. The age models in most previous studies have been simulated on exposure data, but the model in this study was simulated on close contact data. There are two advantages in evaluating close contact data. The first is that the data are more reflective of the true exposure pattern of the infected person; the second is that such data of close contacts are more readily available.

Vaccines are a powerful tool in the current fight against the epidemic, but with limited supplies, it is important to consider who should be prioritized for the new-coronavirus vaccine and what vaccine distribution scheme should be implemented to maximize health and economic benefits. WHO believes that those who could benefit from safe and effective COVID-19 vaccines should have access as quickly as possible, starting with those at the highest risk of serious disease or death[8]. As age is one of the risk factors for serious COVID-19 infections and death[9], the elderly population is the most frequently cited priority target group[10], but studies have also suggested that there is a strong moral case for giving children priority for vaccination to protect the elderly[11]. Depending on the objectives, several studies have suggested that priority should be given to vaccinating the elderly population against the new coronavirus to reduce the number of seriously ill patients and reduce mortality, and to the young population to reduce the spread of the new coronavirus[12, 13]. According to the effectiveness and availability of the vaccine, it has been suggested that in cases of low vaccine effectiveness and low vaccine availability, it is preferable to allocate the vaccine to the higher age groups first; in cases of high vaccine effectiveness and high vaccine coverage, it is preferable to allocate the vaccine to the lower age[7]. As the dynamics of the novel coronavirus pneumonia epidemic and the status of vaccine supply continue to change, we should adjust our immunization strategy promptly and identify priority target groups for vaccination.

To better understand the impact of age-specific exposure patterns on the spread of the New Coronavirus outbreak and to prioritize age-specific immunization, this study analyzed local exposure patterns and vaccine effects using real-life event data from Hunan Province and simulated the transmission characteristics of age groups based on exposure patterns; future real-time vaccination scenarios were further simulated on existing vaccination patterns.

## Methods

We first evaluate the vaccine coverage in population by inspecting the vaccination state (none, un-fully vaccinated, fully vaccinated, booster vaccinated) of close contacts in contact data.

In the outbreak in Hunan, the total attack rates (TAR) of people in different age groups and people with 4 vaccination states finished are very interesting parameters. To handle the uncertainty of the collected data, we use bootstrap to construct the empirical distribution of all these TARs. We performed the bootstrap 10000 times on the contact data, with each bootstrapped data set containing equally 10000 contacts. Each of those bootstrapped data sets produces an 8 × 4 matrix of TAR in age groups (in rows) and vaccine groups (in columns). Based on those resampled TARs, we depict the distribution of TAR in age groups and vaccine groups in box-plot respectively.

A bipartite graph model is adopted to describe the contact pattern (contact matrix) between age groups (more details in appendices). The population size in each group is considered as a modification). We first construct the contact data matrix by rearranging all the case-contact data pairs. Then, based on this model, three methods (least square, weighted least square, and maximum likelihood) are developed for estimating the contact matrix.

To evaluate the effectiveness of vaccination, a grouped *VEFIAR* model (Figure 1) is developed with different age groups and vaccinated groups. The vaccine efficacy is considered as a reducing coefficient of susceptibility and is set to be 0, 0.40, 0.56, 0.802 for 0, 1, 2, 3 vaccination states. The definition of compartments and parameters is given in appendices.

**Figure 1:**
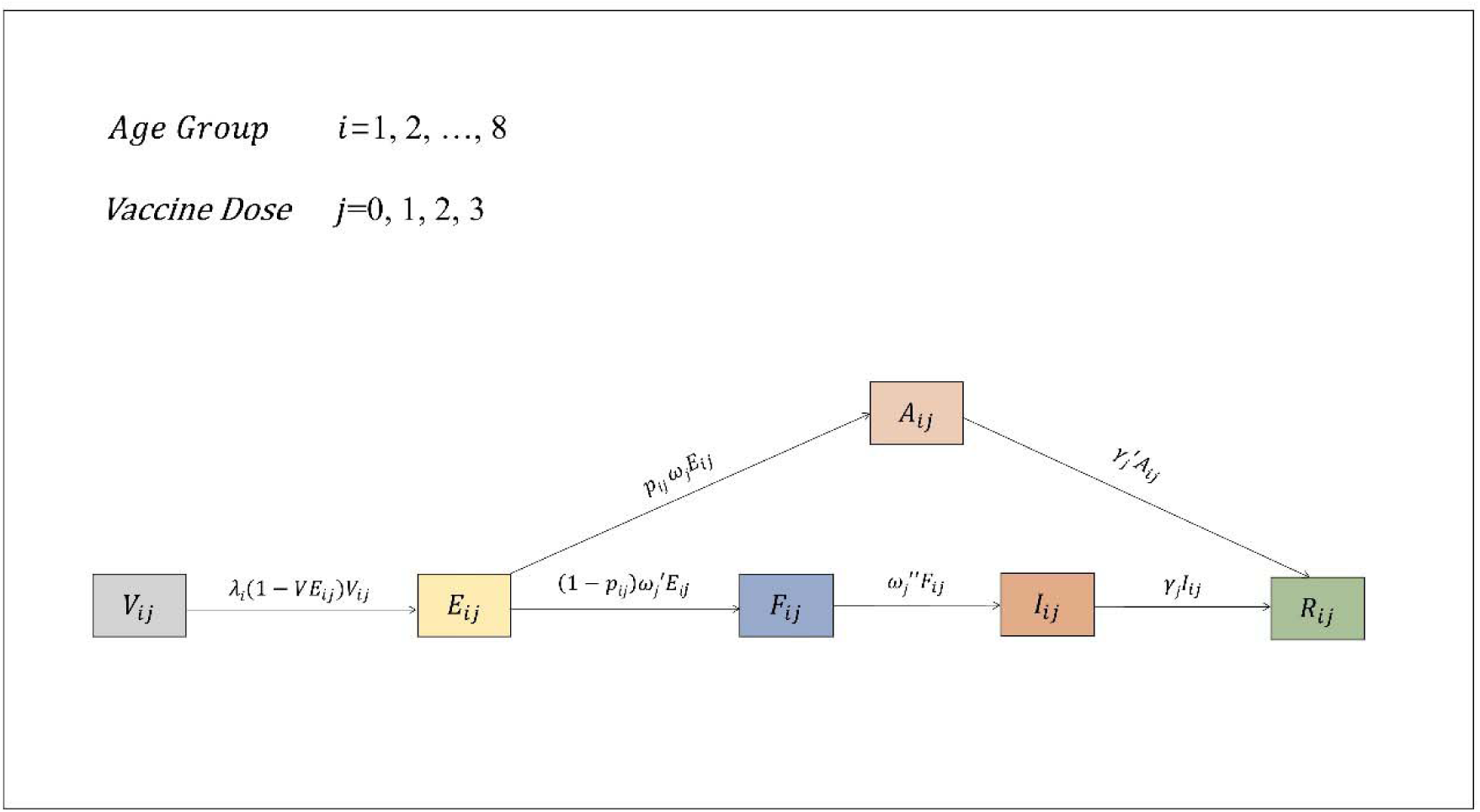
Flowchart of The Multi-Group VEFIAR Model.

In Figure 1, *λ*_*i*_ is the force of infection defined as:

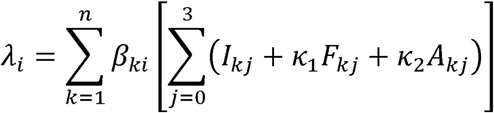

To introduce the impact of contact patterns in the grouped *VEFIAR* model, the transmission rate coefficients *β*_*ij*_ are formularized as the products of the number of daily average contacts, the probability of infection after single one-time contact, and the susceptibility of people in the *j*-th group, that is, *β*_*ij*_ *N*_*i*_ = *c*_*ji*_ *q σ*_*j*_. Where *c*_*ji*_ is the *ji*-th entry of contact matrix; *q* is the probability of infection after a single one-time contact; *σ*_*j*_ is the susceptibility of group *j*.

The reproduction number *R*_0_ computed by next-generation method is a function of all *β*_*ij*_ and other parameters and the disease-free equilibrium. With the assumption of decomposition of *β*_*ij*_ and the assumption of equal susceptibility in age groups, *R*_0_ is indeed a function of contact matrix *C*, probability of contact infection *q*, and all other parameters and the disease-free equilibrium. In our simulation, the probability of infection *q* is solved from the simulated *R*_0_, and all *β*_*ij*_ are recovered from the contact matrix and *q*.

The grouped *VEFIAR* model is tested by simulating with different *R*_0_. The contact pattern involved for recovering all *β*_*ij*_ from *R*_0_ is set to be the contact matrix estimated via maximum likelihood estimation (MLE), and the first case is set to be in age group 50 to 59, with 3 doses finished (as it was in this outbreak). For each simulated *R*_0_, the daily incidence rates are shown for each vaccine group, and the total attack rates in 100-days are shown for each group.

Optimizing the dynamic vaccinating process is indeed an optimal control problem that can be transformed into a nonlinear programming problem (NLP) by discretizing the control, and solved by a variety of numerical methods. In the real world, the vaccinating process is not always parallel with abundant vaccines, rather than a small amount of step-wise vaccination. For this consideration, we use the greedy algorithm to find the optimal vaccinating process under the current vaccine coverage. The object function (the function to be minimized) is constructed as the accumulative number of cases (summation of integration of all *F*_*ij*_ and *A*_*ij*_) during the 100-days simulation (that is, if we assigned this dose to such group, then how many accumulative cases are expected to reduce within the next 100-days).

The directional derivative is introduced as an index for the effectiveness of the current dose vaccinated in different groups. The optimal step-wise vaccinating strategy is then obtained by minimizing the directional derivative for each dose. We visualized the optimal vaccine process by showing the population curves of each group (age groups and vaccine groups).

## Result

The bar-plot (Figure 2) of vaccine coverage (people vaccinated at least 1 dose, 2 doses, or 3 doses) showed a single peak function of ages, with the youngest and the oldest has the minimal vaccine coverage, and those in middle ages (40 to 59 years old) has the highest vaccine coverage. Children of 0 to 9 years old are almost non-vaccinated (only 0.9% coverage of least 1 dose), and for other groups, there are still significant proportions of non-vaccinated people (without even 1 dose).

**Figure 2:**
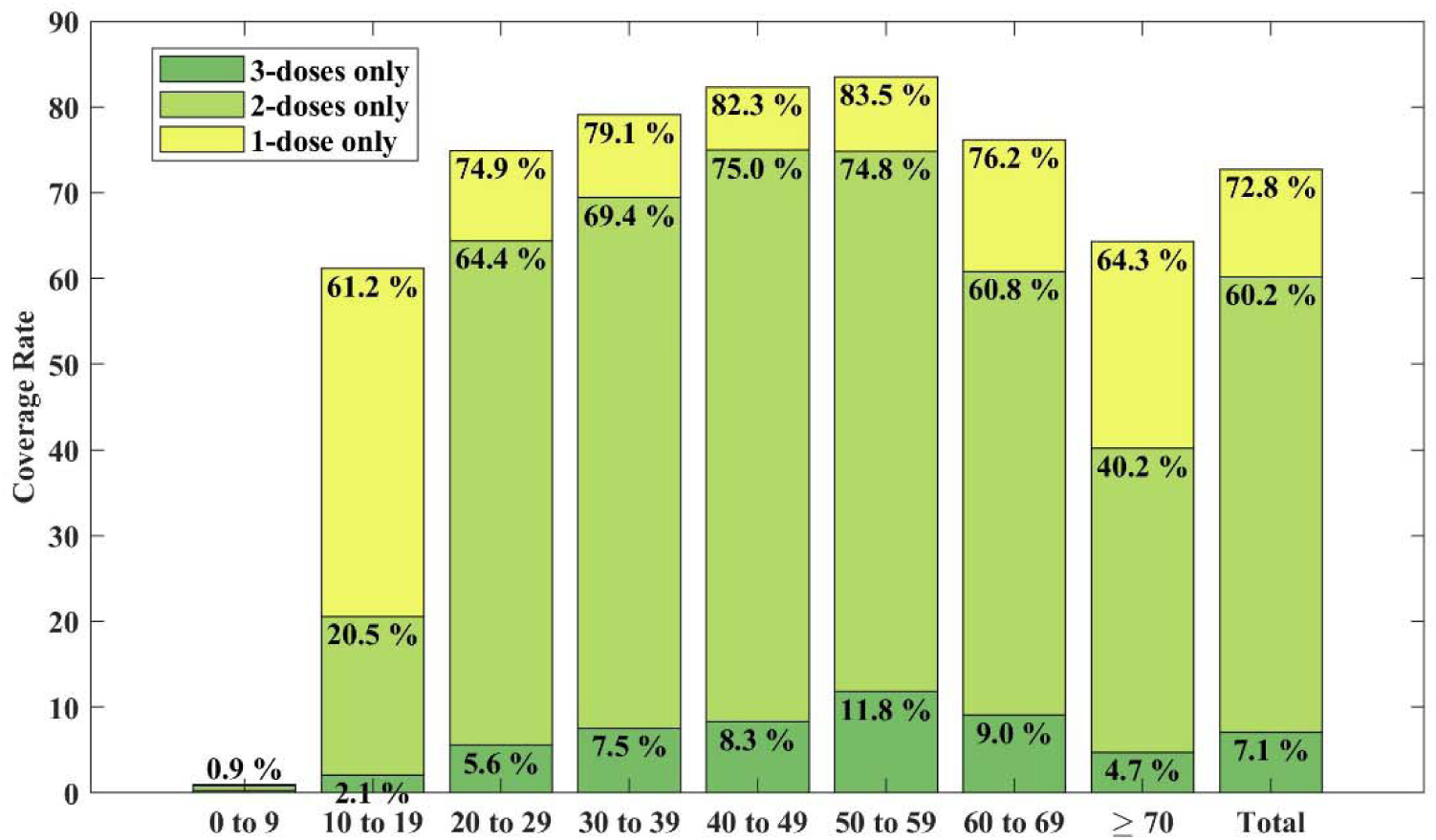
The Current Vaccine Coverage, based on vaccination data of the 17821 close contacts in this outbreak.

The boxplot in Figure 3 shows the different distribution of bootstrapped TAR in age groups and vaccine groups. TAR in age group 0 to 9 showed a positive-skewed distribution, mainly due to the lack of cases in this group. Thus, we may consider the TAR in age groups 0 to 9 as the lowest. Besides this skewed group, since the notches in the boxplot do not overlap, we are sufficient with 95% confidence that the true medians of TAR in age group greater than 70 are lowest, and followed by age group 20 to 29. The medians of TARs in all age groups are around 10%, which means the TAR in age groups does not change much.

**Figure 3:**
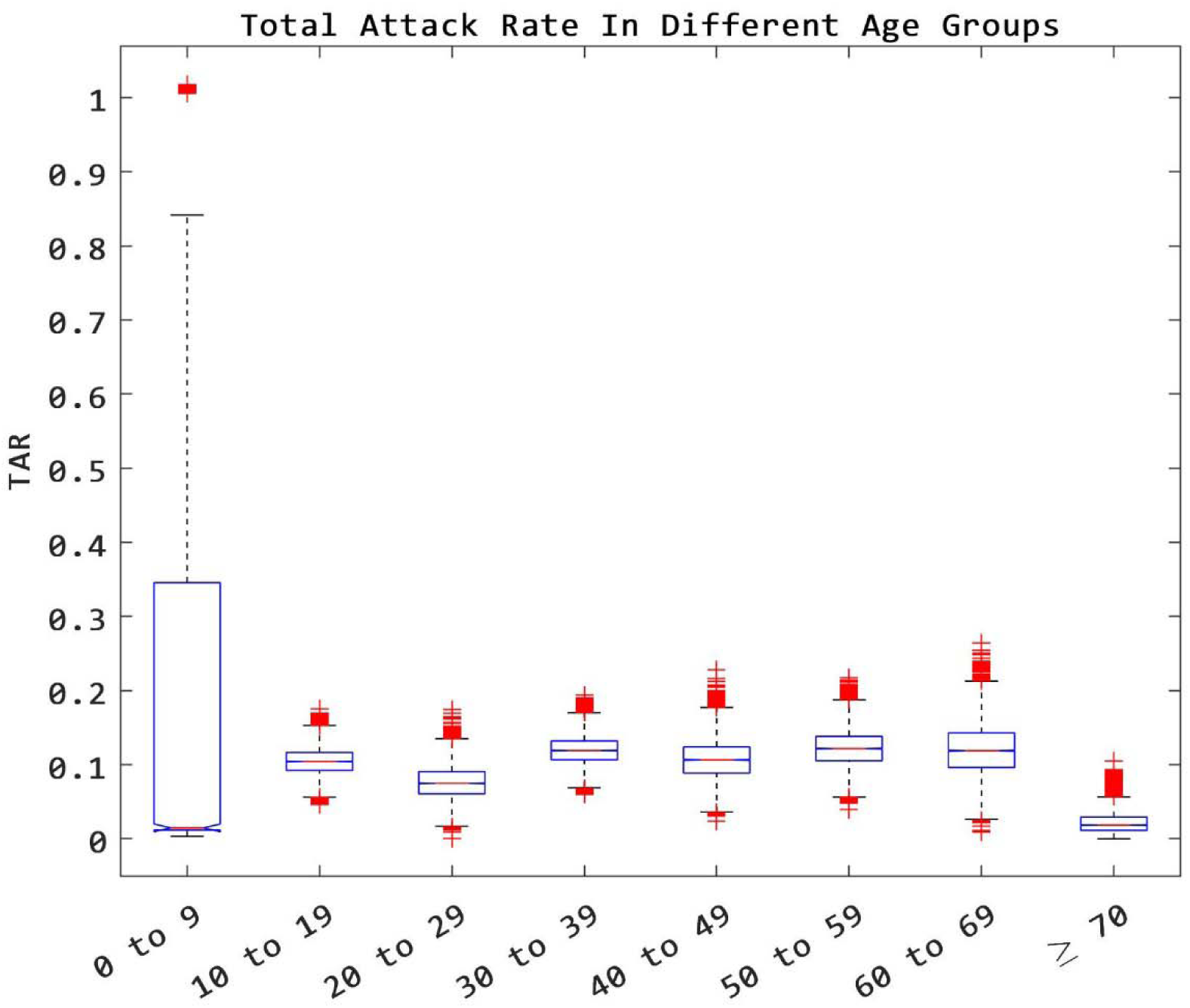
Total Attack Rate In Different Age Groups. The method of bootstrap is used to illustrate the distribution of TAR in specific groups. 10000 times of bootstrap of the close contact data are performed, with each bootstrap sample size equal 10000. Each bootstrapped data set produces a sample matrix of TAR (of each age/dose group)

For the vaccine groups (Figure 4), similarly by inspecting the notches, we conclude with 95% confidence that the median of TAR decrease by doses.

**Figure 4:**
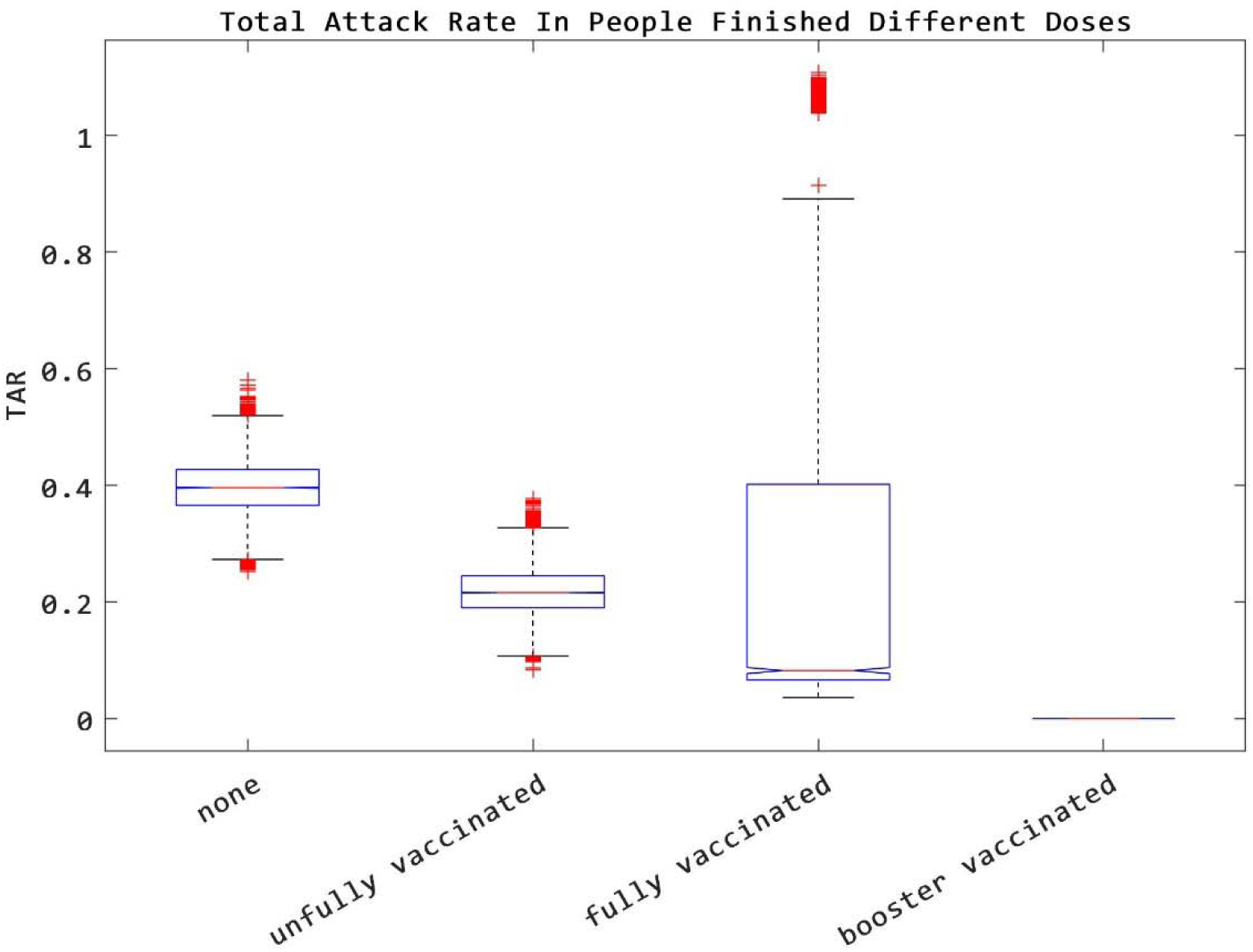
Total Attack Rate In People Finished Different Doses. The method of bootstrap is used to illustrate the distribution of TAR in specific groups. 10000 times of bootstrap of the close contact data is performed, with bootstrap sample size equals 10000. Each bootstrapped data set produces a sample matrix of TAR (of each age/dose group)

The contact matrix (Figure 5) showed a small adjustment compared with the contact data matrix. Both of the matrices showed a concentration in diagonal entries and middle entries, which means people in the same age group are more likely to make contact, and people in middle ages are more likely to contact with others (middle ages are more likely to have children and parents to feed). Our matrices are coherent with the result of the contact matrix in Wuhan and Shanghai, two cities in China, given by.

**Figure 5:**
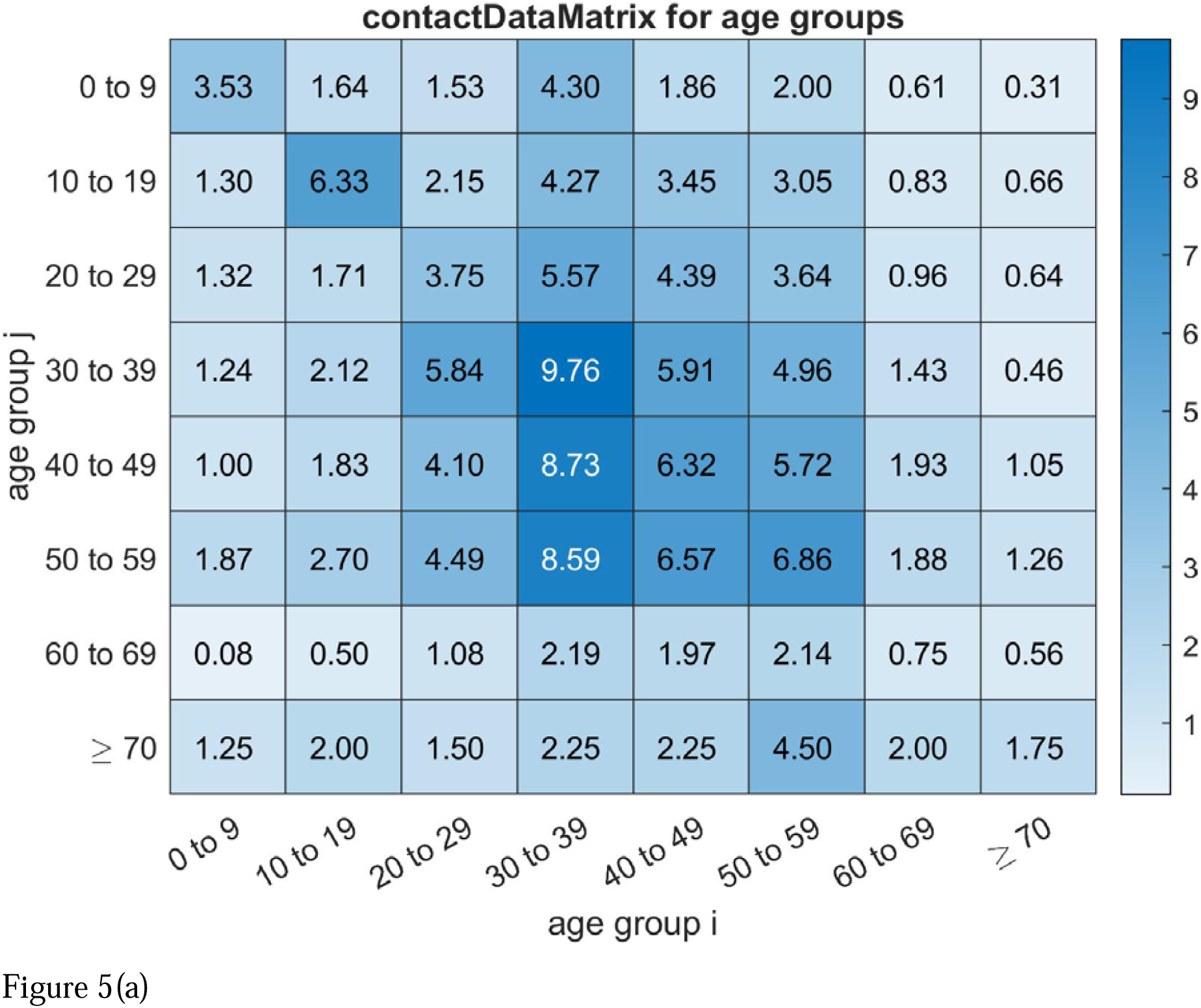
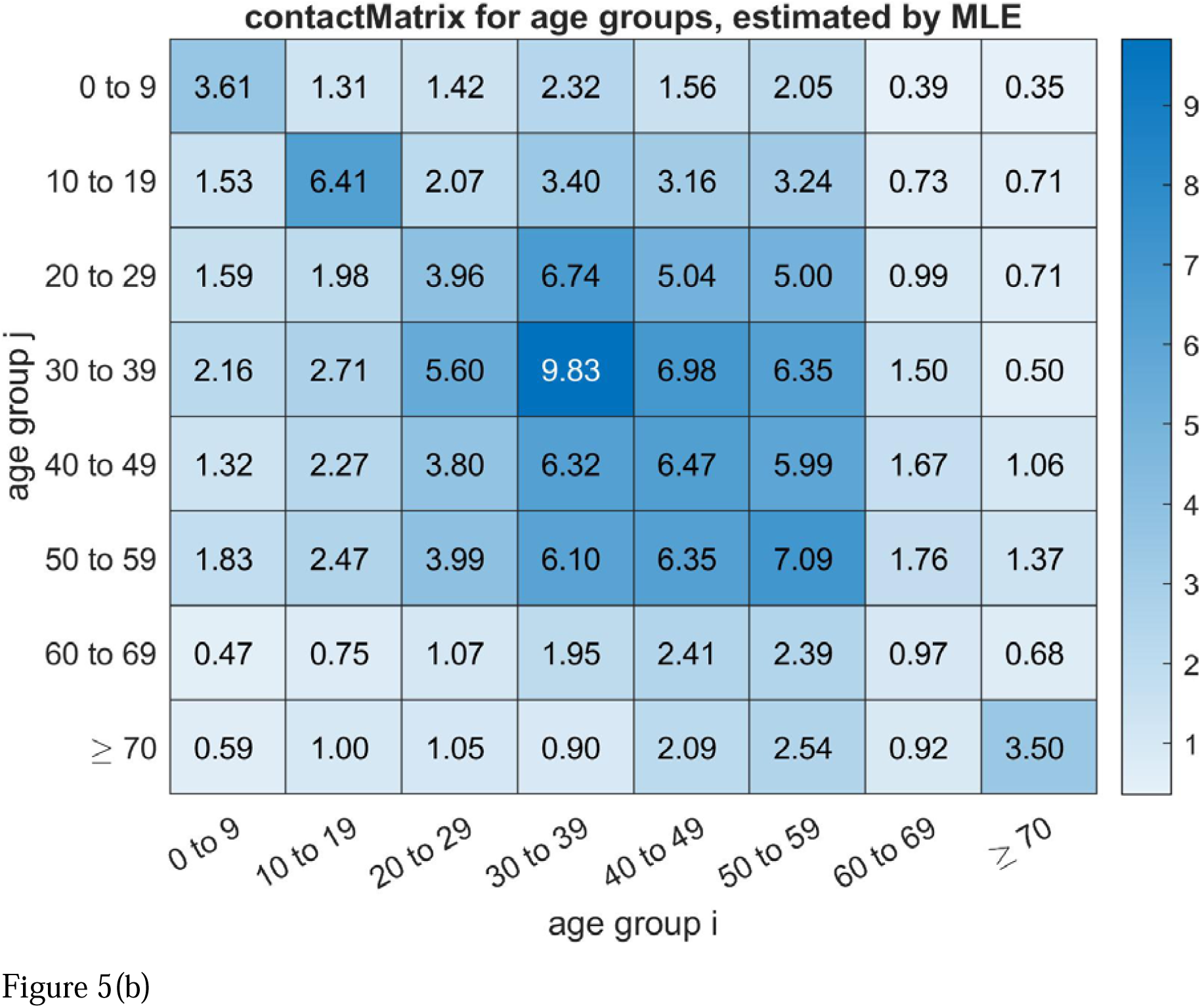
Contact Data Matrix and Contact Matrix. (a) Contact Data Matrix filled with contact data. (b) Contact Matrix is estimated by the method of maximum likelihood estimation (based on a bipartite graph model).

The simulation under current vaccine coverage and VE with initial case in 30 to 39 group (Figure 6) showed quantitatively that the more doses finished, the lesser the daily incidence is. For *R*_0_ < 1, the disease distinct fast. For *R*_0_ =1, due to the existence of rounding error, we can see that the disease persists within an acceptable error. For *R*_0_ < 1, the peak value of daily incidence increased, and the duration of the epidemic shorten, with the increase of *R*_0_.

**Figure 6:**
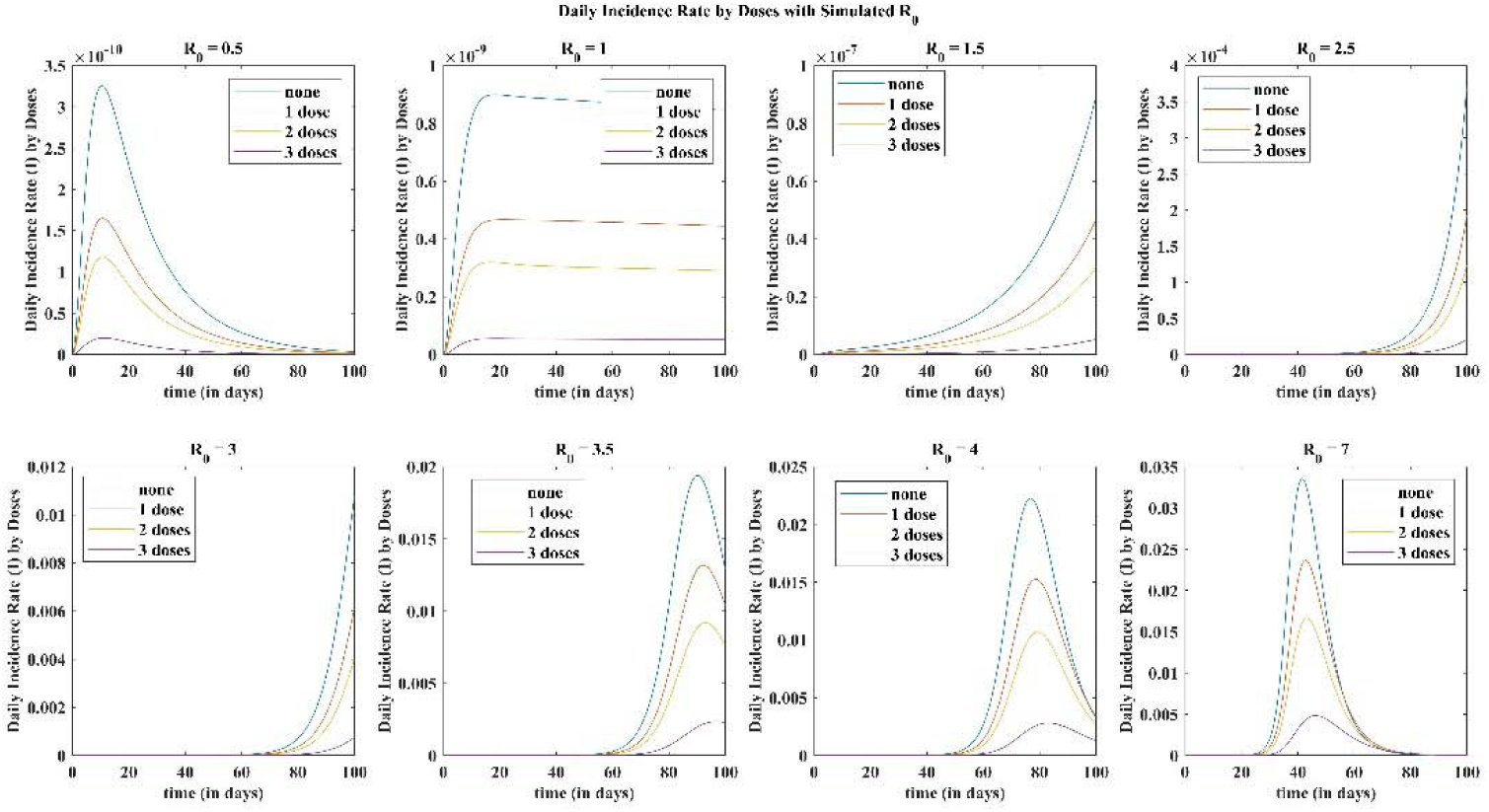
Daily Incidence Rate Given by The VEFIAR Model with Simulated ***R***_**0**_.

The total attack rate in each (age, vaccine) group in the 100-days simulation (Figure 7) told us how risky each group is (under the current vaccine coverage and contact pattern in the area). It can be seen from the following Figure that the risk decreases with doses, and concentrated on those in the middle ages. The TAR of the fully vaccinated group is much lower than those un-fully vaccinated. The result of the booster-vaccinated group is unreliable due to its small sample size.

**Figure 7:**
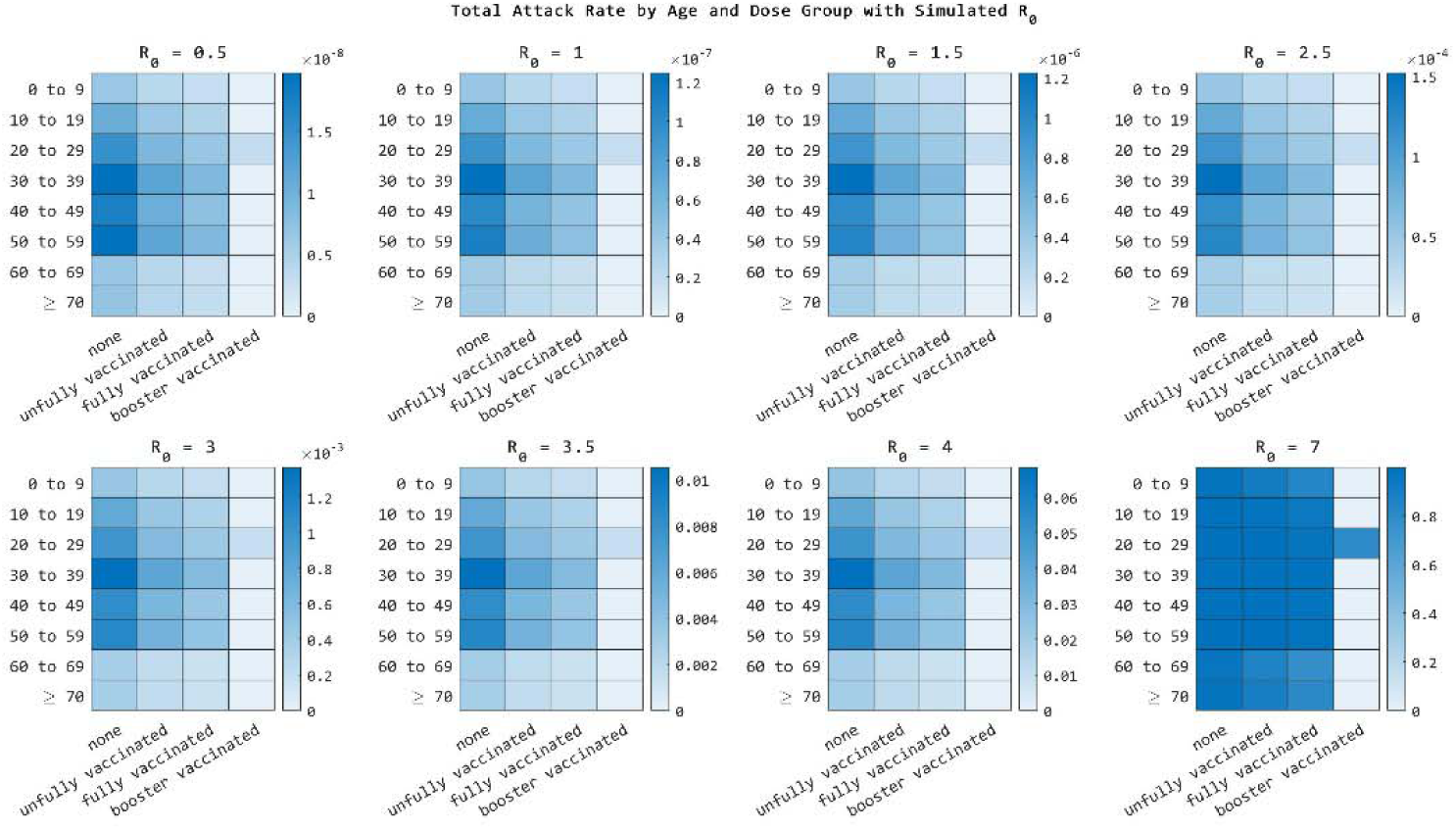
Total Attack Rate in A 100-Days Simulation of Current Vaccine Coverage and Contact Pattern with Different Simulated ***R***_**0**._

The optimal vaccinating process in Figure 8 is obtained by minimizing the directional derivative in each vaccinating step. This Figure gives a hint of how shall we distributing the not-abundant vaccines in groups and which group should be first to vaccinate.

**Figure 8:**
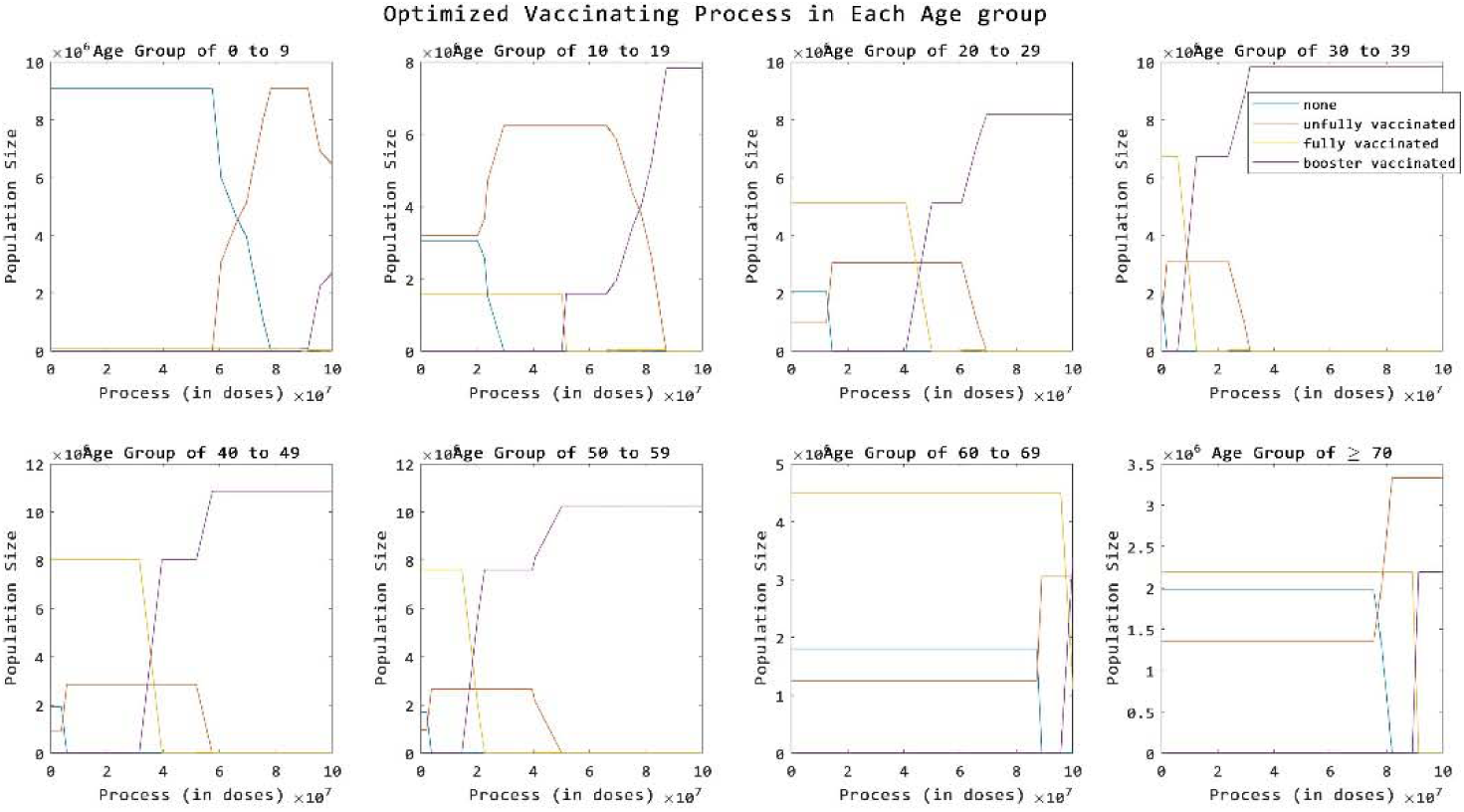
Optimal Vaccinating Process Under Current Contact Pattern and Vaccine Coverage.

We can see that the optimal process requires an immediate vaccination to those un-vaccinated in age group 30 to 39 after the high-risk occupations been well vaccinated (In the upper right sub-Figure, the population size of un-vaccinated group increase with the first dose, which means this group should be vaccinated first), in the sense of minimal accumulative cases with 100-days.

If despite the age, and focus only on the vaccine groups, we can found that the unvaccinated group should be vaccinated first, since, in all 8 sub-figures, the un-vaccinated are always vaccinated at the first.

Figure 9 showed how the accumulative cases decrease with the optimal vaccinating process (the effectiveness of vaccination), under different simulated *R*_0_.

**Figure 9:**
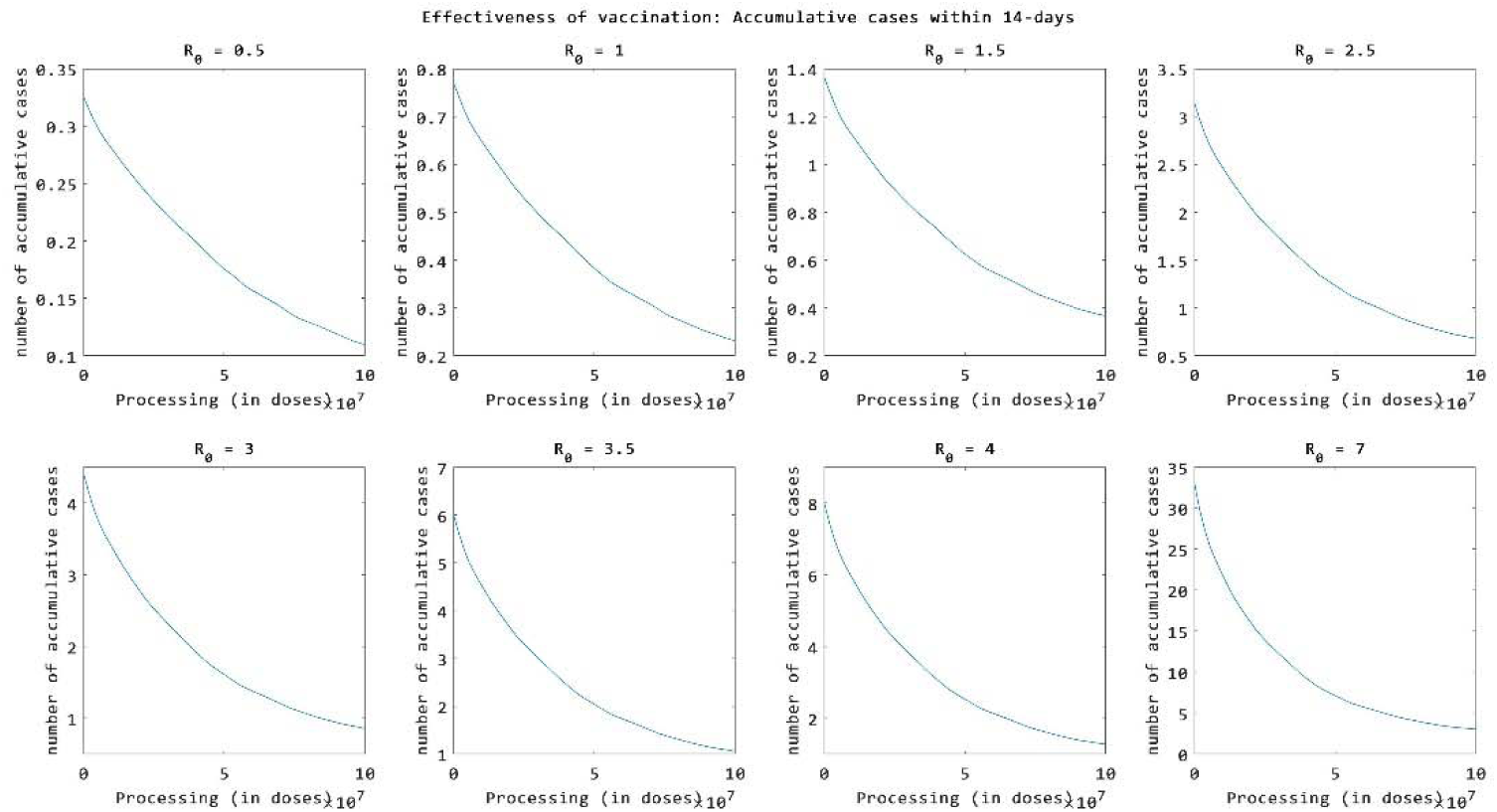
Reduction of Accumulative Cases within 14-days with the Vaccinating Processing

We found that all the curves are concave upward, and the slope decreases all the time. That is, the effectiveness of a single dose will be reduced if the vaccination is nearly complete. This may indicate the relatively average distribution in each group.

## Discussion

**Figure 1: The Current Vaccine Coverage, based on vaccination data of the 17821 close contacts in this outbreak**.

### Validity of close-contact matrix

Usually, contact degrees in the different age groups from a survey are always used to simulate the age-specific transmission in SARS-CoV-2[14]. However, the dataset of close-contact surveyed from reported cases was firstly used to estimate the transmission pattern in different age groups. We found that our close-contact matrix was consistent with an age-specific contact surveyed by a study[6], suggesting it is feasible to simulate some scenarios (like sex, age, school, workplace, etc.) by the close-contact matrix. In the study, we further found that the high contact frequency occurred in the middle age group, also leading it a high proportion of reported cases. Therefore, it could adopt the data of close contact to simulate the transmission in different age groups.

### Real-word vaccine effectiveness

As of November 2021, China has been fully vaccinated by 50% of people[15]. We found a relatively high vaccine coverage with 27.24% of none vaccinated, 13.54% of vaccinated but un-fully, 59.21% of fully vaccinated, 0.0006942% of booster vaccinated in Hunan Province, leading the local area an immune barrier to control the outbreak of Delta variant. The vaccine effectiveness had been increased with the enhancing of vaccination doses, which is also similar reported in some studies[16]. Although there was a significant difference in the relative risk of infection in different age groups, the high age group is only 1.03 times that in the low age group, which exists a low effect in our model.

### Model validity

The age-specific SEFIAR model can effectively simulate the transmission pattern in different age groups, which was similar to our previous studies[17]. Furthermore, the model further considered the vaccine coverage and immunity of real-world data from Hunan Province, leading a representative with our simulation of the age-specific model.

### Age-specific transmission

We calculated most cases aged 30 to 39 years old, which was consistent with a 27.42% proportion reported cases mainly reported in those above groups. The contact degree (mean: 9.831 people per day) in those age groups was higher than the mean value of the total population (mean: 2.797 people per day), remaining a relatively high transmission of the virus. This is similar to a study indicating that transmission mainly occurred in middle and old people[17].

### Real-word vaccine effectiveness

The total population would early establish the immunity after xx% with vaccination, xx with full vaccination, and only xx with strengthening. However, the transmission of the Delta variant could not be fully interrupted after vaccination in Hunan Province. R_0_ of the Delta variant was reported from 3.2-8.0, 1.5 times than the adolescent virus according to the previous study[18]. Our finding also suggested the new variant existed breakthrough of infection. The vaccine efficacy was higher than 60% after full vaccination according to reports[19, 20]. The vaccine had produced some effectiveness in Hunan Province according to real-world data analysis. In the study, we found a low difference in relative risk of infection in various age groups after vaccination, therefore, we did not consider it equal in our dynamics model.

### Effectiveness of real-time vaccination

According to the current immunity barrier and contact pattern, we found that the none vaccinated group of age 30 to 39 should be in the next vaccination plan. Our finding suggested that. Although a study had been proposed real-time vaccination strategy can effectively reduce morbidity and mortality[21], they could not sufficiently consider the real-world parameters such as immunity barrier and contact pattern.

## Limitation

We limited the close-contact data surveyed by reported cases that could not fully represent the contact pattern of Hunan Province. The reliability of data needs to be further tested contact patterns from the real world. Vaccine coverage of Hunan Province had been gradually increased, suggesting we should simulate most scenarios about the current level of the immune barrier.

## Data Availability

All data produced in the present study are available upon reasonable request to the authors

