## Appendices for "Optimal vaccination with time-varying based on immunity barrier in Hunan Province, China": Appendices20220301.pdf

### Appendix 1

February 28, 2022

#### 1 Contact Matrices

##### 1.1 Construction of the Contact Matrix

**Definition:**(Contact Matrix)

The contact matrix $C$ is a square matrix with its $ij$-th entry $c_{ij}$ denotes the average number of daily contacts in group $j$ produced by a individual in group $i$.

##### 1.2 Graph Model of the Contact Matrix

To illustrate the relation between $c_{ij}$ and $c_{ji}$, we consider a bipartite graph of group $i$ and $j$, by omitting their inner edges and edges connect other groups. In group $i$, there are $N_i$ individuals denoted by vertices $V_1, V_2, \cdots, V_{N_i}$; and for group $j$, the $N_j$ individuals are denoted by vertices $W_1, W_2, \cdots, W_{N_j}$. Each contact data pair is denoted by an edge connect group $i$ and group $j$.

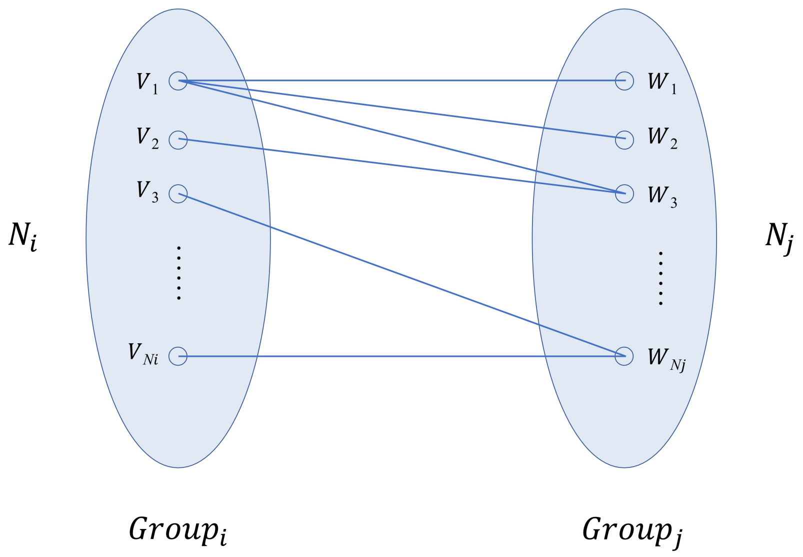


(a)

Fig. 1: Bipartite Graph Model

By the previous definition of contact matrix, $c_{ij}$ is the average degree of vertex in group $i$ (in this bipartite graph):

$$c_{ij} = d_i \stackrel{def}{=} \frac{1}{N_i} \sum_{k=1}^{N_i} deg(V_k) \tag{1}$$

$$c_{ji} = d_j \stackrel{def}{=} \frac{1}{N_j} \sum_{k=1}^{N_j} deg(W_k) \tag{2}$$

where $deg()$ is the the degree of a vertex, i.e. the number of edges attached to the vertex.

In order to establish the relation between $c_{ij}$ and $c_{ji}$, it is naturally to consider the conservative quantity in this system. It is found that for these two groups of vertices, the total number of edges between them is constant, no matter how the connective pattern changes. Therefore:

$$N_i d_i = N_j d_j \tag{3}$$

That is,

$$d_j = \frac{N_i}{N_j} d_i \tag{4}$$

i.e.

$$c_{ji} = \frac{N_i}{N_j} c_{ij} \tag{5}$$

#### 1.3 Construction of the Contact Data Matrix

In construction of contact matrix $C$, the contact data matrix $A$ is firstly filled using (case, contact) data pairs——after initialized $A = 0$, we locate the age group for each case, then add its close contacts to the corresponding row of A. After all data pairs are filled, we divide each row of A by the total number of cases belongs to this group (of averagely one individual), and then divide a 4 day period of the close contact survey.

Matrix $A$ reflects the contact pattern of the (caseAge, contactAge) pairs. However, it may has significant noise due small sample size of incident cases. Therefore, a correction for $A$ is needed to approximate the real contact matrix $C$.

The following are data matrix $A$ known, and the contact matrix $C$ with $\frac{n(n+1)}{2}$ parameters to be fitted:

$$A = \begin{array}{|c|c|c|c|c|c|c|} \hline a_{11} & a_{12} & a_{13} & \cdots & \cdots & \cdots & a_{1n} \\ \hline a_{21} & a_{22} & a_{23} & \cdots & \cdots & \cdots & a_{2n} \\ \hline a_{31} & a_{32} & a_{33} & \cdots & \cdots & \cdots & a_{3n} \\ \hline \vdots & \vdots & \vdots & \ddots & & & \vdots \\ \hline \vdots & \vdots & \vdots & & \cdots & & \vdots \\ \hline \vdots & \vdots & \vdots & & & \ddots & \vdots \\ \hline a_{n1} & a_{12} & a_{13} & \cdots & \cdots & \cdots & a_{nn} \\ \hline \end{array} \qquad C = \begin{array}{|c|c|c|c|c|c|c|} \hline c_{11} & c_{12} & c_{13} & \cdots & \cdots & \cdots & c_{1n} \\ \hline \frac{N_1}{N_2}c_{12} & c_{22} & c_{23} & \cdots & \cdots & \cdots & c_{2n} \\ \hline \frac{N_1}{N_3}c_{13} & \frac{N_2}{N_3}c_{23} & c_{33} & \cdots & \cdots & \cdots & c_{3n} \\ \hline \vdots & \vdots & \vdots & \ddots & & & \vdots \\ \hline \vdots & \vdots & \vdots & & \cdots & & \vdots \\ \hline \vdots & \vdots & \vdots & & & \ddots & \vdots \\ \hline \frac{N_1}{N_n}c_{1n} & \frac{N_2}{N_n}c_{2n} & \frac{N_3}{N_n}c_{3n} & \cdots & \cdots & \cdots & c_{nn} \\ \hline \end{array}$$

#### 1.4 Estimations of Contact Matrix

Here we use one of the three following methods to estimate the contact matrix $C$ from contact data matrix $A$.

- least square estimation
- weighted least square estimation
- maximum likelihood estimation

##### 1.4.1 Least Square Estimation of the Contact Matrix

The simplest estimation of those $c_{ij}, \ \ i \leq j, \ \ i, j = 1, 2, \cdots, n$ is the least square fitting described by the following optimization problem:

$$\underset{\forall c_{ij},\ i \leq j}{minimize} \|A - C\|_F \tag{6}$$

where $\|\cdot\|_F$ denotes the Frobenius norm of matrix.

The optimization problem 6 can be reduced to sub-problems to divide and conquer:

$$\hat{c}_{ij} = \underset{c_{ij}}{argmin} \;\; (c_{ij} - a_{ij})^2 + [(\frac{N_i}{N_j})c_{ij} - a_{ji}]^2 \tag{7}$$

$$= \underset{c_{ij}}{argmin} \;\; (1 + \frac{N_i^2}{N_j^2})c_{ij}^2 - (2a_{ij} + 2\frac{N_i}{N_j}a_{ji})c_{ij} + (a_{ij}^2 + a_{ji}^2) \tag{8}$$

$$= \frac{a_{ij} + \frac{N_i}{N_j}a_{ji}}{1 + \frac{N_i^2}{N_j^2}}, \quad \forall ij, \; i < j \tag{9}$$

$$\tag{10}$$

$$\hat{c}_{ji} = \frac{N_i}{N_j}\hat{c}_{ij}, \quad \forall ij, \; i < j \tag{11}$$

$$\tag{12}$$

$$\hat{c}_{ii} = \underset{c_{ii}}{argmin} \;\; (c_{ii} - a_{ii})^2 \tag{13}$$

$$= a_{ii}, \quad i = 1, 2, \cdots, n \tag{14}$$

We can see that the contact matrix $C$ and its estimation are both symmetric if the population of groups are equal, i.e. $N_i = N_j$.

##### 1.4.2 Weighted Least Square Estimation of the Contact Matrix

The unbalanced sample size of cases in each groups requires different importance in errors of each entry of $C$. Therefore, we use sample size (number of cases in each groups) to weight the error, which leads the following adjusted optimization problems:

$$\hat{c}_{ij} = \underset{c_{ij}}{argmin} \ \ n_i(c_{ij} - a_{ij})^2 + n_j[(\frac{N_i}{N_j})c_{ij} - a_{ji}]^2 \tag{15}$$

$$= \underset{c_{ij}}{argmin} \ \ (n_i + \frac{N_i^2}{N_j^2}n_j)c_{ij}^2 - (2n_ia_{ij} + 2n_j\frac{N_i}{N_j}a_{ji})c_{ij} + (n_ia_{ij}^2 + n_ja_{ji}^2) \tag{16}$$

$$= \frac{n_ia_{ij} + n_j\frac{N_i}{N_j}a_{ji}}{n_i^2 + \frac{N_i^2}{N_j^2}n_j^2} \tag{17}$$

$$\tag{18}$$

$$\hat{c}_{ji} = \frac{N_i}{N_j}\hat{c}_{ij} \tag{19}$$

$$\tag{20}$$

$$\hat{c}_{ii} = \underset{c_{ii}}{argmin} \ \ (c_{ii} - a_{ii})^2 \tag{21}$$

$$= a_{ii} \tag{22}$$

where $n_i$ and $n_j$ denote the number of cases in group $i$ and $j$.

##### 1.4.3 Maximum Likelihood Estimation of the Contact Matrix

Since the Poisson distribution is frequently used to describe phenomena that have few positive outcomes over many repeating trials, we assume that:

$$c_{ij} \sim Poisson(\lambda) \tag{23}$$

$$c_{ji} \sim Poisson(\frac{N_i}{N_j}\lambda) \tag{24}$$

Let $b_{ij}(k)$ denotes the number of contact in group $j$ produced by $k$-th observed case in group $i$ .

Then the data set for training $c_{ij}$ can be represented as:

$$b_{ij}(k), \ \ k = 1, 2, \cdots, K_i \tag{25}$$

$$b_{ji}(k), \ \ k = 1, 2, \cdots, K_j \tag{26}$$

where $K_i$ is the total number of cases lies in group $i$, $K_j$ is the total number of cases lies in group $j$.

The likelihood function:

$$Likelihood(\lambda) = (\prod_{k=1}^{K_i} \frac{\lambda^{b_{ij}(k)}e^{-\lambda}}{b_{ij}(k)!})(\prod_{k=1}^{K_j} \frac{(\frac{N_i}{N_j}\lambda)^{b_{ji}(k)}e^{-\frac{N_i}{N_j}\lambda}}{b_{ji}(k)!}) \tag{27}$$

The log likelihood function:

$$LogLikelihood(\lambda) \propto \sum_{k=1}^{K_i} log(\lambda^{b_{ij}(k)} e^{-\lambda}) + \sum_{k=1}^{K_j} log((\frac{N_i}{N_j}\lambda)^{b_{ji}(k)} e^{-\frac{N_i}{N_j}\lambda}) \quad (28)$$

$$= \sum_{k=1}^{K_i} log(\lambda^{b_{ij}(k)}) - K_i\lambda + \sum_{k=1}^{K_j} log((\frac{N_i}{N_j}\lambda)^{b_{ji}(k)}) - \frac{N_i}{N_j}K_j\lambda \quad (29)$$

$$\sum_{k=1}^{K_i} log(\lambda^{b_{ij}(k)}) + \sum_{k=1}^{K_j} log((\lambda)^{b_{ji}(k)}) - (K_i + \frac{N_i}{N_j}K_j)\lambda + \sum_{k=1}^{K} log((\frac{N_i}{N_j})^{b_{ji}(k)}) \quad (30)$$

$$\stackrel{def}{=} LL(\lambda) \quad (31)$$

Taking derivative of $LL(\lambda)$:

$$\frac{d}{d\lambda}LL(\lambda) = \sum_{k=1}^{K_i} \frac{(b_{ij}(k))\lambda^{b_{ij}(k)-1}}{\lambda^{b_{ij}(k)}} + \sum_{k=1}^{K_j} \frac{(b_{ji}(k))\lambda^{b_{ji}(k)-1}}{\lambda^{b_{ji}(k)}} - (K_i + \frac{N_i}{N_j}K_j) \quad (32)$$

$$= \sum_{k=1}^{K_i} \frac{b_{ij}(k)}{\lambda} + \sum_{k=1}^{K_j} \frac{b_{ji}(k)}{\lambda} - (K_i + \frac{N_i}{N_j}K_j) \quad (33)$$

$$= \frac{1}{\lambda}(\sum_{k=1}^{K_i} b_{ij}(k) + \sum_{k=1}^{K_j} b_{ji}(k)) - (K_i + \frac{N_i}{N_j}K_j) \quad (34)$$

The maximum likelihood estimation of $\lambda$ ($\lambda$ that maximizing $LL(\lambda)$) is obtained by letting $\frac{d}{d\lambda}LL(\lambda) = 0$:

$$\hat{\lambda} = \frac{\sum_{k=1}^{K_i} b_{ij}(k) + \sum_{k=1}^{K_j} b_{ji}(k)}{K_i + \frac{N_i}{N_j}K_j} \quad (35)$$

that is,

$$\hat{c}_{ij} = \hat{\lambda} \quad (36)$$

$$(37)$$

$$\hat{c}_{ji} = \frac{N_i}{N_j}\hat{c}_{ij} \quad (38)$$

$$(39)$$

#### 1.5 Results

Three proposed methods (LSE, WLSE, MLE) are adopted to the data set. The contact data matrix $A$ and estimated contact matrices $C$ are shown as follows (Figure 2). Further more, by using the MLE, the 95% confidential intervals for all entries of contact matrix are given in Figure 3.

We split the data set by the diagnosed date of cases at August 4, 00:00 (the peak of daily new cases), and estimating the contact matrices $C_1$, $C_2$ (contact matrix before and after the peak, see figure 4) using these two data set. The scatter plot of $C_1$, $C_2$ shows a strong linear correlation between entries of these two matrices (Figure **??**). Therefore, we processed a linear regression, and use the slope as a correction to $C_1$, $C_2$. There is an expected reduction of maximum eigenvalues of $C_1$ and $C_2$: $\lambda_{max}(C_1) = 32.17$, and $\lambda_{max}(C_2) = 24.58$.

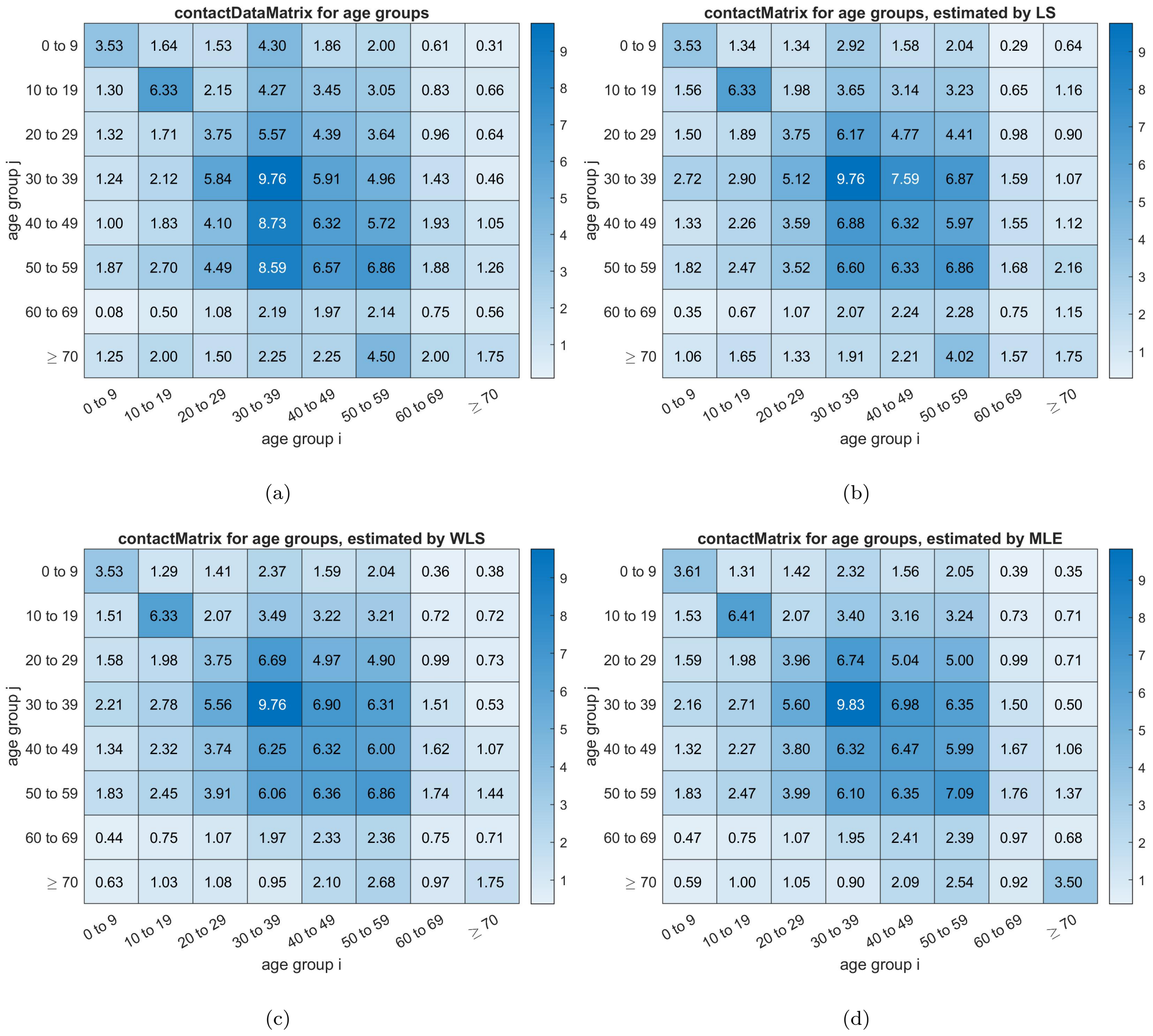


Fig. 2: Contact Data Matrix and Contact Matrices Estimated by different Methods: (a): The contact data matrix for age groups, filled with the case-contact data pairs; (b): The contact matrix for age groups, estimated by least square estimation; (c): The contact matrix for age groups, estimated by weighted least square estimation; (d): The contact matrix for age groups, estimated by maximum likelihood estimation.

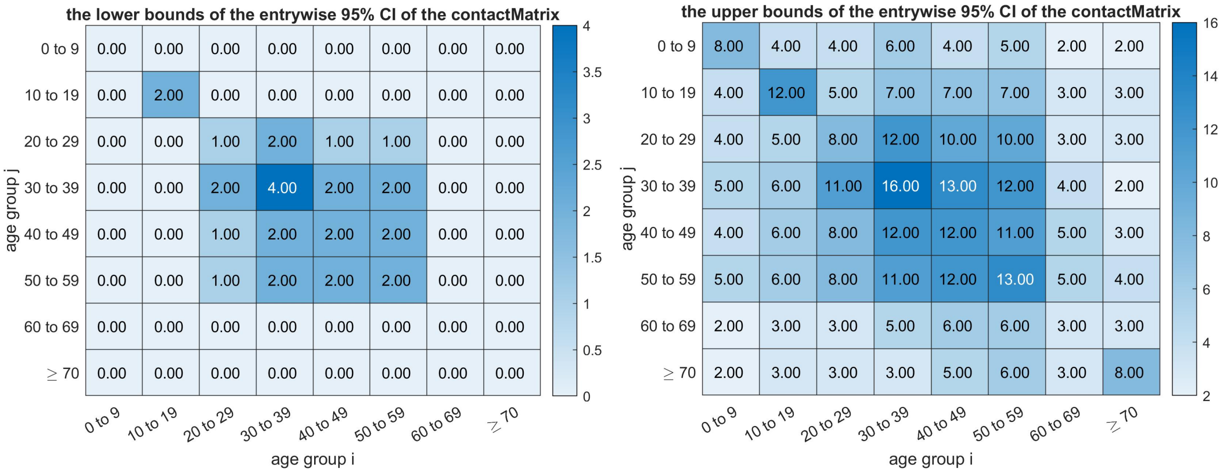


(a) (b)

Fig. 3: The entrywise estimation of 95% confidential interval of contact matrix, (a): the lower bound, (b): the upper bound

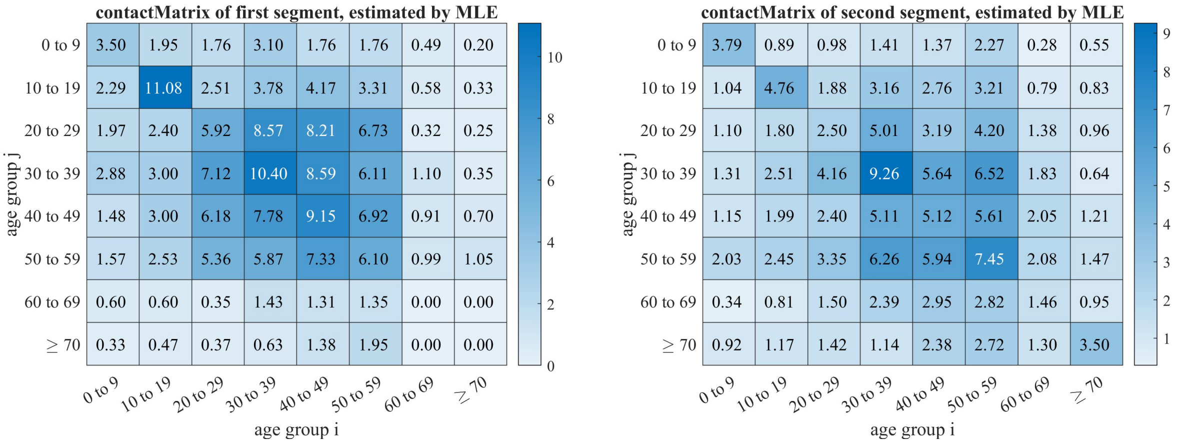


(a) (b)

Fig. 4: The estimated contact matrix before and after the peak of daily new incidents, (a): before the peak, with its eigenvalue equals to 32.17; (b): after the peak, with its eigenvalue equals to 24.58

### 2 Multi-Group SEIAR Simulation for given $R_0$

Due to the significant proportion of asymptomatic cases, the differences in transmissibilities of symptomatic and asymptomatic cases, and the influence of the latent and incubation period, the multi-group SEIAR framework is adopted for Covid-19 simulation.

#### 2.1 An Introduction to the Multi-Group SEIAR Model

The multi-group SEIAR model is a deterministic dynamic model based two assumptions: (1) the homogeneity of populations, (2) populations of groups are fully mixed (inside and between them), and described by the following ordinary differential equations (ODEs):

$$\begin{aligned}
\frac{dS_i}{dt} &= br_i N - S_i \sum_{j=1}^{n} \beta_{ji} \left(I_j + \kappa_1 F_j + \kappa_2 A_j\right) - dr_i S_i \\
\frac{dE_i}{dt} &= S_i \sum_{j=1}^{n} \beta_{ji} \left(I_j + \kappa A_j\right) - p_i \omega_i' E_i - (1 - p_i)\, \omega_i E_i - dr_i E_i \\
\frac{dI_i}{dt} &= (1 - p_i)\, \omega_i E_i - (dr_i + f_i + \gamma_i)\, I_i \\
\frac{dA_i}{dt} &= p_i {\omega_i}' E_i - (dr_i + \gamma_i')\, A_i \\
\frac{dR_i}{dt} &= \gamma_i I_i + \gamma_i' A_i - dr_i R_i \\
i &= 1, 2, \cdots, n
\end{aligned}$$

where

- subscript $i$ denotes the variable or parameter is specified to the $i$-th group
- $n$ denotes the total number of groups
- $S_i$ denotes the number of susceptible population in group $i$
- $E_i$ denotes the number of exposed population (i.e, infected but not infectious) population in group $i$
- $I_i$ denotes the number of symptomatic infectious population in group $i$
- $A_i$ denotes the number of asymptomatic infectious population in group $i$
- $R_i$ denotes the number of fully immunized (i.e. impossible to be infected) population in group $i$
- $N_i = S_i + E_i + I_i + A_i + R_i$ is the population size of group $i$
- $N = N_1 + N_2 + \cdots + N_n$ is the total population size
- $br_i$ denotes the age specific birth rate (for age-grouped model, only one $br_i$ of the youngest age group is nonzero)
- $dr_i$ denotes the mortality rate of group $i$
- $\beta_{ij}$ is the coefficients describing the daily transmission rate from group $i$ to group $j$

- $\kappa$ is the relatively of transmission ability of asymptomatic cases compared with the symptomatic cases, it is assumed to be group irrelevant

- $f_i$ is the case fatality rate of group $i$

- $p_i$ is the probability that one infected individual in group $i$ will developed into a asymptomatic case

- $\omega_i$ is inverse of the average latent period of the symptomatic population in the $i$-th group, it is used to quantify the remove rate of compartment $E_i$. The inverse of incubation period is usually adopted for practice

- $\omega_i^{'}$ is the inverse of the average latent period of the asymptomatic population in the $i$-th group

- $\gamma_i$ is the inverse of average infectious period for symptomatic cases

- $\gamma_i^{'}$ is the inverse of average infectious period for asymptomatic cases

#### 2.2 Formulation of The Newly Infection Term

The widely used newly infection in group $i$, $S_i \sum_{j=1}^{n} \beta_{ji} (I_j + \kappa A_j)$, is based on the assumption that the newly infections is proportional to the number of susceptible individuals and the number of infectious individuals, therefore, the constants $\beta_{ji}$ are introduced as coefficients of such proportion.

To introduce the impact of contact patterns in the grouped SEIAR model, the transmission rate coefficients $\beta_{ij}$ are formularized as the product of number of daily average contacts, the probability of infection after contact and the susceptibility of people in the $i$-th group, that is,

$$\beta_{ji} N_j = c_{ij} q \sigma_i \tag{40}$$

where $c_{ij}$ is the $ij$-th entry of contact matrix; $q$ is the probability of infection after a one time contact; $\sigma_j$ is the susceptibility of group $j$.

In this way, the newly infection term $S_i \sum_{j=1}^{n} \beta_{ji} (I_j + \kappa A_j)$ is formulated as:

$$S_i \sum_{j=1}^{n} c_{ij} q \sigma_i \frac{I_j + \kappa A_j}{N_j} \tag{41}$$

This formulation of newly infections can be interpreted as follows.

- At the beginning of an outbreak, people of infected takes only a small proportion in population, which means a arbitrarily picked individual $x$ is more likely to be susceptible.

- Hence, we shall consider how many infectious individuals that a susceptible will contact, rather than the number of susceptible individuals that an infectious individual will contact.

- Let $x_i$ denotes a susceptible individual in group $i$. Among all $\sum_{j=1}^{n} c_{ij}$ individuals $x_i$ had contacted (during a time step of one day), there will be averagely $\sum_{j=1}^{n} c_{ij} \frac{I_j+\kappa A_j}{N_j}$ infectious individuals. (Note that $\sum_{j=1}^{n} c_{ij} \frac{I_j+\kappa A_j}{N_j}$ is very small quantity, but it still showed significant results by summing up all susceptible individuals).

- Then, product by the probability $q$ of infection with a single infectious contact, and summing up all $S_i$ susceptible individuals, we have the expected number of infections in group $i$:

$$qS_i \sum_{j=1}^{n} c_{ij} \frac{I_j + \kappa A_j}{N_j} \tag{42}$$

- If we want to consider the different susceptibilities in different groups, then a parameter of relative susceptibility $\sigma_i$ could be introduced for each group:

$$q\sigma_i S_i \sum_{j=1}^{n} c_{ij} \frac{I_j + \kappa A_j}{N_j} \tag{43}$$

  Note that $\sigma_i$ is a relative quantity, that it should had a magnitude for $o(1)$.

#### 2.3 Profiles of Simulations

We defined a series basic reproduction number $R_0 = [0.5, 1.0, 1.5, \cdots, 6]$ for simulation. In the expression of $R_0$ of grouped SEIAR model, $R_0$ is formularized as a scalar valued function of $\beta_{ij}$ and other model parameters. Hence by replacing $\beta_{ij} = c_{ij} q \sigma_j / N_i$, the probability of infection $q$ can be solved from the expression of $R_0$.

For the sake of the balance between simplicity and accuracy of models, the parameters $\kappa$, $dr_i$, $f_i$, $p_i$, $\omega_i$, $\omega_i^{'}$, $\gamma_i$, $\gamma_i^{'}$, are both set to be group irrelevant, and represented by the corresponding variable without subscribe. And for the short term outbreak, the birth rate and the mortality are omitted, i.e. set as zeros.

#### 2.4 Solve $q$ From $R_0$

##### 2.4.1 The Next Generation $R_0$

The next generation method gives the following $R_0$ [Reference]:

$$R_0 = \frac{\lambda_{\max}(B)}{d_r + p\omega' + (1-p)\omega} \left[ \frac{\kappa p \omega'}{(\gamma' + d_r)} + \frac{(1-p)\omega}{(\gamma + d_r + f)} \right] \tag{44}$$

$$B = \begin{bmatrix} b_{11} & b_{12} & \cdots & b_{1n} \\ b_{21} & b_{22} & \cdots & b_{2n} \\ \vdots & \vdots & \ddots & \vdots \\ b_{n1} & b_{n2} & \cdots & b_{nn} \end{bmatrix} \tag{45}$$

where $\lambda_{\max}(\cdot)$ denotes the leading eigenvalue (with maximum real part) of a matrix; $b_{ij} = N_i \beta_{ji},\ \ i, j = 1, 2, \cdots, n$.

By substituting $\beta_{ij} = c_{ij} q \sigma_j / N_i$, we have:

$$q = R_0 / \left\{ \frac{\lambda_{\max}(B/q)}{d_r + p\omega' + (1-p)\omega} \left[ \frac{\kappa p \omega'}{(\gamma' + d_r)} + \frac{(1-p)\omega}{(\gamma + d_r + f)} \right] \right\} \tag{46}$$

Note that the right hand side of the above expression is irrelevant to $q$ ($q$ is eliminated in $B/q$), that is, one $R_0$ corresponding one probability of infection once the model parameters are fixed.

##### 2.4.2 The Definition Based $R_0$

By using the definition-based method, the interactive $R_{ij}$ (the expectation of secondary infections that one infected individual in group $i$ will produced in group $j$ during its lifespan as infectious) is firstly computed via its definition. Then the true $R_0$ is computed by taking expectation. [Reference] The result gives:

$$R_0 = \sum_{i=1}^{n} \left[ \frac{N_i}{N} \sum_{j=1}^{n} R_{ij} \right] \tag{47}$$

$$R_{ij} = \frac{\beta_{ij} N_j}{d_r + p\omega' + (1-p)\omega} \left( \frac{\kappa p \omega'}{(\gamma' + d_r)} + \frac{(1-p)\omega}{(\gamma + d_r + f_i)} \right) \tag{48}$$

Substitute $\beta_{ij} = c_{ij} q \sigma_j / N_i$, we have:

$$q = R_0 / \sum_{i=1}^{n} \left[ \frac{N_i}{N} \sum_{j=1}^{n} \frac{R_{ij}}{q} \right] \tag{49}$$

Note that the right hand side of the above expression is irrelevant to $q$ ($q$ is eliminated in $\frac{R_{ij}}{q}$), that is, one $R_0$ corresponding one probability of infection once the model parameters are fixed.

#### 2.5 Parameters for simulation

Once the probability of infection $q$ is solved form the expression of $R_0$, the coefficients of transmission $\beta_{ij}$ can be easily determined by $\beta_{ij} = c_{ij} q \sigma_i / N_i$. Therefore, we have all parameter needed for simulation (i.e. solve the ODE with given initial point).

The following chapter considers the vaccine effect, and generalizes the multi-group $SEIAR$ model into an $VEFIAQR$ model. All parameters for the $SEIAR$ simulation can be found in Table 2 in next chapter.

#### 2.6 $SEIAR$ Simulation Results

We start the simulation with case zero, and perform a 14-days and a 100-days simulation for identical parameters.

##### 2.6.1 A 14-days Simulation

#### Contents


- steup simulation
- probability vector q (DBM and NGM are adopted respectively), for infection by a one-time contact
- simulate for each self-defined base-line R0


```
clear all; close all; clc;

ImportParameters;   % import parameters omega, gamma, p, kappa,...
ImportFigureLegends; % import the following cell array for figure legend:
                     % groupLegend; ageLegend;
N = readmatrix('agePopulationVector.xlsx'); % the population vector (stratified by age)
C = readmatrix('contactMatrix.xlsx');   % the contact matrix
```

#### steup simulation

```
% initial value
xInit = [N, zeros(n,4)];
xInit(4,3) = 1;

% time span
tInit = 0;
tFinal = 14;

% step size
stepSize = 0.01;

% a series R0 for simulation
R0 = [0.2, 0.5:6.5];     R0 = R0';
```

#### probability vector q (DBM and NGM are adopted respectively), for infection by a one-time contact

```
q_DBM = zeros(numel(R0),1);
q_NGM = zeros(numel(R0),1);

for i = 1:n
    for j = 1:n
        R(i,j) = C(j,i) / (dr(i) + p(i)*omegap(i) + (1-p(i))*omega(i)) * (kappa(i)*p(i)*omegap(i)/(gammap(i)+dr(i)) + (1-p(i))*omega(i)/(gamma(i)+dr(i)+f(i)));
    end
end


for i = 1:numel(R0)
    temp1 = R .* (N.') ./ sum(N);
    temp2 = sum(temp1,1);
    q_DBM(i) = R0(i) / sum(temp2);

    temp3 = 1 / (dr(i) + p(i)*omegap(i) + (1-p(i))*omega(i)) * (kappa(i)*p(i)*omegap(i)/(gammap(i)+dr(i)) + (1-p(i))*omega(i)/(gamma(i)+dr(i)+f(i)));
    q_NGM(i) = R0(i) / (max(eig(C)) * temp3);
end
fprintf('[R0, q_DBM, q_NGM] = \n');
disp([R0, q_DBM, q_NGM]);
```

```
[R0, q_DBM, q_NGM] =
   2.0000e-01   1.5693e-03   1.3569e-03
   5.0000e-01   3.9233e-03   3.3922e-03
   1.5000e+00   1.1770e-02   1.0177e-02
   2.5000e+00   1.9617e-02   1.6961e-02
   3.5000e+00   2.7463e-02   2.3746e-02
   4.5000e+00   3.5310e-02   3.0530e-02
   5.5000e+00   4.3157e-02   3.7314e-02
   6.5000e+00   5.1003e-02   4.4099e-02
```

#### simulate for each self-defined base-line R0

```
for i = 1:numel(R0)
    params.Beta   = C * q_DBM(i) ./ (N');
%     params.Beta   = C * q_NGM(i) ./ (N');

    fun = @(x,t)dxdt_SEIAR(x,t,params);
    [x,t,S,E,I,A,R] = odeSolveRK4(fun, tInit, xInit, tFinal, stepSize);

    % currentSymptomaticCases
    figure;
    plot(t,I);
    legend(ageLegend);
    title(['current symptomatic cases \it{I}, with R_0 = ', num2str(R0(i))]);
    xlabel('time (in days)');
    ylabel('population size (in person)')

    % dailySymptomaticNewCases
    figure;
    dailySymptomaticNewCases = ((1-p).*omega)' .* E;
    plot(t,dailySymptomaticNewCases);
    legend(ageLegend);
    title(['daily new symptomatic cases \it{(1-p) \omega E}, with R_0 = ', num2str(R0(i))]);
    xlabel('time (in days)');
    ylabel('population size (in person)')

    % allCurrentCases
    figure;
    allCurrentCases = I + A;
    plot(t,allCurrentCases);
    legend(ageLegend);
    title(['all current cases \it{I + A}, with R_0 = ', num2str(R0(i))]);
    xlabel('time (in days)');
    ylabel('population size (in person)')

    % accumulativeNumberOfCases
```

```
    figure;
    dailyNewCases = (p.*omegap)' .* E + ((1-p).*omega)' .* E;
    accumulativeNumberOfCases = zeros(size(dailyNewCases,1), n);
    for k = 1:n
        accumulativeNumberOfCases(:,k) = cumtrapz(t, dailyNewCases(:,k));
    end
    plot(t,accumulativeNumberOfCases);
    legend(ageLegend);
    title(['accumulative number of cases, with R_0 = ', num2str(R0(i))]);
    xlabel('time (in days)');
    ylabel('population size (in person)')


    % how to define and compute the accumulativeNumberOfCases from model directly
    % instead of a extra numerical integration, it is still an very interesting question.
    Nt = sum(S + E + I + A + R, 2) / sum(N);
end
```

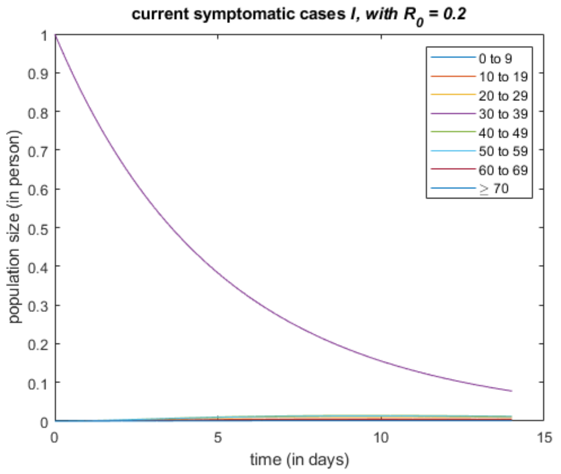


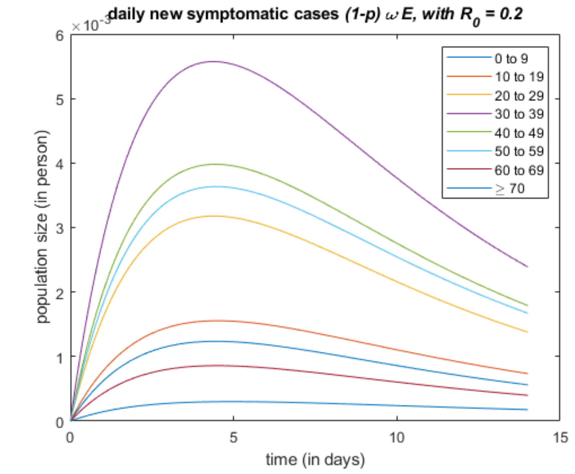


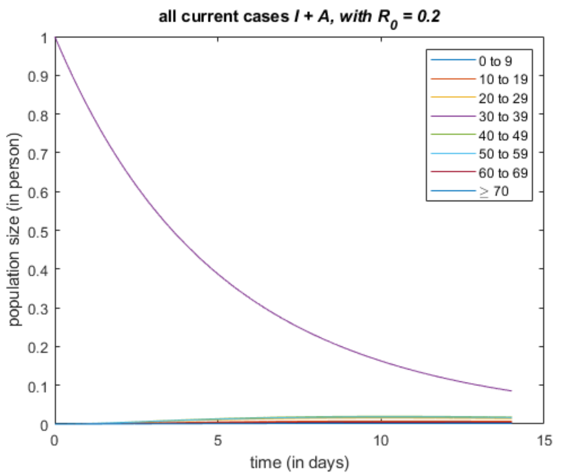


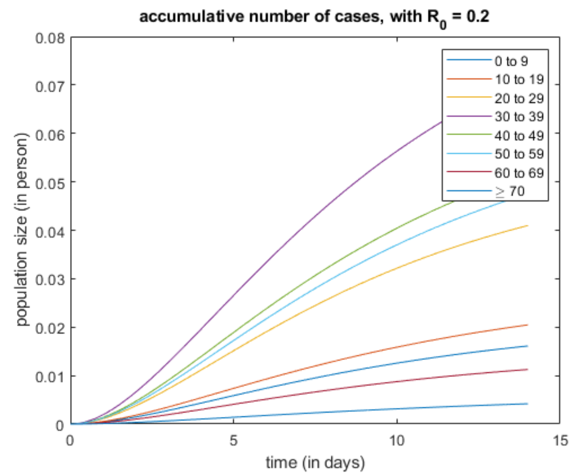


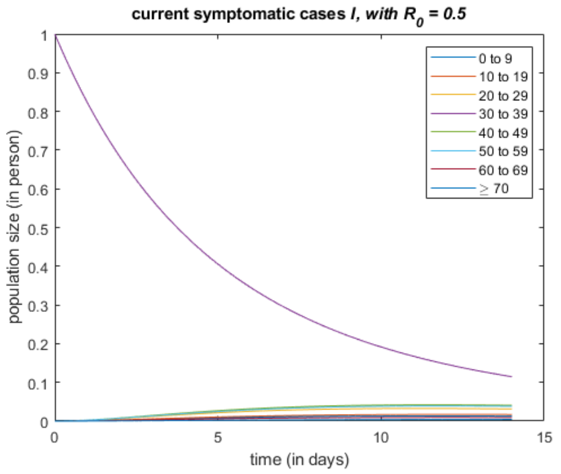


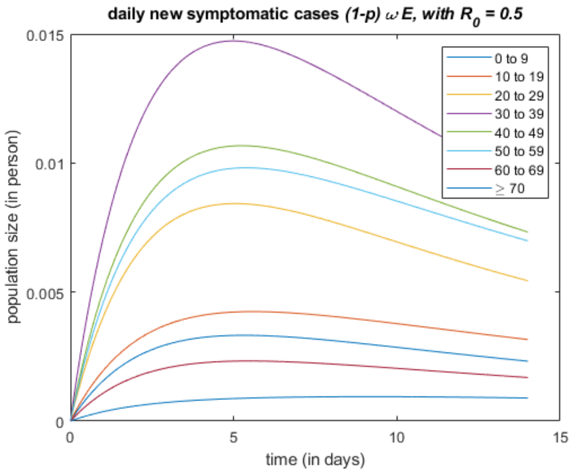

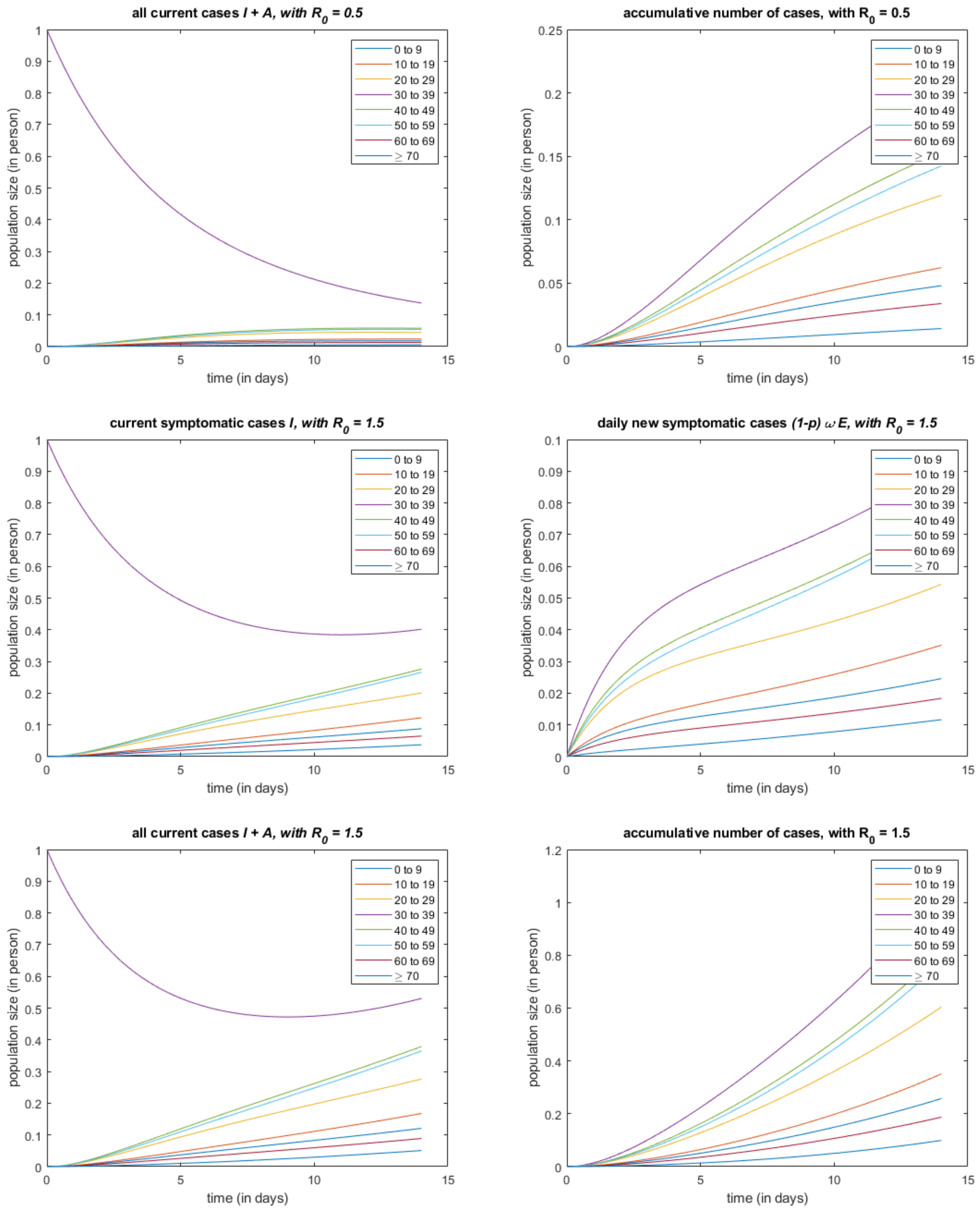
all current cases I + A, with $R_0$ = 0.5
accumulative number of cases, with $R_0$ = 0.5
current symptomatic cases I, with $R_0$ = 1.5
daily new symptomatic cases (1-p) $\omega$ E, with $R_0$ = 1.5
all current cases I + A, with $R_0$ = 1.5
accumulative number of cases, with $R_0$ = 1.5
population size (in person)
time (in days)
0 to 9
10 to 19
20 to 29
30 to 39
40 to 49
50 to 59
60 to 69
≥ 70

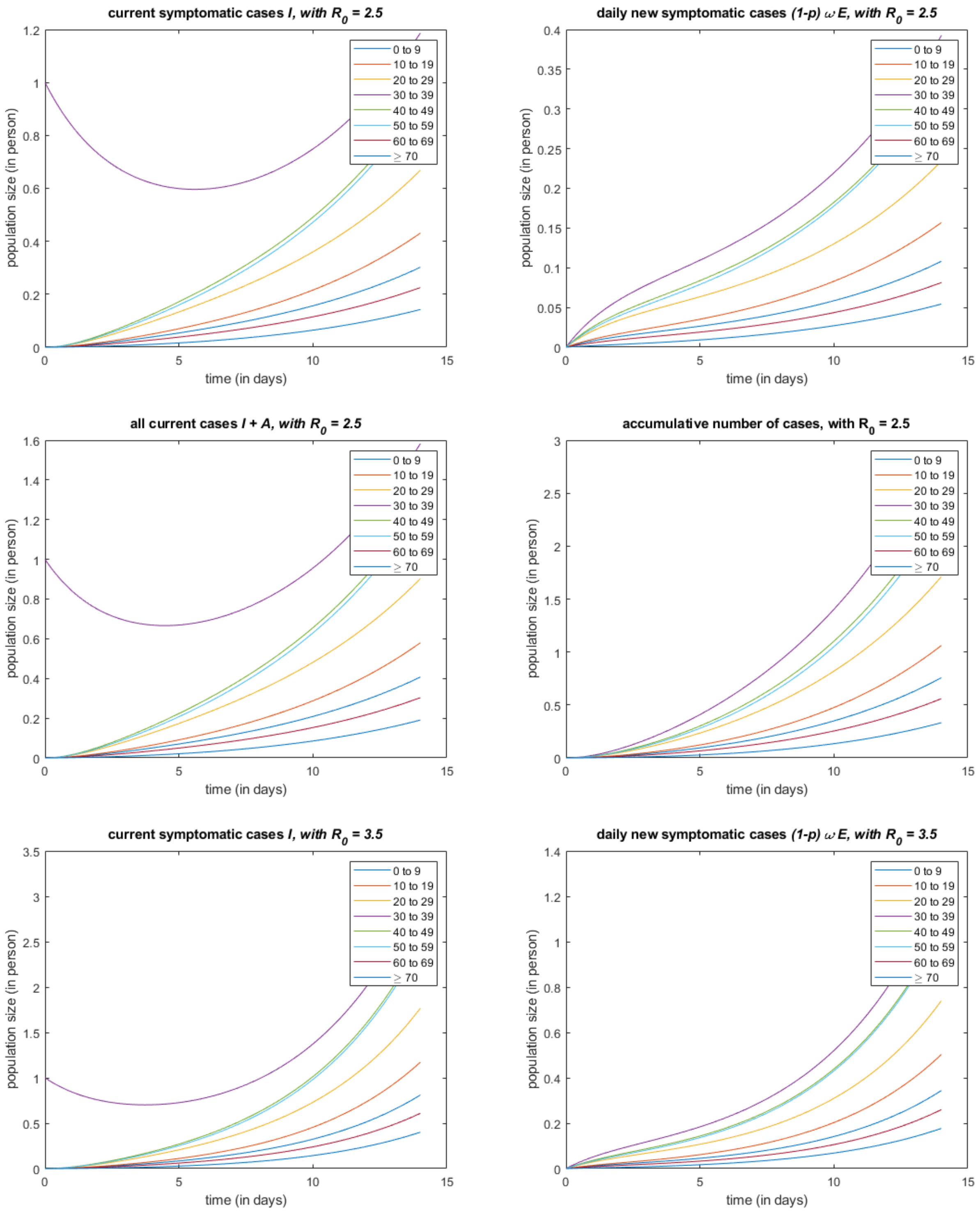

current symptomatic cases I, with R0 = 2.5
daily new symptomatic cases (1-p) ω E, with R0 = 2.5
all current cases I + A, with R0 = 2.5
accumulative number of cases, with R0 = 2.5
current symptomatic cases I, with R0 = 3.5
daily new symptomatic cases (1-p) ω E, with R0 = 3.5
population size (in person)
time (in days)
0 to 9
10 to 19
20 to 29
30 to 39
40 to 49
50 to 59
60 to 69
≥ 70

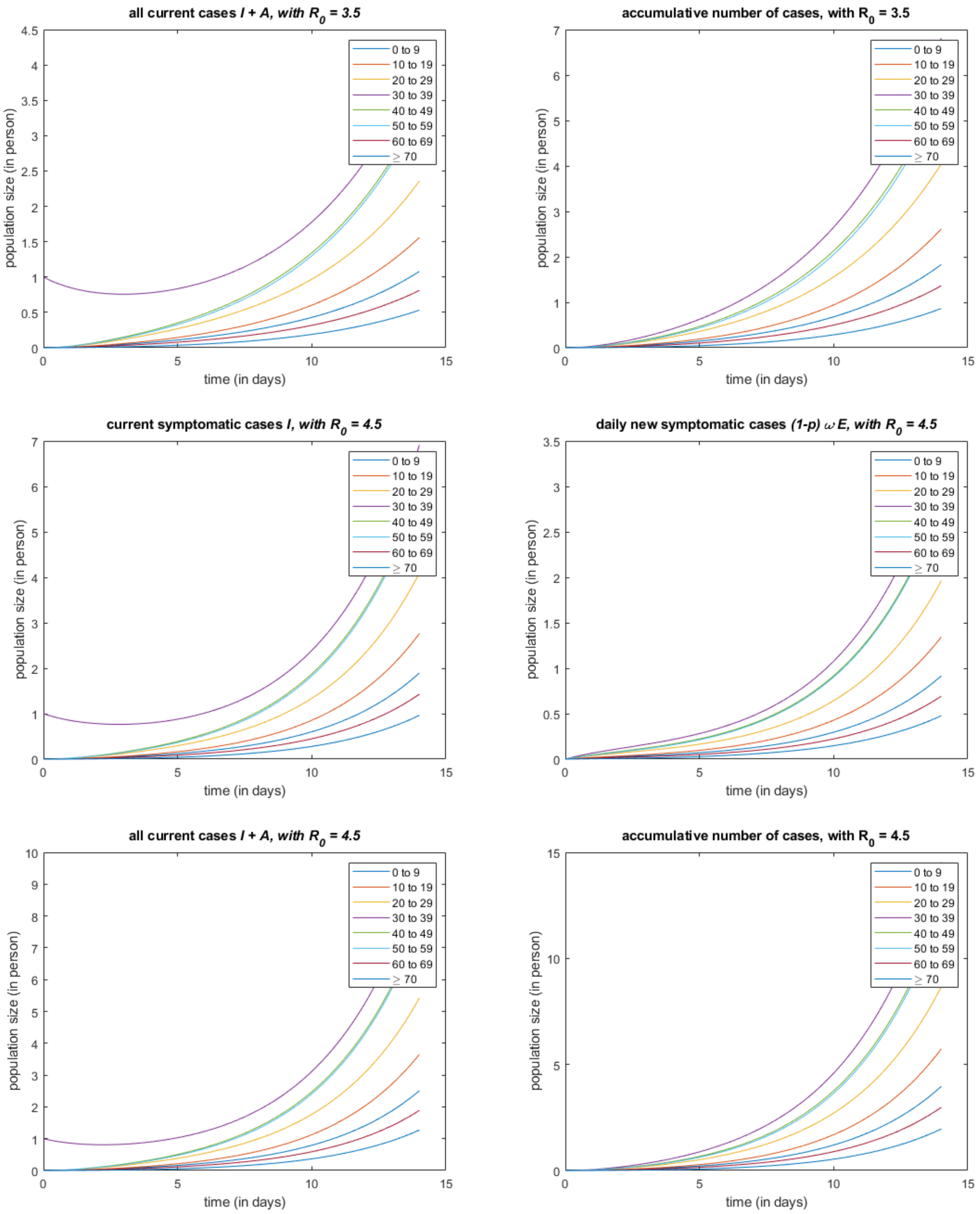
all current cases I + A, with $R_0$ = 3.5
accumulative number of cases, with $R_0$ = 3.5
current symptomatic cases I, with $R_0$ = 4.5
daily new symptomatic cases (1-p) ω E, with $R_0$ = 4.5
all current cases I + A, with $R_0$ = 4.5
accumulative number of cases, with $R_0$ = 4.5
population size (in person)
time (in days)
0 to 9
10 to 19
20 to 29
30 to 39
40 to 49
50 to 59
60 to 69
≥ 70

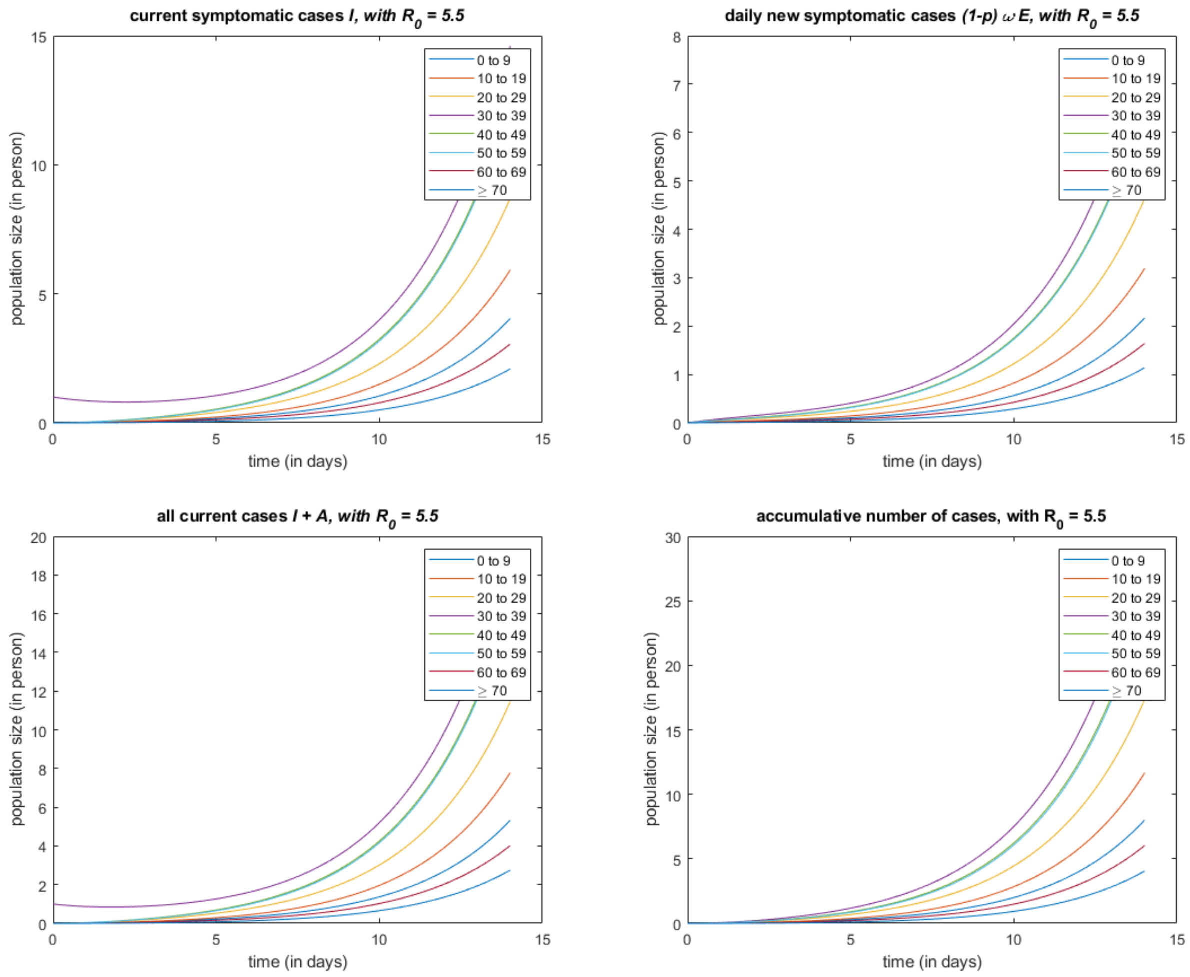

current symptomatic cases I, with $R_0$ = 5.5
daily new symptomatic cases (1-p) $\omega$ E, with $R_0$ = 5.5
all current cases I + A, with $R_0$ = 5.5
accumulative number of cases, with $R_0$ = 5.5
population size (in person)
time (in days)
0 to 9
10 to 19
20 to 29
30 to 39
40 to 49
50 to 59
60 to 69
≥ 70

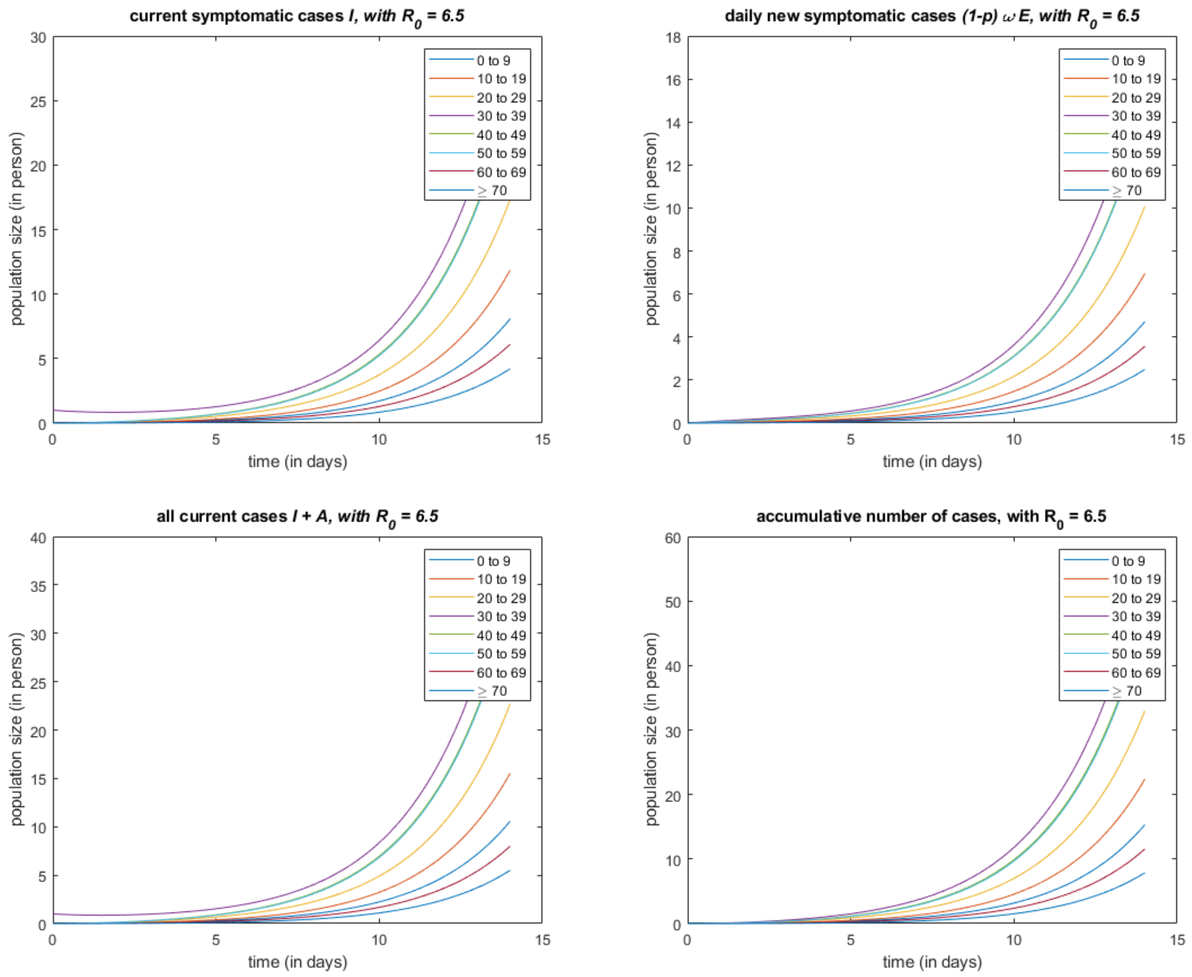


*Published with MATLAB® R2021a*

##### 2.6.2 A 100-days Simulation

#### Contents


- steup simulation
- probability vector q (DBM and NGM are adopted respectively), for infection by a one-time contact
- simulate for each self-defined base-line R0


```
clear all; close all; clc;

ImportParameters;   % import parameters omega, gamma, p, kappa,...
ImportFigureLegends; % import the following cell array for figure legend:
                     % groupLegend; ageLegend;
N = readmatrix('agePopulationVector.xlsx'); % the population vector (stratified by age)
C = readmatrix('contactMatrix.xlsx');   % the contact matrix
```

#### steup simulation

```
% initial value
xInit = [N, zeros(n,4)];
xInit(4,3) = 1;

% time span
tInit = 0;
tFinal = 100;

% step size
stepSize = 0.01;

% a series R0 for simulation
R0 = [0.2, 0.5:6.5];     R0 = R0';
```

#### probability vector q (DBM and NGM are adopted respectively), for infection by a one-time contact

```
q_DBM = zeros(numel(R0),1);
q_NGM = zeros(numel(R0),1);

for i = 1:n
    for j = 1:n
        R(i,j) = C(j,i) / (dr(i) + p(i)*omegap(i) + (1-p(i))*omega(i)) * (kappa(i)*p(i)*omegap(i)/(gammap(i)+dr(i)) + (1-p(i))*omega(i)/(gamma(i)+dr(i)+f(i)));
    end
end


for i = 1:numel(R0)
    temp1 = R .* (N.') ./ sum(N);
    temp2 = sum(temp1,1);
    q_DBM(i) = R0(i) / sum(temp2);

    temp3 = 1 / (dr(i) + p(i)*omegap(i) + (1-p(i))*omega(i)) * (kappa(i)*p(i)*omegap(i)/(gammap(i)+dr(i)) + (1-p(i))*omega(i)/(gamma(i)+dr(i)+f(i)));
    q_NGM(i) = R0(i) / (max(eig(C)) * temp3);
end
fprintf('[R0, q_DBM, q_NGM] = \n');
disp([R0, q_DBM, q_NGM]);
```

```
[R0, q_DBM, q_NGM] =
   2.0000e-01   1.5693e-03   1.3569e-03
   5.0000e-01   3.9233e-03   3.3922e-03
   1.5000e+00   1.1770e-02   1.0177e-02
   2.5000e+00   1.9617e-02   1.6961e-02
   3.5000e+00   2.7463e-02   2.3746e-02
   4.5000e+00   3.5310e-02   3.0530e-02
   5.5000e+00   4.3157e-02   3.7314e-02
   6.5000e+00   5.1003e-02   4.4099e-02
```

#### simulate for each self-defined base-line R0

```
for i = 1:numel(R0)
    params.Beta   = C * q_DBM(i) ./ (N');
%     params.Beta   = C * q_NGM(i) ./ (N');

    fun = @(x,t)dxdt_SEIAR(x,t,params);
    [x,t,S,E,I,A,R] = odeSolveRK4(fun, tInit, xInit, tFinal, stepSize);

    % currentSymptomaticCases
    figure;
    plot(t,I);
    legend(ageLegend);
    title(['current symptomatic cases \it{I}, with R_0 = ', num2str(R0(i))]);
    xlabel('time (in days)');
    ylabel('population size (in person)')

    % dailySymptomaticNewCases
    figure;
    dailySymptomaticNewCases = ((1-p).*omega)' .* E;
    plot(t,dailySymptomaticNewCases);
    legend(ageLegend);
    title(['daily new symptomatic cases \it{(1-p) \omega E}, with R_0 = ', num2str(R0(i))]);
    xlabel('time (in days)');
    ylabel('population size (in person)')

    % allCurrentCases
    figure;
    allCurrentCases = I + A;
    plot(t,allCurrentCases);
    legend(ageLegend);
    title(['all current cases \it{I + A}, with R_0 = ', num2str(R0(i))]);
    xlabel('time (in days)');
    ylabel('population size (in person)')

    % accumulativeNumberOfCases
```

```
    figure;
    dailyNewCases = (p.*omegap)' .* E + ((1-p).*omega)' .* E;
    accumulativeNumberOfCases = zeros(size(dailyNewCases,1), n);
    for k = 1:n
        accumulativeNumberOfCases(:,k) = cumtrapz(t, dailyNewCases(:,k));
    end
    plot(t,accumulativeNumberOfCases);
    legend(ageLegend);
    title(['accumulative number of cases, with R_0 = ', num2str(R0(i))]);
    xlabel('time (in days)');
    ylabel('population size (in person)')


    % how to define and compute the accumulativeNumberOfCases from model directly
    % instead of a extra numerical integration, it is still an very interesting question.
    Nt = sum(S + E + I + A + R, 2) / sum(N);
end
```

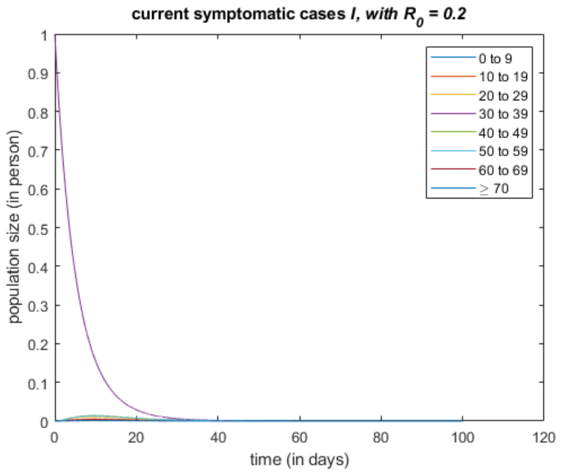


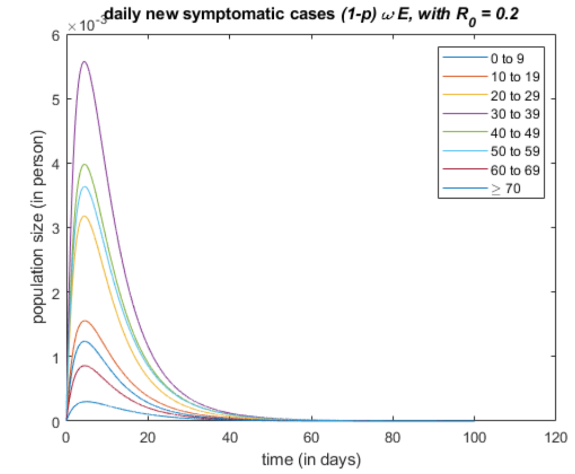


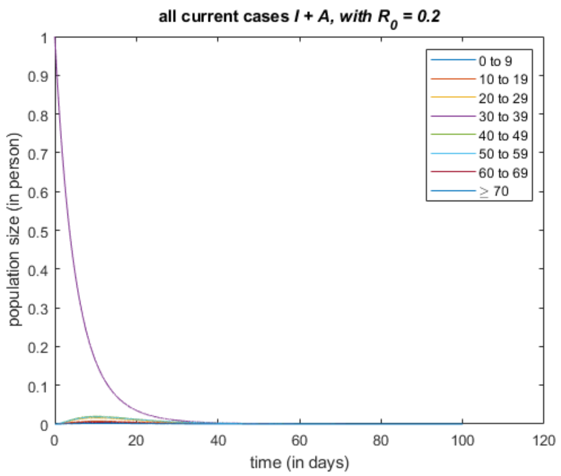


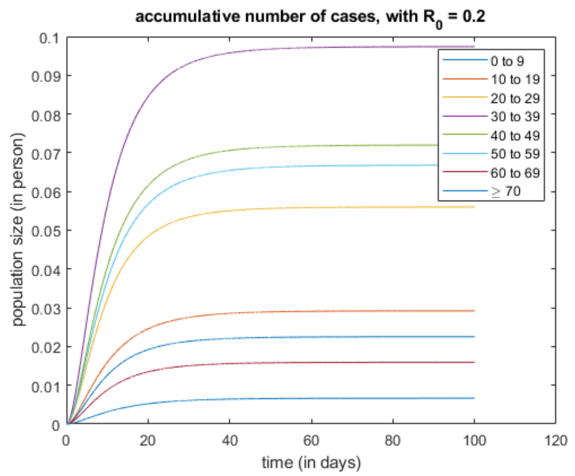


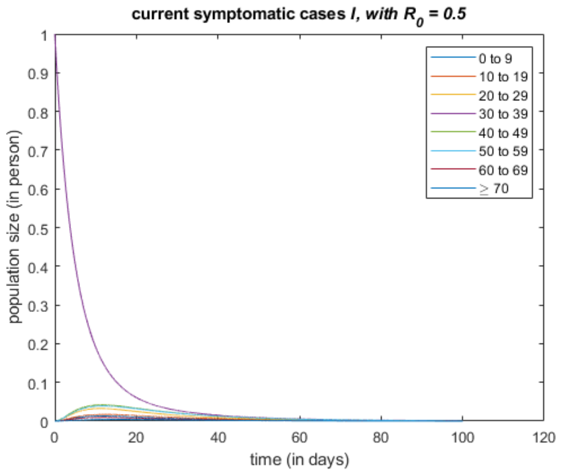


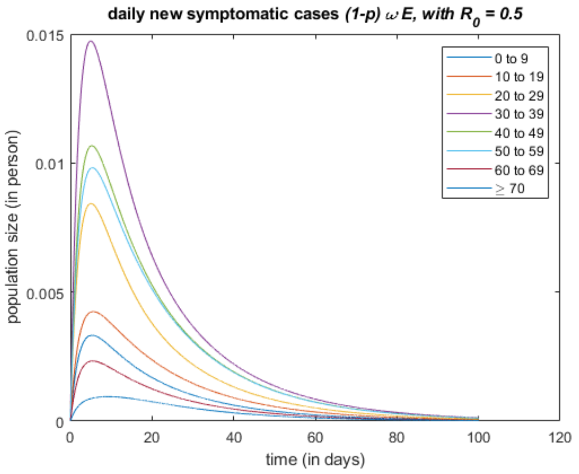

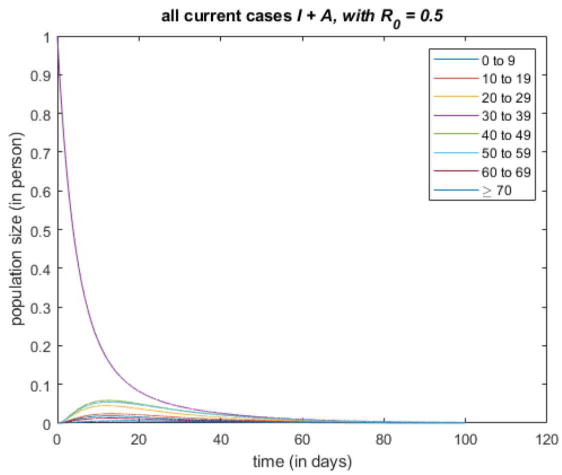
all current cases I + A, with R0 = 0.5
population size (in person)
time (in days)
0 to 9
10 to 19
20 to 29
30 to 39
40 to 49
50 to 59
60 to 69
≥ 70

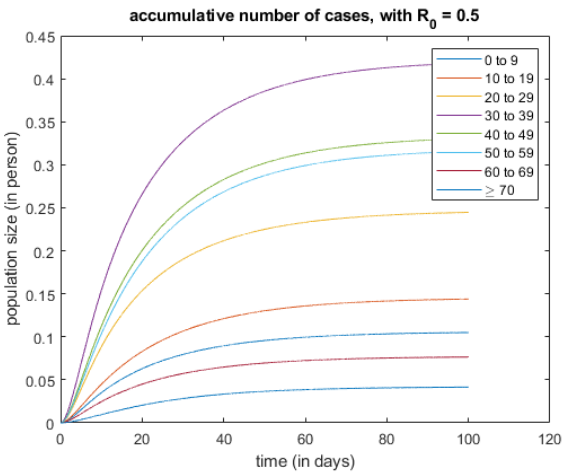
accumulative number of cases, with R0 = 0.5
population size (in person)
time (in days)
0 to 9
10 to 19
20 to 29
30 to 39
40 to 49
50 to 59
60 to 69
≥ 70

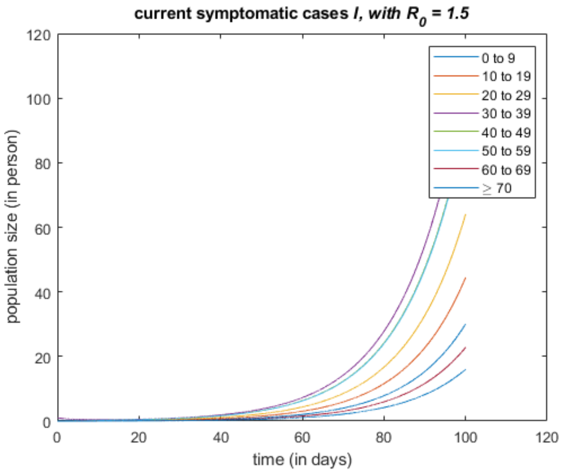
current symptomatic cases I, with R0 = 1.5
population size (in person)
time (in days)
0 to 9
10 to 19
20 to 29
30 to 39
40 to 49
50 to 59
60 to 69
≥ 70

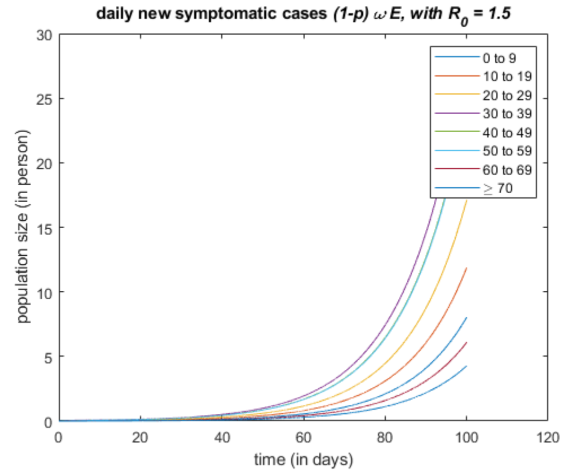
daily new symptomatic cases (1-p) ω E, with R0 = 1.5
population size (in person)
time (in days)
0 to 9
10 to 19
20 to 29
30 to 39
40 to 49
50 to 59
60 to 69
≥ 70

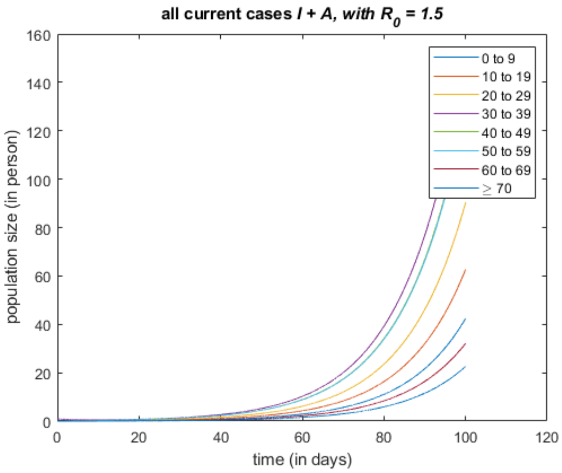
all current cases I + A, with R0 = 1.5
population size (in person)
time (in days)
0 to 9
10 to 19
20 to 29
30 to 39
40 to 49
50 to 59
60 to 69
≥ 70

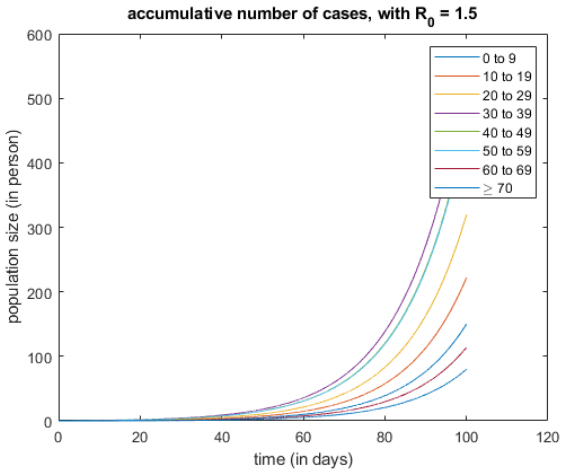
accumulative number of cases, with R0 = 1.5
population size (in person)
time (in days)
0 to 9
10 to 19
20 to 29
30 to 39
40 to 49
50 to 59
60 to 69
≥ 70

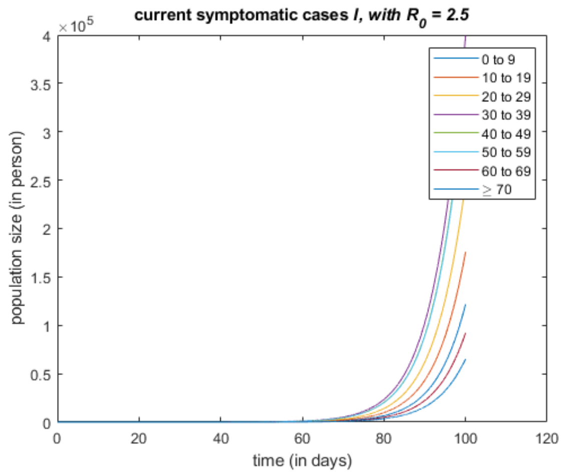
current symptomatic cases I, with $R_0$ = 2.5
population size (in person)
time (in days)
0 to 9
10 to 19
20 to 29
30 to 39
40 to 49
50 to 59
60 to 69
≥ 70

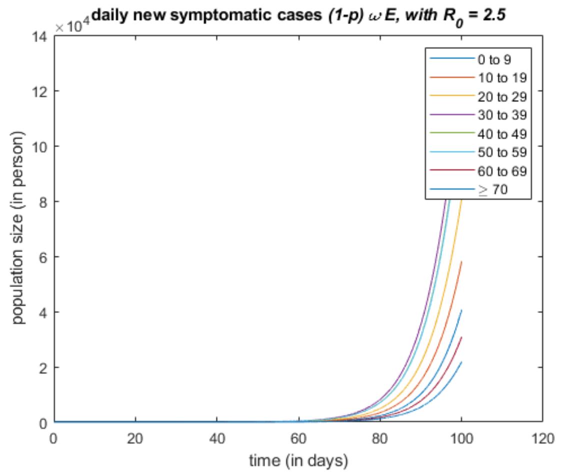
daily new symptomatic cases (1-p) ω E, with $R_0$ = 2.5
population size (in person)
time (in days)
0 to 9
10 to 19
20 to 29
30 to 39
40 to 49
50 to 59
60 to 69
≥ 70

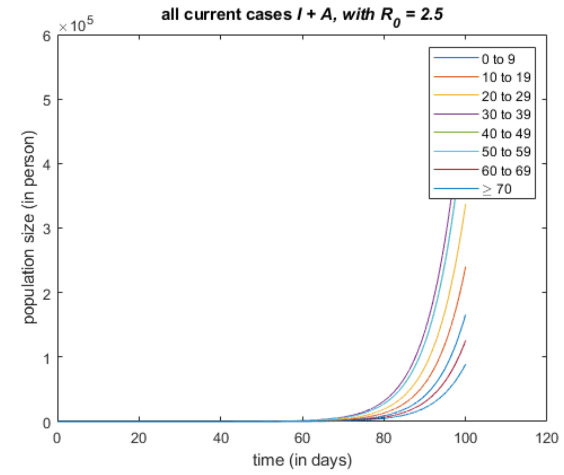
all current cases I + A, with $R_0$ = 2.5
population size (in person)
time (in days)
0 to 9
10 to 19
20 to 29
30 to 39
40 to 49
50 to 59
60 to 69
≥ 70

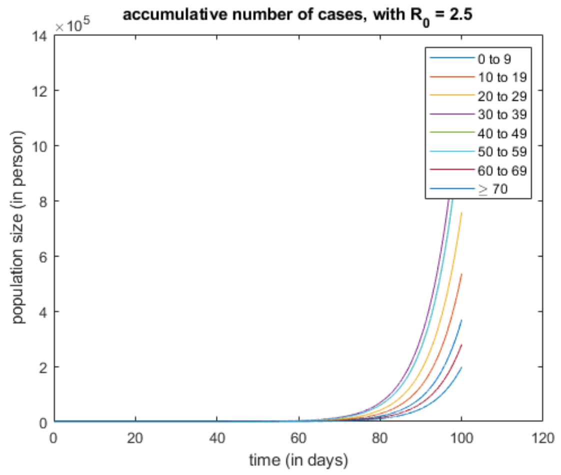
accumulative number of cases, with $R_0$ = 2.5
population size (in person)
time (in days)
0 to 9
10 to 19
20 to 29
30 to 39
40 to 49
50 to 59
60 to 69
≥ 70

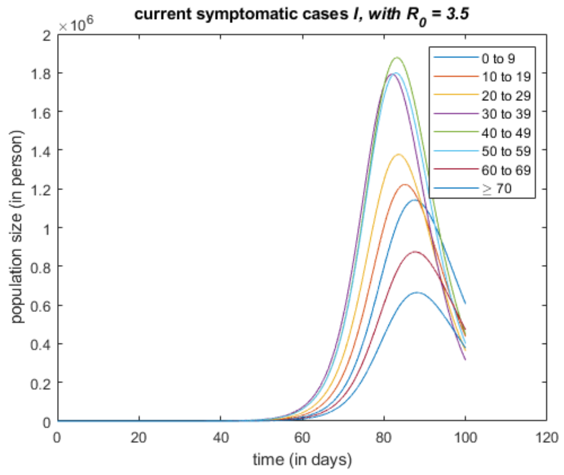
current symptomatic cases I, with $R_0$ = 3.5
population size (in person)
time (in days)
0 to 9
10 to 19
20 to 29
30 to 39
40 to 49
50 to 59
60 to 69
≥ 70

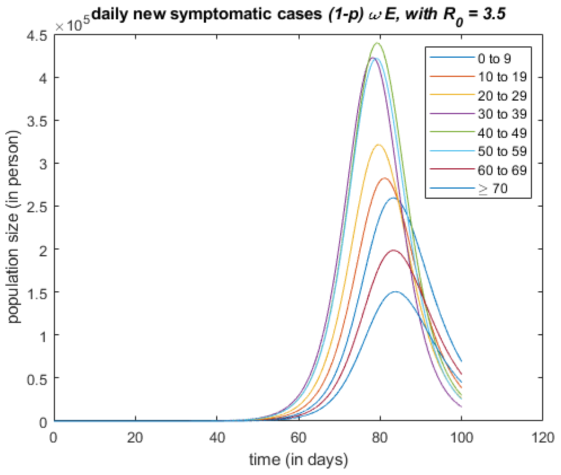
daily new symptomatic cases (1-p) ω E, with $R_0$ = 3.5
population size (in person)
time (in days)
0 to 9
10 to 19
20 to 29
30 to 39
40 to 49
50 to 59
60 to 69
≥ 70

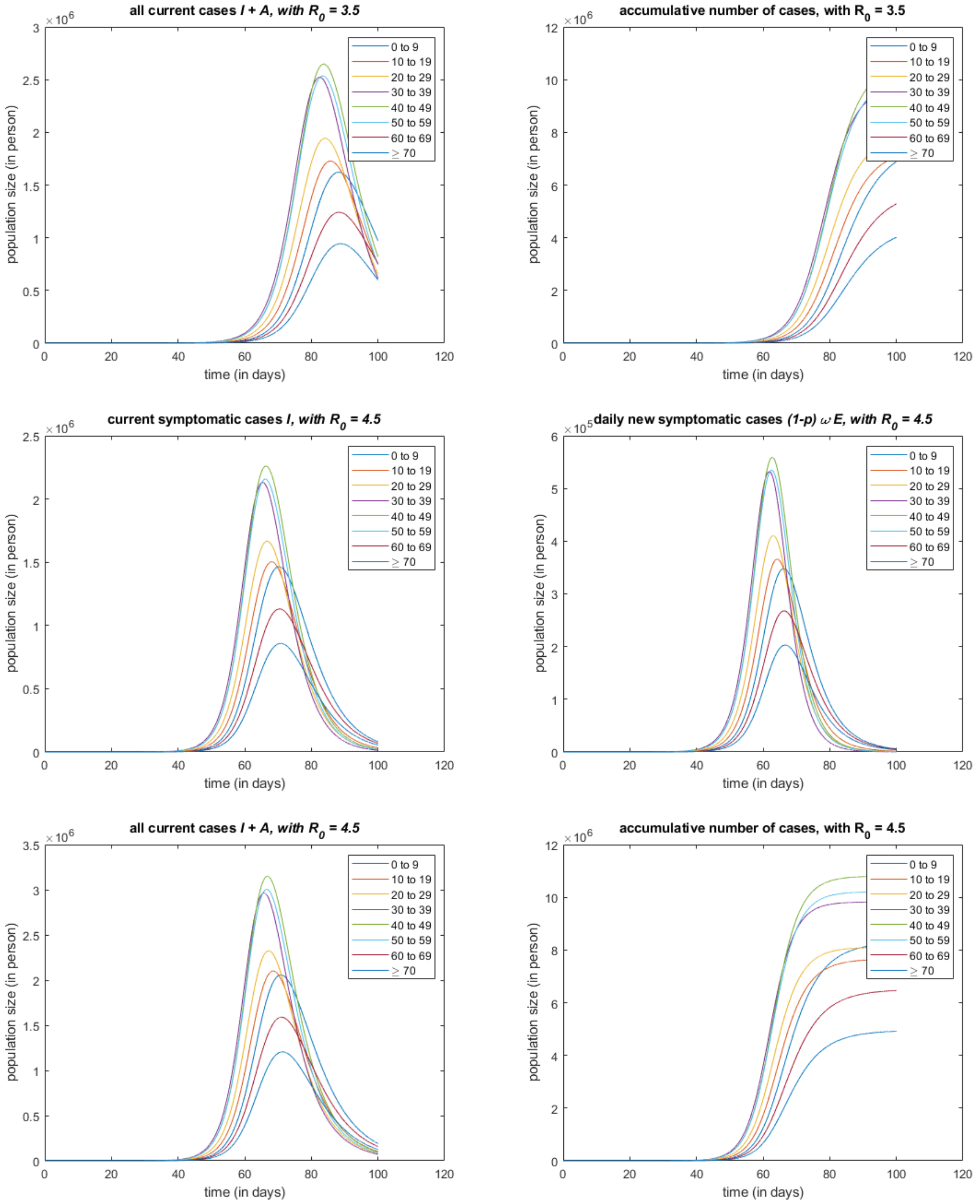
all current cases I + A, with $R_0$ = 3.5
accumulative number of cases, with $R_0$ = 3.5
current symptomatic cases I, with $R_0$ = 4.5
daily new symptomatic cases (1-p) ω E, with $R_0$ = 4.5
all current cases I + A, with $R_0$ = 4.5
accumulative number of cases, with $R_0$ = 4.5
population size (in person)
time (in days)
0 to 9
10 to 19
20 to 29
30 to 39
40 to 49
50 to 59
60 to 69
≥ 70

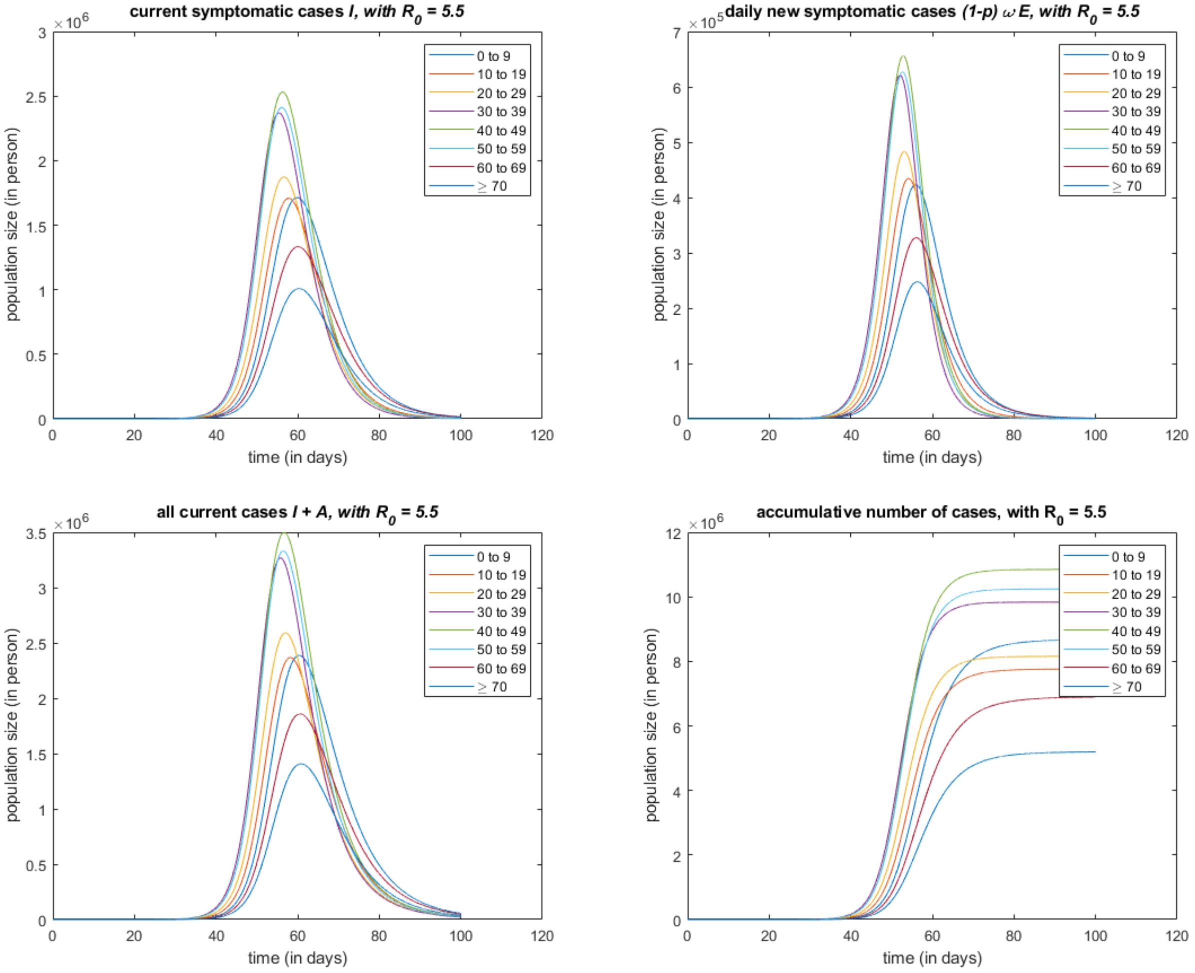
current symptomatic cases I, with R0 = 5.5
daily new symptomatic cases (1-p) ω E, with R0 = 5.5
all current cases I + A, with R0 = 5.5
accumulative number of cases, with R0 = 5.5
population size (in person)
time (in days)
0 to 9
10 to 19
20 to 29
30 to 39
40 to 49
50 to 59
60 to 69
≥ 70

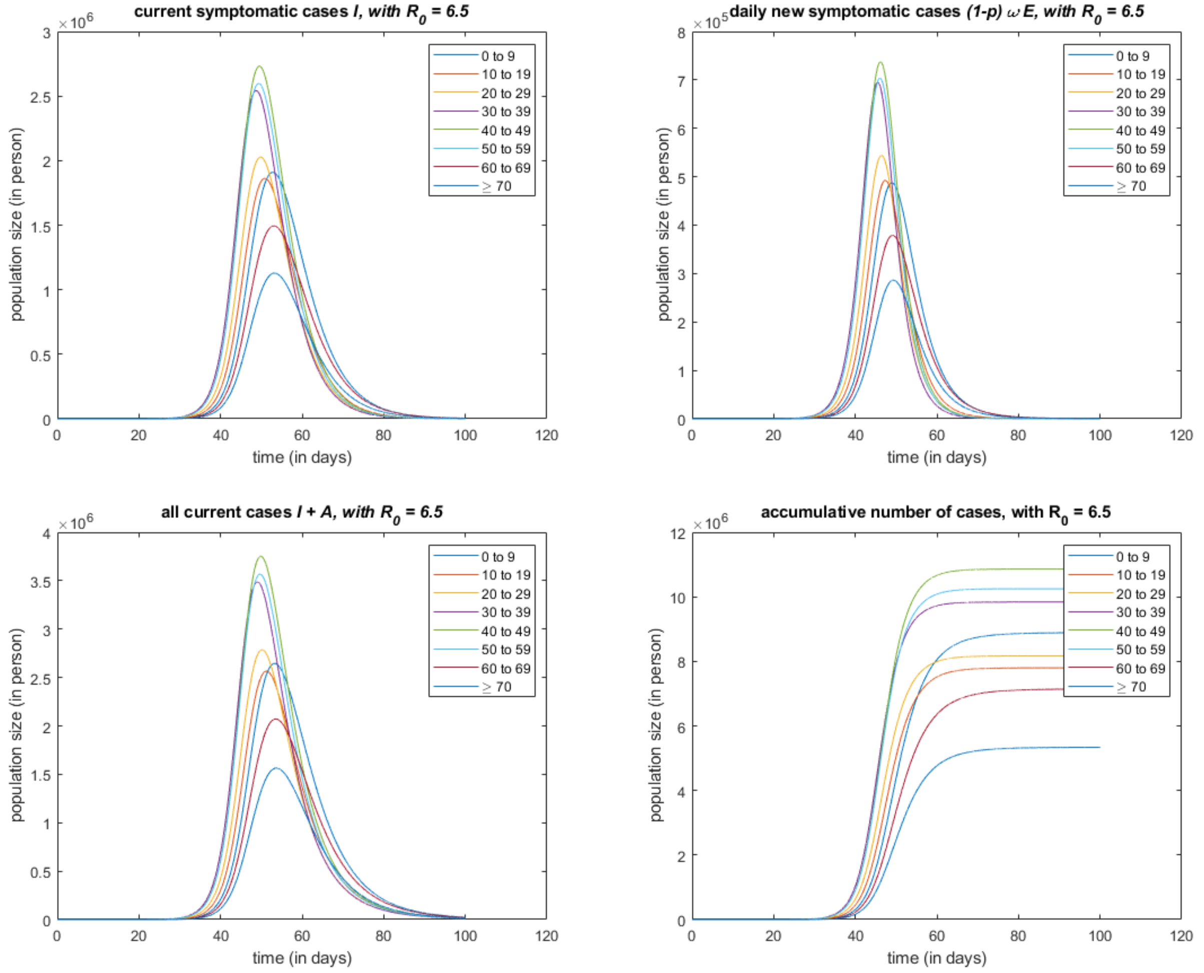


*Published with MATLAB® R2021a*

### 3 Vaccination In Populations

In China, while the vaccinating campaign proceeds steadily, there are only small outbreaks scattering, which attributes to the effectiveness of the social distancing policy. Therefore, for considering a short term outbreak, we might omit the vaccinate progress and treat the vaccine converge rate as constant in each age groups.

Such simplification is natural, however, makes it possible to consider the efficiency of vaccine without introducing extra compartments in the grouped SEIAR model (new compartments are still required to evaluate the dynamical vaccination).

To consider the efficiency of quarantine in NPIs, a new compartment $Q$ represent quarantine is added in our model. We also added a compartment $F$, represent the initial state of been infectious. The flowchart of the $VEFIAQR$ model is showed in Figure 1.

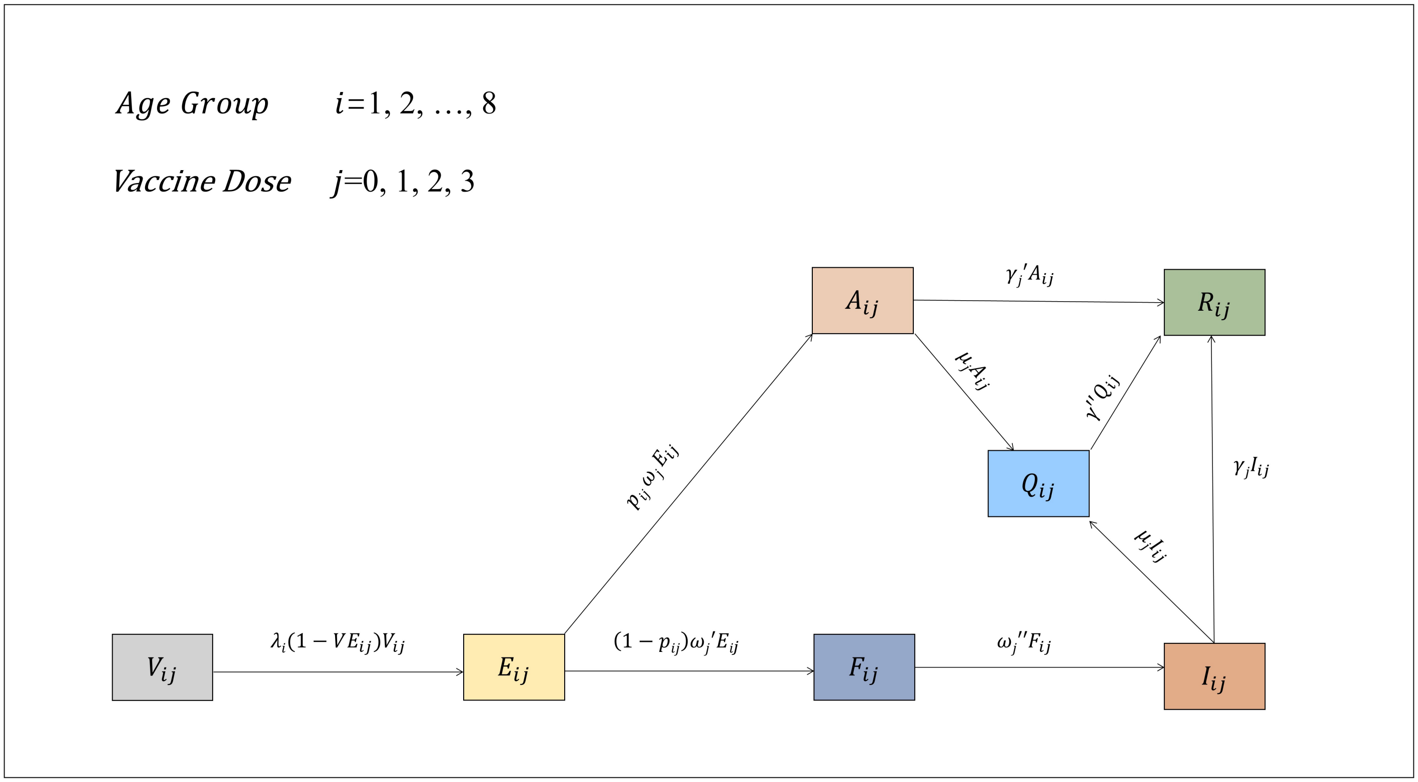


Fig. 5: Flowchart of The VEFIAQR Model

Tab. 1: Model Compartments

| Compartment | Meaning |
|---|---|
| group $i$ | age group $i$ |
| group $ij$ | population of those vaccinated $j$ doses in age group $i$ |
| $V_{ij}$ | population of susceptible in group $ij$ |
| $E_{ij}$ | those infected but not yet infectious in group $ij$ |
| $F_{ij}$ | pre-symptom state in group $ij$, which is infectious |
| $I_{ij}$ | symptomatic cases in group $ij$ |
| $A_{ij}$ | asymptomatic cases in group $ij$ |
| $Q_{ij}$ | quarantined in group $ij$ |
| $R_{ij}$ | removed in group $ij$ |

Tab. 2: Model Parameters

| Parameter | Meaning | Value (Range) | Reference |
|---|---|---|---|
| $n$ | number of age groups | 8 | self-defined |
| $c_{ij}$ | daily average number of contact in age group $j$ that a individual in age group $i$ will produce | positive | data estimated |
| $\beta_{ij}$ | transmission rate | positive | compute by $c_{ij}$ and $R_0$ |
| $VE_{ij}$ | the average vaccine efficacy of age group $i$ with $j$-th doses | [0, 0.4, 0.56, 0.802] | reference |
| $\lambda_i$ | force of infection acted to group $i$ | positive | compute from $beta_{ij}$, $E_{ij}$, $A_{ij}$ and $I_{ij}$ |
| $1/\omega_{ij}$ | average latent period of those vaccinated $j$ doses in age group $i$ | 3 | reference |
| $1/\omega_{ij}^{'}$ | difference of average incubation period and average latent period, of those vaccinated $j$ doses in age group $i$ | 5 | reference |
| $1/\omega_{ij}^{''}$ | connector between $F_{ij}$ and $A_{ij}$ | equal to $\omega_{ij}^{'}$ | by $\omega_{ij}^{'}$ |
| $p_{ij}$ | probability that one exposed individual in those vaccinated $j$ doses in age group $i$ will become symptomatic | [0.3, 0.4, 0.5, 0.7] | reference |
| $\mu_{ij}$ | rate of quarantine | positive | assumed |
| $1/\gamma$ | average infectious period of those symptomatic vaccinated $j$ doses in age group $i$ | 5 | reference |
| $1/\gamma^{'}$ | average infectious period of those asymptomatic vaccinated $j$ doses in age group $i$ | 10 | reference |
| $1/\gamma^{''}$ | average recover rate by quarantine | $\infty$ | assumed |

Tab. 3: Group Stratified Vaccine Efficacy $VE_{ij}$

| age group (year) | none vaccinated | un-fully vaccinated | fully vaccinated | booster vaccinated |
|---|---|---|---|---|
| 0 to 9 | 0 | 0.4 | 0.56 | 0.802 |
| 10 to 19 | 0 | 0.4 | 0.56 | 0.802 |
| 20 to 29 | 0 | 0.4 | 0.56 | 0.802 |
| 30 to 39 | 0 | 0.4 | 0.56 | 0.802 |
| 40 to 49 | 0 | 0.4 | 0.56 | 0.802 |
| 50 to 59 | 0 | 0.4 | 0.56 | 0.802 |
| 60 to 69 | 0 | 0.4 | 0.56 | 0.802 |
| ≥70 | 0 | 0.4 | 0.56 | 0.802 |

Tab. 4: Group Stratified Asymptomatic Proportion $p_{ij}$

| age group (year) | none vaccinated | un-fully vaccinated | fully vaccinated | booster vaccinated |
|---|---|---|---|---|
| 0 to 9 | 0.3 | 0.4 | 0.5 | 0.7 |
| 10 to 19 | 0.3 | 0.4 | 0.5 | 0.7 |
| 20 to 29 | 0.3 | 0.4 | 0.5 | 0.7 |
| 30 to 39 | 0.3 | 0.4 | 0.5 | 0.7 |
| 40 to 49 | 0.3 | 0.4 | 0.5 | 0.7 |
| 50 to 59 | 0.3 | 0.4 | 0.5 | 0.7 |
| 60 to 69 | 0.3 | 0.4 | 0.5 | 0.7 |
| ≥70 | 0.3 | 0.4 | 0.5 | 0.7 |

Tab. 5: Age-Stratified Vaccine Count via Data

| age group (year) | 0 dose | 1 dose | 2 doses | 3 doses |
|---|---|---|---|---|
| 0 to 9 | 838 | 1 | 5 | 2 |
| 10 to 19 | 582 | 609 | 277 | 31 |
| 20 to 29 | 537 | 225 | 1260 | 119 |
| 30 to 39 | 806 | 374 | 2388 | 291 |
| 40 to 49 | 460 | 192 | 1738 | 216 |
| 50 to 59 | 383 | 201 | 1463 | 274 |
| 60 to 69 | 174 | 112 | 378 | 66 |
| ≥70 | 144 | 97 | 143 | 19 |

Tab. 6: Age-Stratified Vaccine Convergence Via Data (in percent)

| age group (year) | 0 dose | 1 dose | 2 doses | 3 doses |
|---|---|---|---|---|
| 0 to 9 | 99.1 | 0.118 | 0.591 | 0.236 |
| 10 to 19 | 38.8 | 40.6 | 18.5 | 2.07 |
| 20 to 29 | 25.1 | 10.5 | 58.9 | 5.56 |
| 30 to 39 | 20.9 | 9.69 | 61.9 | 7.54 |
| 40 to 49 | 17.7 | 7.37 | 66.7 | 8.29 |
| 50 to 59 | 16.5 | 8.66 | 63.0 | 11.8 |
| 60 to 69 | 23.8 | 15.3 | 51.8 | 9.04 |
| $\geq$70 | 35.7 | 24.1 | 35.5 | 4.71 |

#### 3.1 VEFIAQR Simulation Results

##### 3.1.1 A 14-days Simulation

#### Contents


- steup simulation
- probability vector q (DBM and NGM are adopted respectively), for infection by a one-time contact
- simulate for each self-defined base-line R0


```
clear all; close all; clc;

ImportParametersVEFIAQR;   % import parameters omega, gamma, p, kappa,...
ImportFigureLegends; % import the following cell array for figure legend:
                     % groupLegend; ageLegend;
N = readmatrix('agePopulationVector.xlsx'); % the population vector (stratified by age)
C = readmatrix('contactMatrix.xlsx');   % the contact matrix
```

#### steup simulation

```
% initial value
% feasiblePoint = rand(n,4);
feasiblePoint = ones(n,4);
feasiblePoint = feasiblePoint ./ sum(feasiblePoint, 2);
xInit = [feasiblePoint .* N, zeros(n,24)];
xInit(4,13) = 1;        % first case in I_{4,0}

% time span
tInit = 0;
tFinal = 14;

% step size
stepSize = 0.01;

% a series R0 for simulation
R0 = [0.2, 0.5:6.5];     R0 = R0';
```

#### probability vector q (DBM and NGM are adopted respectively), for infection by a one-time contact

```
q_DBM = zeros(numel(R0),1);
q_NGM = zeros(numel(R0),1);

for i = 1:n
    for j = 1:n
        R(i,j) = C(j,i) / (dr(i) + p(i)*omegap(i) + (1-p(i))*omega(i)) * (kappa(i)*p(i)*omegap(i)/(gammap(i)+dr(i)) + (1-p(i))*omega(i)/(gamma(i)+dr(i)+f(i)));
    end
end


for i = 1:numel(R0)
    temp1 = R .* (N.') ./ sum(N);
    temp2 = sum(temp1,1);
    q_DBM(i) = R0(i) / sum(temp2);

    temp3 = 1 / (dr(i) + p(i)*omegap(i) + (1-p(i))*omega(i)) * (kappa(i)*p(i)*omegap(i)/(gammap(i)+dr(i)) + (1-p(i))*omega(i)/(gamma(i)+dr(i)+f(i)));
    q_NGM(i) = R0(i) / (max(eig(C)) * temp3);
end
fprintf('[R0, q_DBM, q_NGM] = \n');
disp([R0, q_DBM, q_NGM]);
```

```
[R0, q_DBM, q_NGM] =
   2.0000e-01   1.5693e-03   1.3569e-03
   5.0000e-01   3.9233e-03   3.3922e-03
   1.5000e+00   1.1770e-02   1.0177e-02
   2.5000e+00   1.9617e-02   1.6961e-02
   3.5000e+00   2.7463e-02   2.3746e-02
   4.5000e+00   3.5310e-02   3.0530e-02
   5.5000e+00   4.3157e-02   3.7314e-02
   6.5000e+00   5.1003e-02   4.4099e-02
```

#### simulate for each self-defined base-line R0

```
for i = 1:numel(R0)
    params.Beta   = C * q_DBM(i) ./ (N');
%     params.Beta   = C * q_NGM(i) ./ (N');

    fun = @(x,t)dxdt_VEFIAQR(x,t,params);
    [x,t] = odeSolveRK4(fun, tInit, xInit, tFinal, stepSize);
    [V, E, F, I, A, Q, R] = extractVariableFromCells_forVEFIAQR(x);


    % currentSymptomaticCases
    figure;
    I_by_doses = extractByVaccineDoses(I);
    plot(t,I_by_doses);
    legend(doseLegend);
    title(['current symptomatic cases \it{I}, with R_0 = ', num2str(R0(i))]);
    xlabel('time (in days)');
    ylabel('population size (in person)')

    % dailySymptomaticNewCases
    figure;
    dailySymptomaticNewCases_by_doses = extractByVaccineDoses((1-p(:)').*omegap(:)'.*F);
    plot(t,dailySymptomaticNewCases_by_doses);
    legend(doseLegend);
    title(['daily new symptomatic cases by doses, with R_0 = ', num2str(R0(i))]);
    xlabel('time (in days)');
    ylabel('population size (in person)')

    % allCurrentCases
    figure;
    allCurrentCases = extractByVaccineDoses(I+A);
    plot(t,allCurrentCases);
```

```
    legend(doseLegend);
    title(['all current cases \it{I + A}, with R_0 = ', num2str(R0(i))]);
    xlabel('time (in days)');
    ylabel('population size (in person)')

    % accumulativeNumberOfCases
    figure;
    dailyNewCases = extractByVaccineDoses(p(:)'.*omegapp(:)'.*F + (1-p(:)').*omegap(:)'.*F);
    accumulativeNumberOfCases = zeros(size(dailyNewCases,1), 4);
    for k = 1:4
        accumulativeNumberOfCases(:,k) = cumtrapz(t, dailyNewCases(:,k));
    end
    plot(t,accumulativeNumberOfCases);
    legend(doseLegend);
    title(['accumulative number of cases, with R_0 = ', num2str(R0(i))]);
    xlabel('time (in days)');
    ylabel('population size (in person)');


    % how to define and compute the accumulativeNumberOfCases from model directly
    % instead of a extra numerical integration, it is still an very interesting question.
    Nt = sum(V + E + F + I + A + Q + R) / sum(N);
end
```

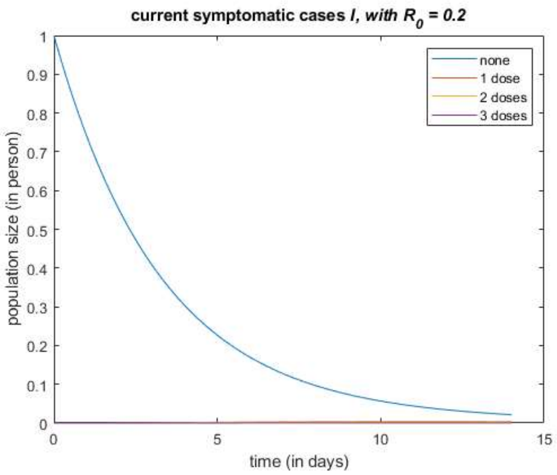


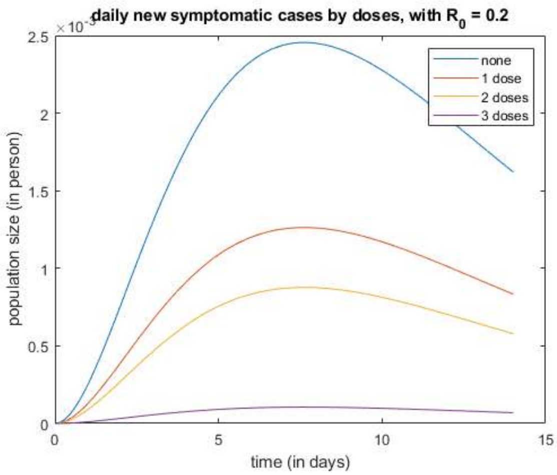


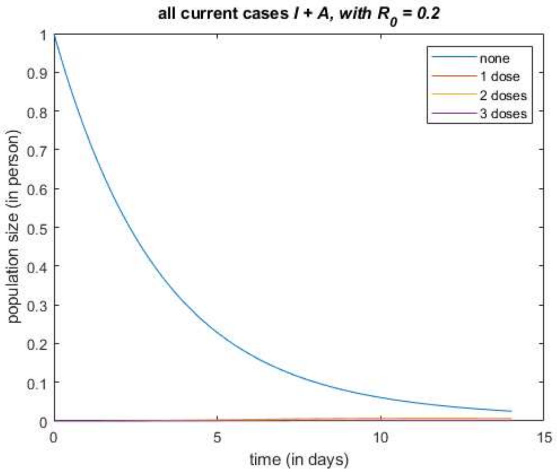


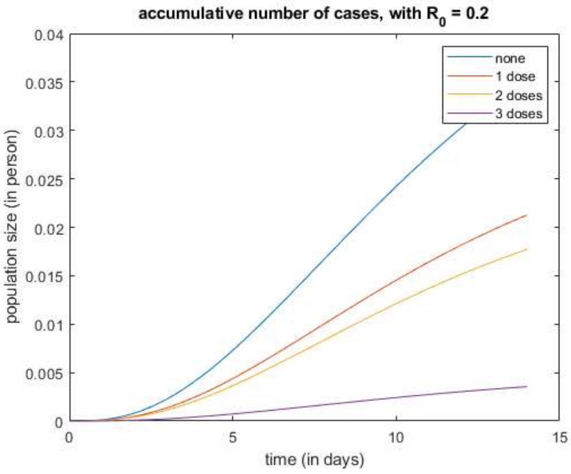


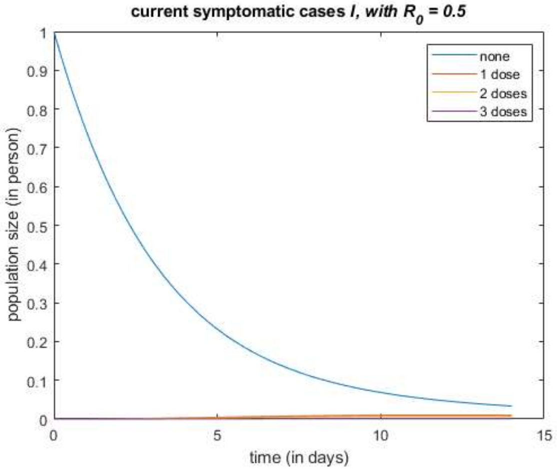


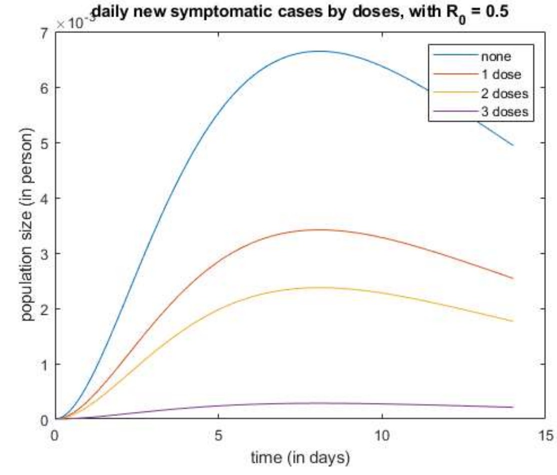

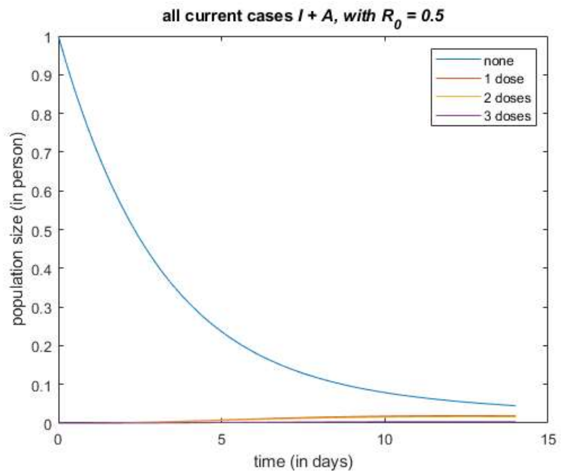
all current cases I + A, with R0 = 0.5
population size (in person)
time (in days)
none
1 dose
2 doses
3 doses

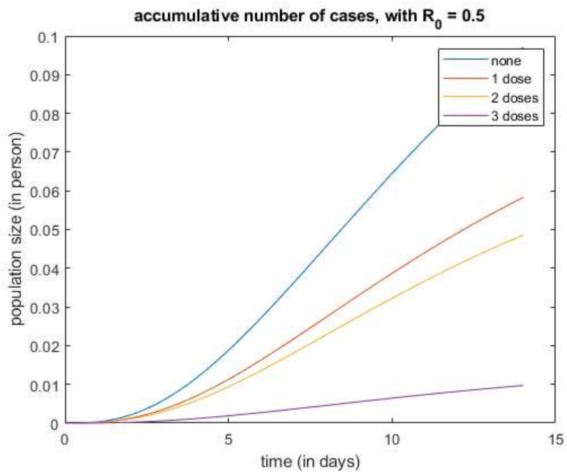
accumulative number of cases, with R0 = 0.5
population size (in person)
time (in days)
none
1 dose
2 doses
3 doses

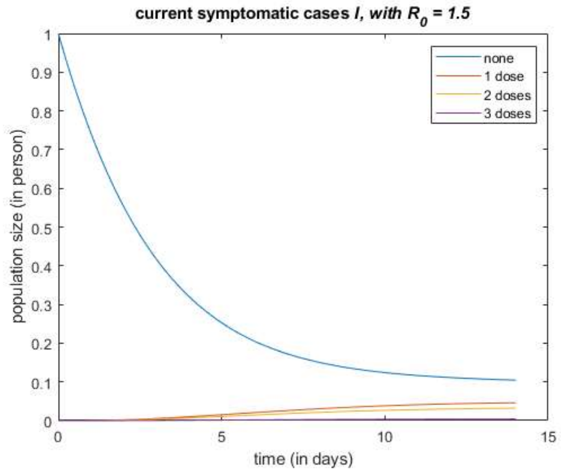
current symptomatic cases I, with R0 = 1.5
population size (in person)
time (in days)
none
1 dose
2 doses
3 doses

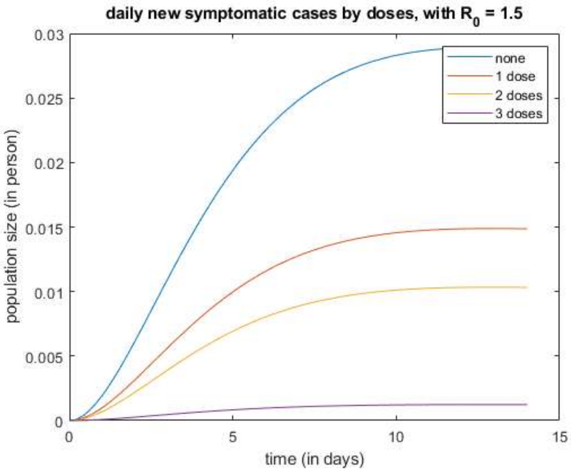
daily new symptomatic cases by doses, with R0 = 1.5
population size (in person)
time (in days)
none
1 dose
2 doses
3 doses

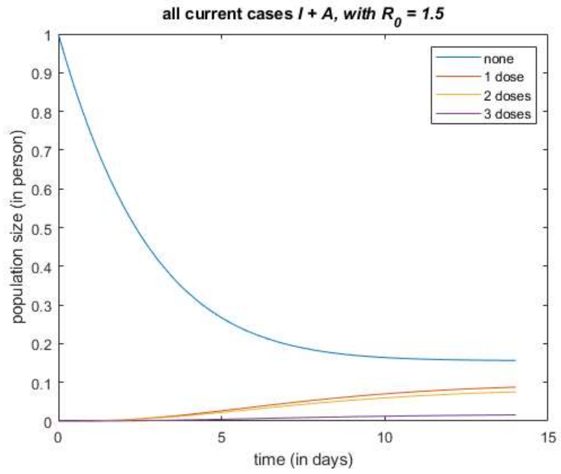
all current cases I + A, with R0 = 1.5
population size (in person)
time (in days)
none
1 dose
2 doses
3 doses

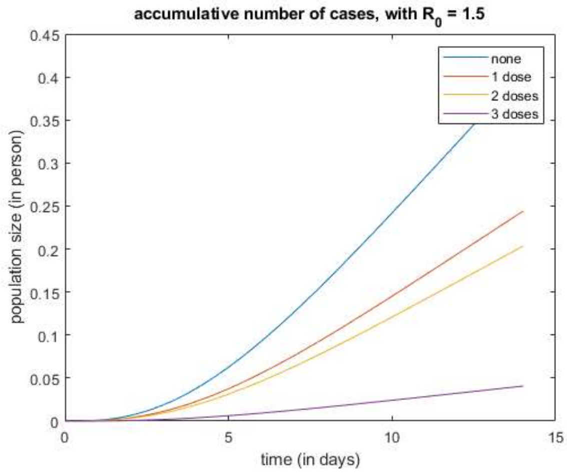
accumulative number of cases, with R0 = 1.5
population size (in person)
time (in days)
none
1 dose
2 doses
3 doses

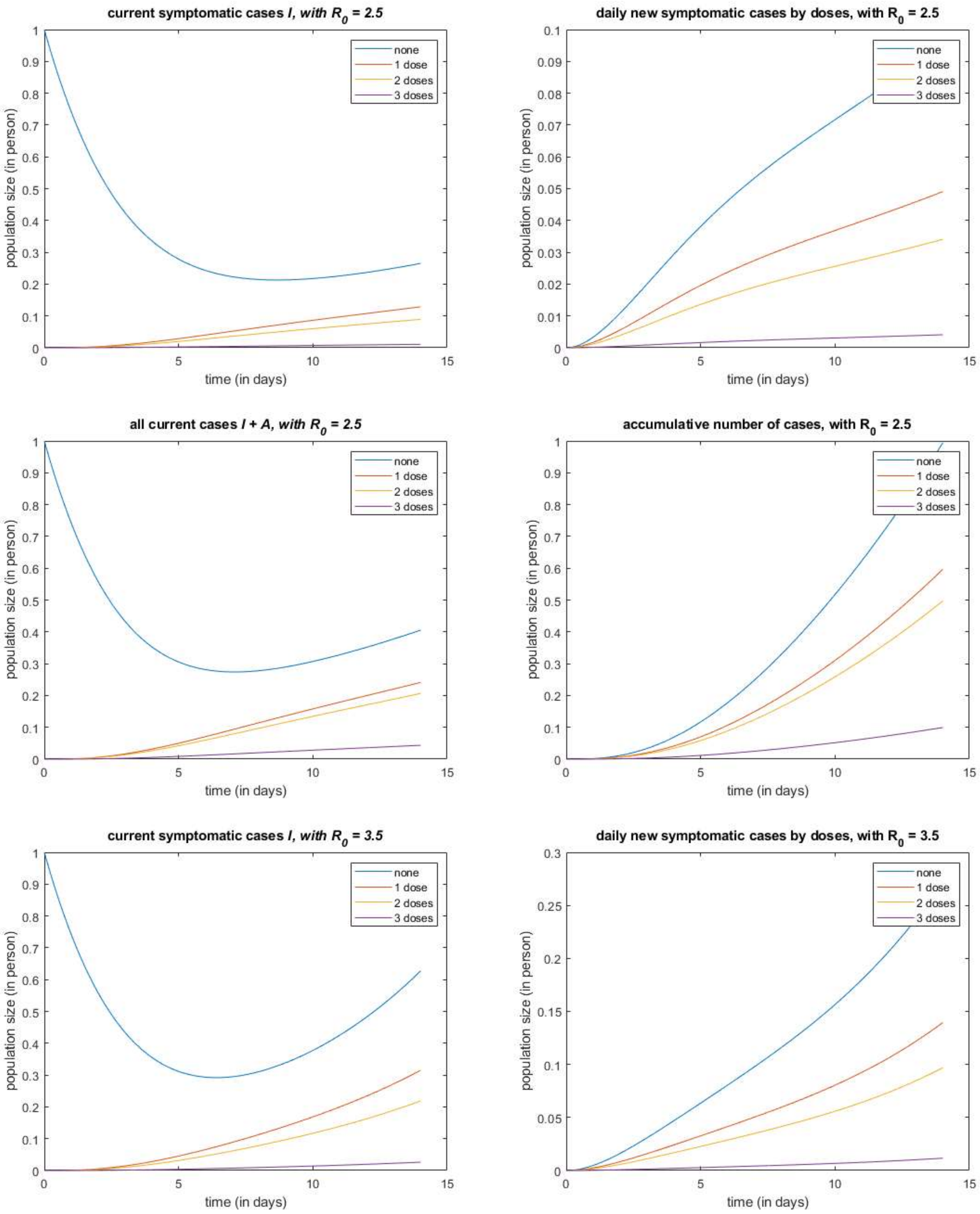
current symptomatic cases I, with R0 = 2.5
daily new symptomatic cases by doses, with R0 = 2.5
all current cases I + A, with R0 = 2.5
accumulative number of cases, with R0 = 2.5
current symptomatic cases I, with R0 = 3.5
daily new symptomatic cases by doses, with R0 = 3.5
population size (in person)
time (in days)
none
1 dose
2 doses
3 doses

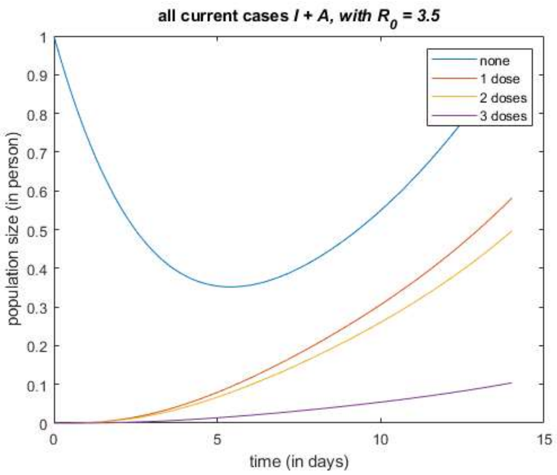
all current cases I + A, with $R_0$ = 3.5
population size (in person)
time (in days)
none
1 dose
2 doses
3 doses

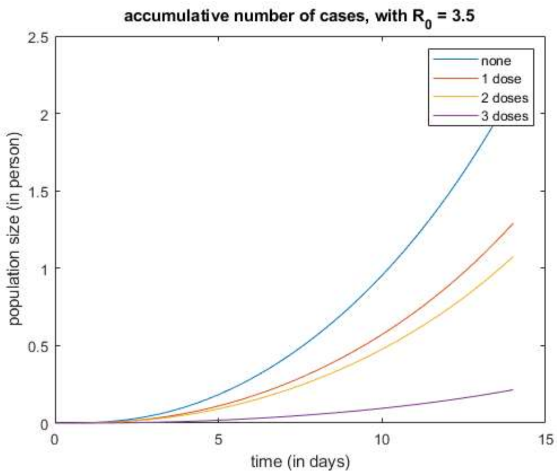
accumulative number of cases, with $R_0$ = 3.5
population size (in person)
time (in days)
none
1 dose
2 doses
3 doses

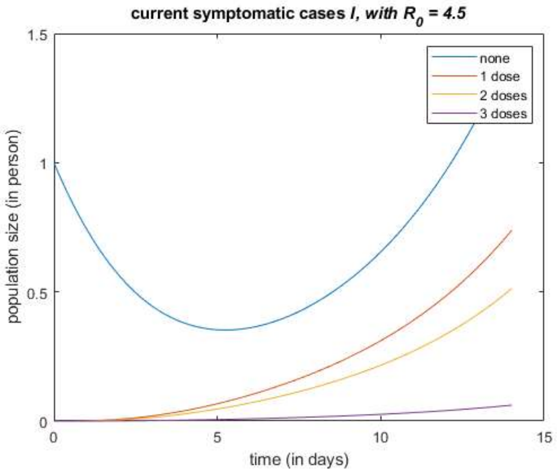
current symptomatic cases I, with $R_0$ = 4.5
population size (in person)
time (in days)
none
1 dose
2 doses
3 doses

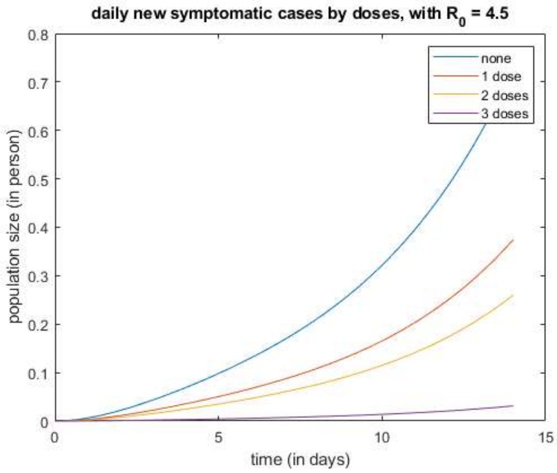
daily new symptomatic cases by doses, with $R_0$ = 4.5
population size (in person)
time (in days)
none
1 dose
2 doses
3 doses

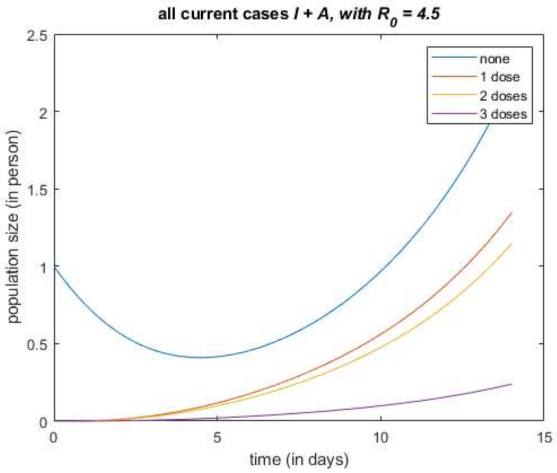
all current cases I + A, with $R_0$ = 4.5
population size (in person)
time (in days)
none
1 dose
2 doses
3 doses

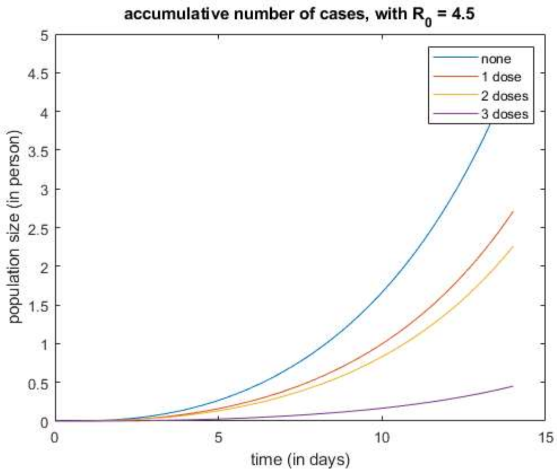
accumulative number of cases, with $R_0$ = 4.5
population size (in person)
time (in days)
none
1 dose
2 doses
3 doses

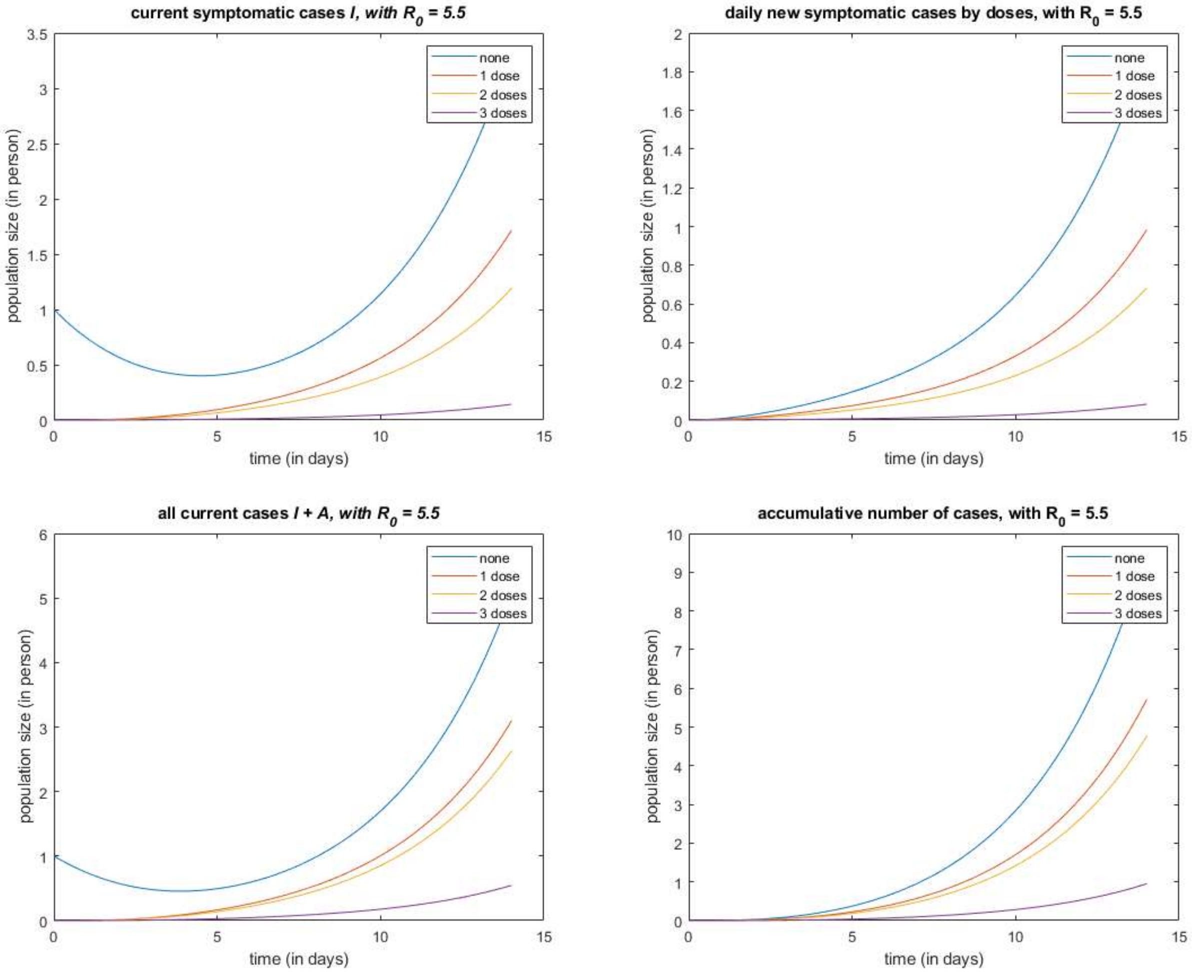
current symptomatic cases I, with $R_0$ = 5.5
daily new symptomatic cases by doses, with $R_0$ = 5.5
all current cases I + A, with $R_0$ = 5.5
accumulative number of cases, with $R_0$ = 5.5
population size (in person)
time (in days)
none
1 dose
2 doses
3 doses

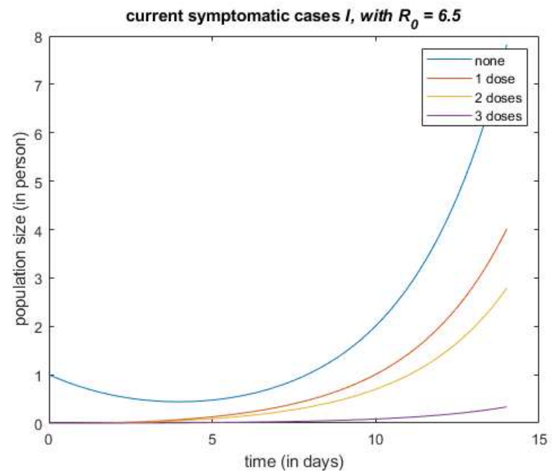


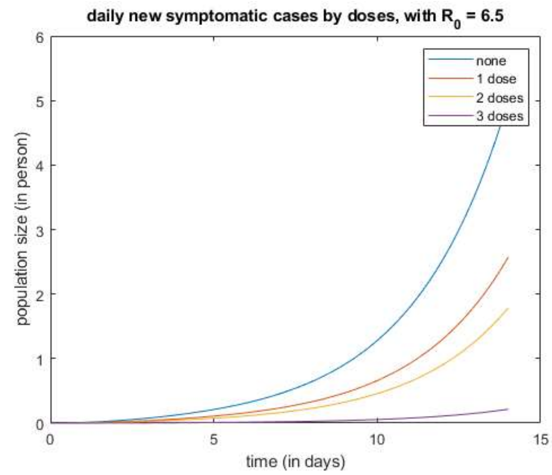


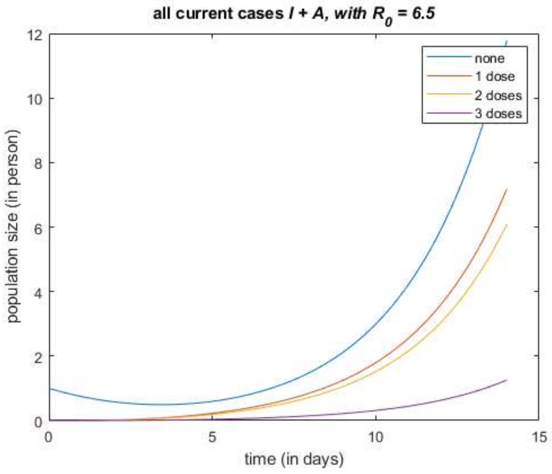


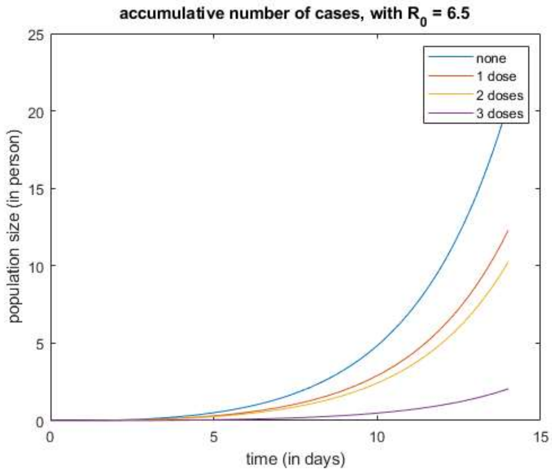


*Published with MATLAB® R2021a*

##### 3.1.2 A 100-days Simulation

#### Contents


- steup simulation
- probability vector q (DBM and NGM are adopted respectively), for infection by a one-time contact
- simulate for each self-defined base-line R0


```
clear all; close all; clc;

ImportParametersVEFIAQR;   % import parameters omega, gamma, p, kappa,...
ImportFigureLegends; % import the following cell array for figure legend:
                     % groupLegend; ageLegend;
N = readmatrix('agePopulationVector.xlsx'); % the population vector (stratified by age)
C = readmatrix('contactMatrix.xlsx');   % the contact matrix
```

#### steup simulation

```
% initial value
% feasiblePoint = rand(n,4);
feasiblePoint = ones(n,4);
feasiblePoint = feasiblePoint ./ sum(feasiblePoint, 2);
xInit = [feasiblePoint .* N, zeros(n,24)];
xInit(4,13) = 1;        % first case in I_{4,0}

% time span
tInit = 0;
tFinal = 100;

% step size
stepSize = 0.01;

% a series R0 for simulation
R0 = [0.2, 0.5:6.5];     R0 = R0';
```

#### probability vector q (DBM and NGM are adopted respectively), for infection by a one-time contact

```
q_DBM = zeros(numel(R0),1);
q_NGM = zeros(numel(R0),1);

for i = 1:n
    for j = 1:n
        R(i,j) = C(j,i) / (dr(i) + p(i)*omegap(i) + (1-p(i))*omega(i)) * (kappa(i)*p(i)*omegap(i)/(gammap(i)+dr(i)) + (1-p(i))*omega(i)/(gamma(i)+dr(i)+f(i)));
    end
end


for i = 1:numel(R0)
    temp1 = R .* (N.') ./ sum(N);
    temp2 = sum(temp1,1);
    q_DBM(i) = R0(i) / sum(temp2);

    temp3 = 1 / (dr(i) + p(i)*omegap(i) + (1-p(i))*omega(i)) * (kappa(i)*p(i)*omegap(i)/(gammap(i)+dr(i)) + (1-p(i))*omega(i)/(gamma(i)+dr(i)+f(i)));
    q_NGM(i) = R0(i) / (max(eig(C)) * temp3);
end
fprintf('[R0, q_DBM, q_NGM] = \n');
disp([R0, q_DBM, q_NGM]);
```

```
[R0, q_DBM, q_NGM] =
   2.0000e-01   1.5693e-03   1.3569e-03
   5.0000e-01   3.9233e-03   3.3922e-03
   1.5000e+00   1.1770e-02   1.0177e-02
   2.5000e+00   1.9617e-02   1.6961e-02
   3.5000e+00   2.7463e-02   2.3746e-02
   4.5000e+00   3.5310e-02   3.0530e-02
   5.5000e+00   4.3157e-02   3.7314e-02
   6.5000e+00   5.1003e-02   4.4099e-02
```

#### simulate for each self-defined base-line R0

```
for i = 1:numel(R0)
    params.Beta   = C * q_DBM(i) ./ (N');
%     params.Beta   = C * q_NGM(i) ./ (N');

    fun = @(x,t)dxdt_VEFIAQR(x,t,params);
    [x,t] = odeSolveRK4(fun, tInit, xInit, tFinal, stepSize);
    [V, E, F, I, A, Q, R] = extractVariableFromCells_forVEFIAQR(x);


    % currentSymptomaticCases
    figure;
    I_by_doses = extractByVaccineDoses(I);
    plot(t,I_by_doses);
    legend(doseLegend);
    title(['current symptomatic cases \it{I}, with R_0 = ', num2str(R0(i))]);
    xlabel('time (in days)');
    ylabel('population size (in person)')

    % dailySymptomaticNewCases
    figure;
    dailySymptomaticNewCases_by_doses = extractByVaccineDoses((1-p(:)').*omegap(:)'.*F);
    plot(t,dailySymptomaticNewCases_by_doses);
    legend(doseLegend);
    title(['daily new symptomatic cases by doses, with R_0 = ', num2str(R0(i))]);
    xlabel('time (in days)');
    ylabel('population size (in person)')

    % allCurrentCases
    figure;
    allCurrentCases = extractByVaccineDoses(I+A);
    plot(t,allCurrentCases);
```

```
    legend(doseLegend);
    title(['all current cases \it{I + A}, with R_0 = ', num2str(R0(i))]);
    xlabel('time (in days)');
    ylabel('population size (in person)')

    % accumulativeNumberOfCases
    figure;
    dailyNewCases = extractByVaccineDoses(p(:)'.*omegapp(:)'.*F + (1-p(:)').*omegap(:)'.*F);
    accumulativeNumberOfCases = zeros(size(dailyNewCases,1), 4);
    for k = 1:4
        accumulativeNumberOfCases(:,k) = cumtrapz(t, dailyNewCases(:,k));
    end
    plot(t,accumulativeNumberOfCases);
    legend(doseLegend);
    title(['accumulative number of cases, with R_0 = ', num2str(R0(i))]);
    xlabel('time (in days)');
    ylabel('population size (in person)');


    % how to define and compute the accumulativeNumberOfCases from model directly
    % instead of a extra numerical integration, it is still an very interesting question.
    Nt = sum(V + E + F + I + A + Q + R) / sum(N);
end
```

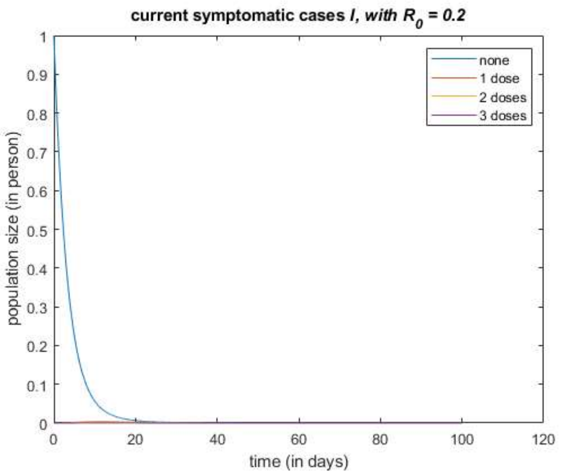


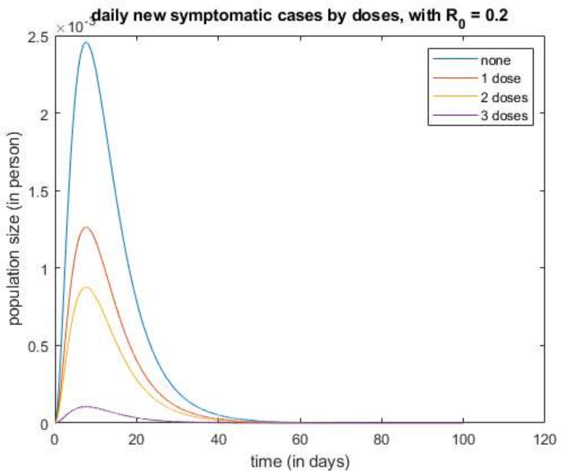


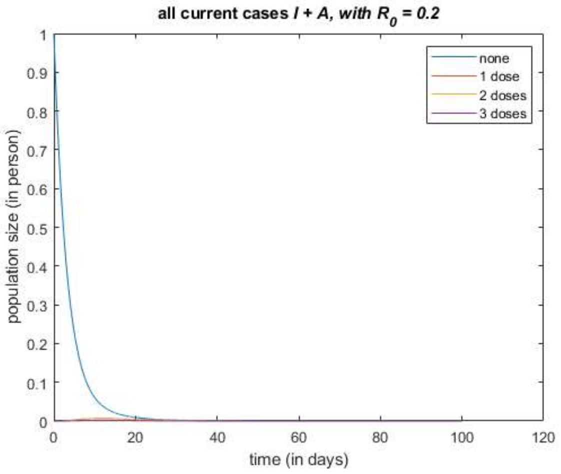


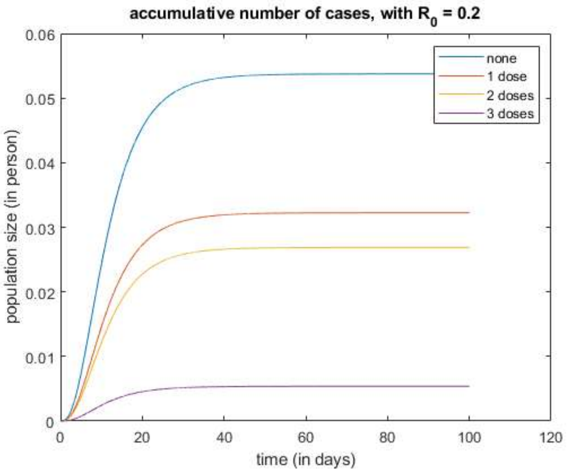


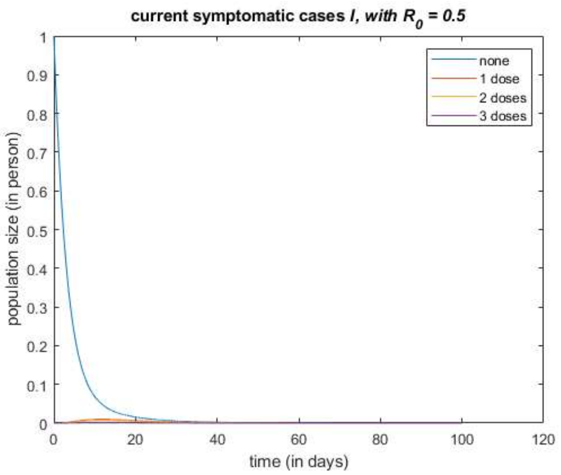


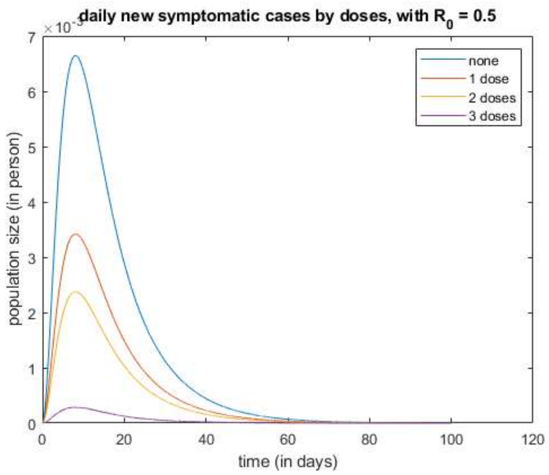

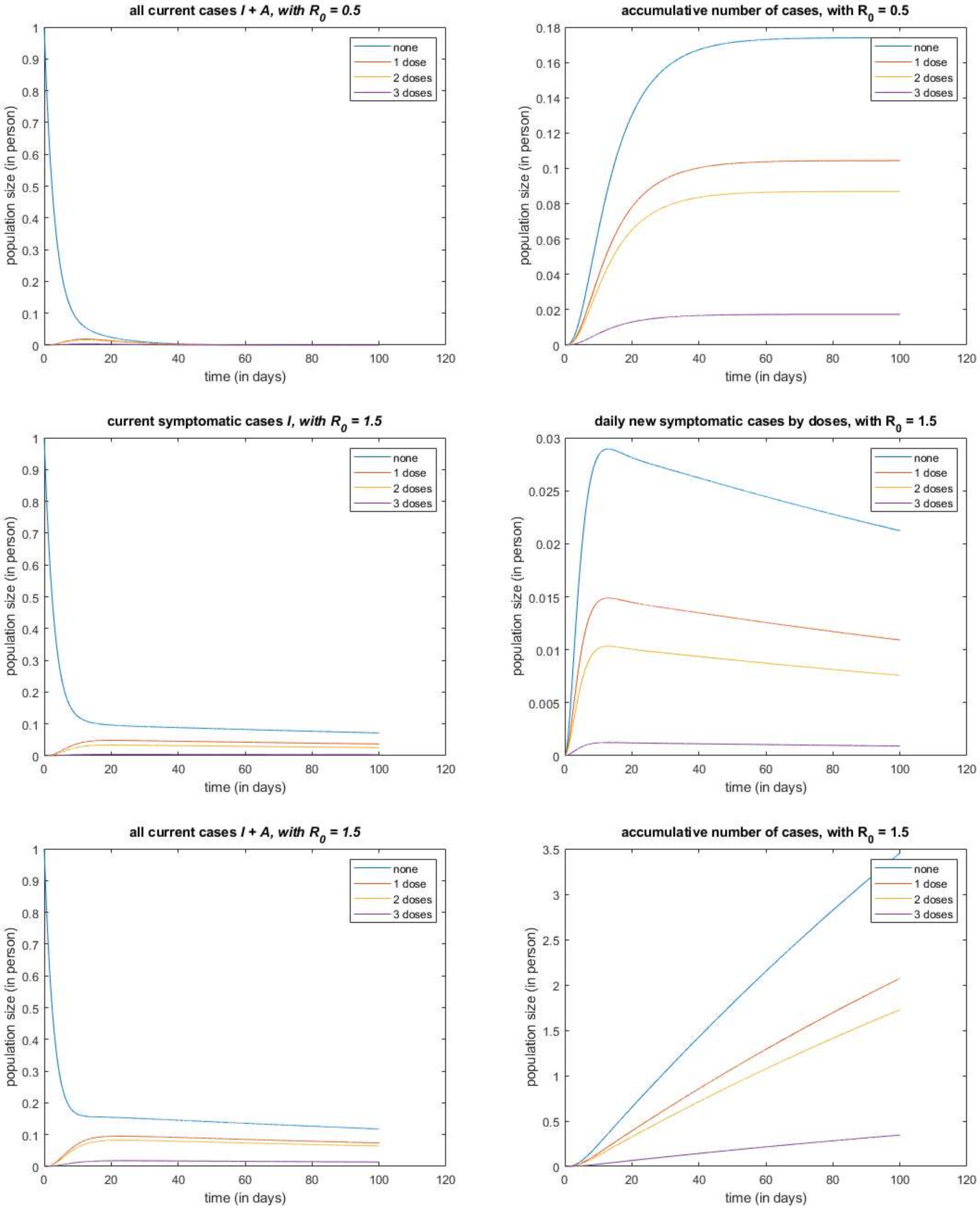
all current cases I + A, with R0 = 0.5
accumulative number of cases, with R0 = 0.5
current symptomatic cases I, with R0 = 1.5
daily new symptomatic cases by doses, with R0 = 1.5
all current cases I + A, with R0 = 1.5
accumulative number of cases, with R0 = 1.5
none
1 dose
2 doses
3 doses
population size (in person)
time (in days)

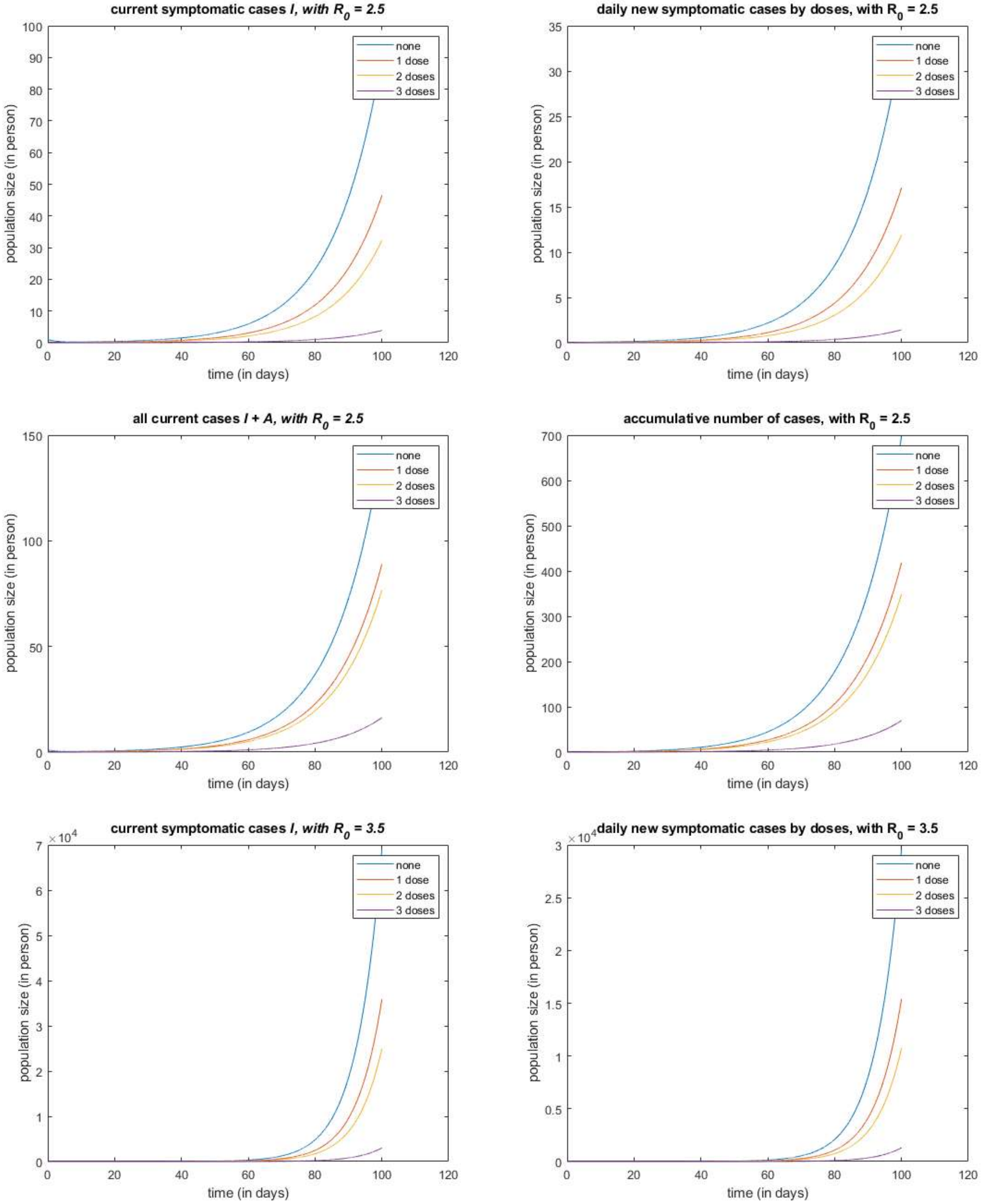
current symptomatic cases I, with R0 = 2.5
daily new symptomatic cases by doses, with R0 = 2.5
all current cases I + A, with R0 = 2.5
accumulative number of cases, with R0 = 2.5
current symptomatic cases I, with R0 = 3.5
daily new symptomatic cases by doses, with R0 = 3.5
population size (in person)
time (in days)
none
1 dose
2 doses
3 doses
×10^4

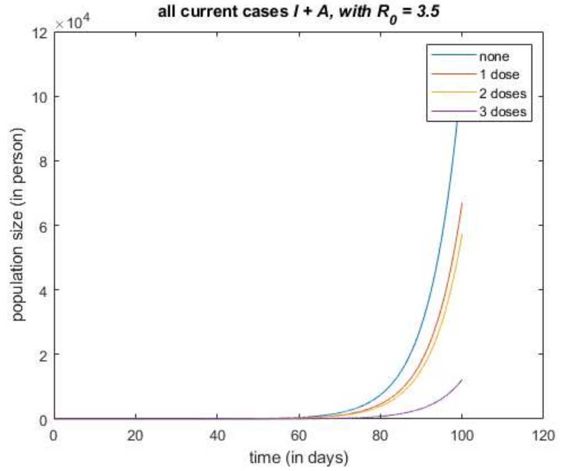
all current cases I + A, with R0 = 3.5
×10^4
population size (in person)
time (in days)
none
1 dose
2 doses
3 doses

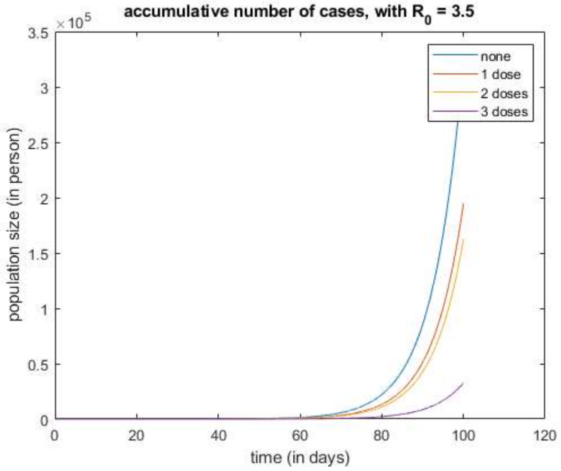
accumulative number of cases, with R0 = 3.5
×10^5
population size (in person)
time (in days)
none
1 dose
2 doses
3 doses

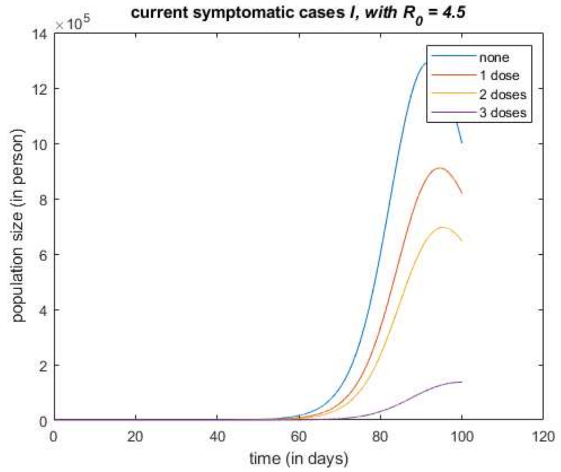
current symptomatic cases I, with R0 = 4.5
×10^5
population size (in person)
time (in days)
none
1 dose
2 doses
3 doses

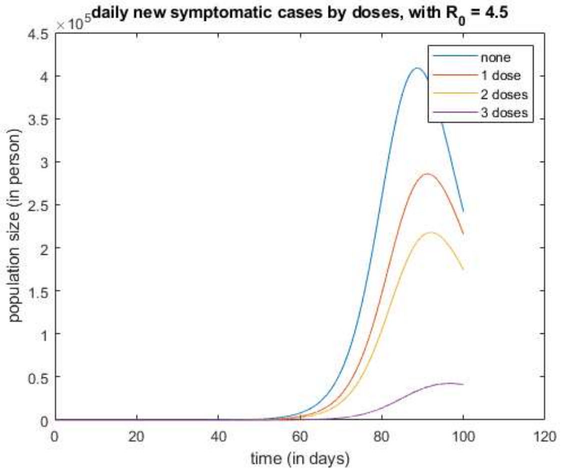
daily new symptomatic cases by doses, with R0 = 4.5
×10^5
population size (in person)
time (in days)
none
1 dose
2 doses
3 doses

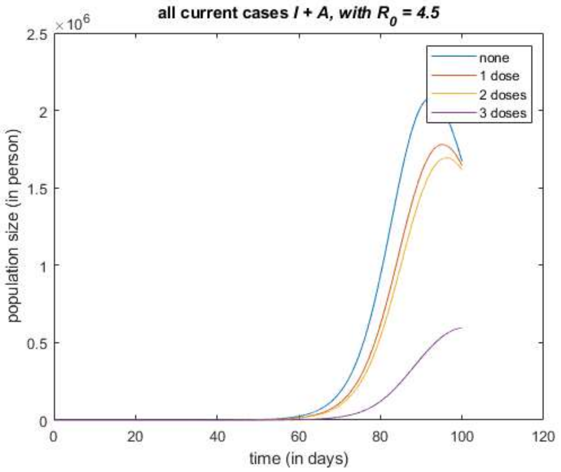
all current cases I + A, with R0 = 4.5
×10^6
population size (in person)
time (in days)
none
1 dose
2 doses
3 doses

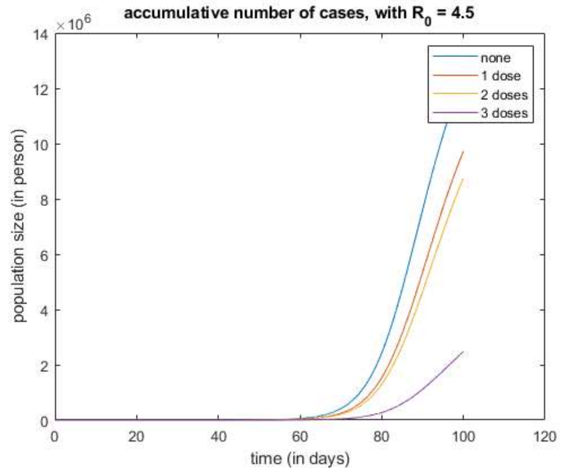
accumulative number of cases, with R0 = 4.5
×10^6
population size (in person)
time (in days)
none
1 dose
2 doses
3 doses

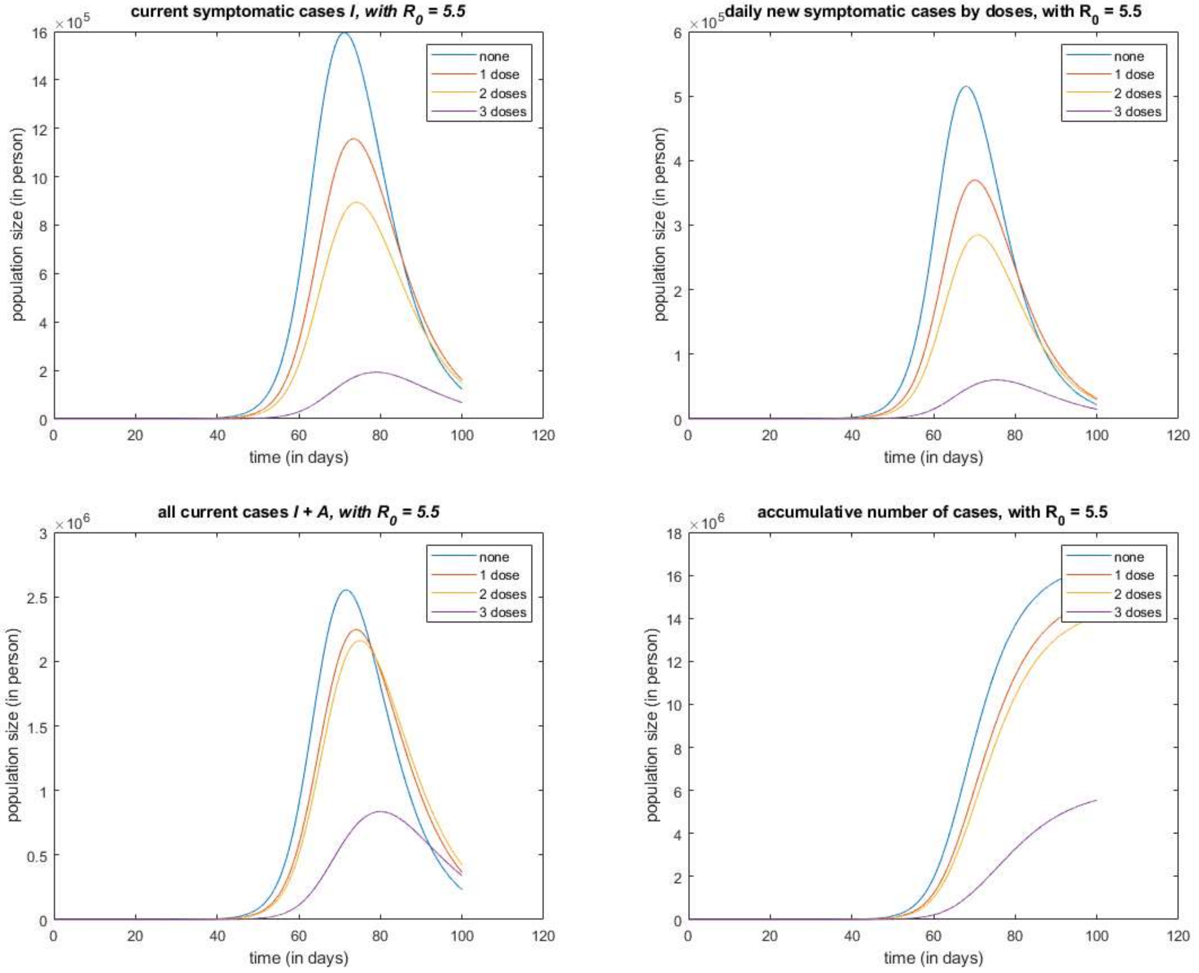
current symptomatic cases I, with R0 = 5.5
daily new symptomatic cases by doses, with R0 = 5.5
all current cases I + A, with R0 = 5.5
accumulative number of cases, with R0 = 5.5
population size (in person)
time (in days)
none
1 dose
2 doses
3 doses
×10^5
×10^6

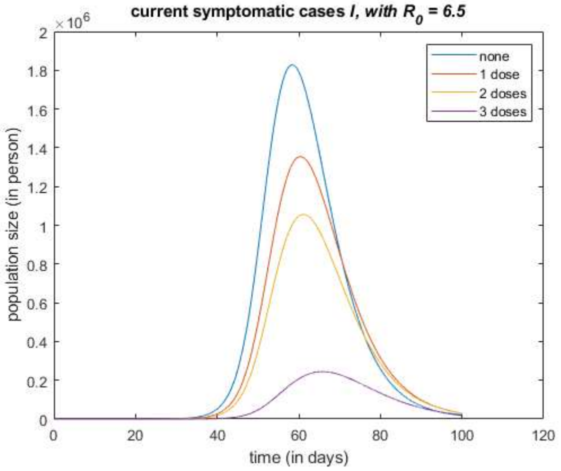


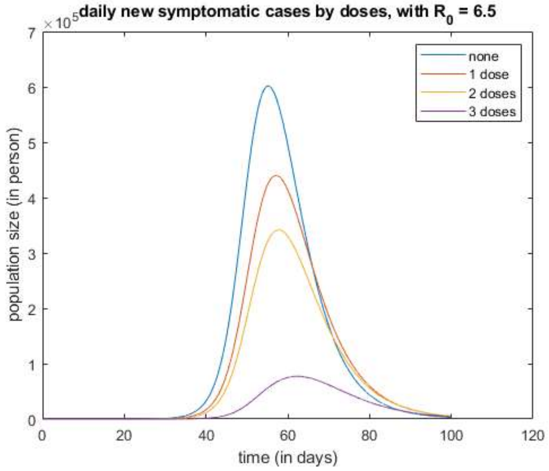


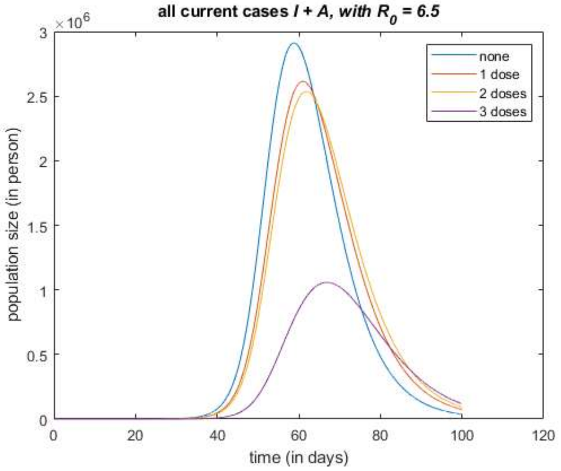


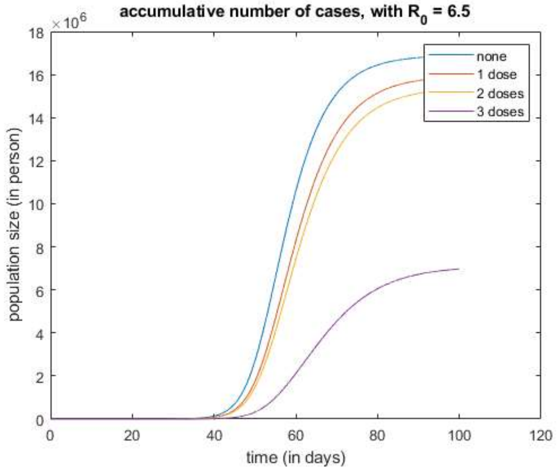


*Published with MATLAB® R2021a*

### 4 Vaccination Optimizing

There are two interesting questions:

- If one has 1 dose vaccine available, then who should got this dose?
- If there are 100 million doses available, then how shall we distribute them? (to each age group and to those already vaccinated 0, 1, or 2 doses)

To answer those two questions, we define the following optimizing problem, and use the directional derivative as a criterion for vaccine distributing.

#### 4.1 The Optimizing Problem

All accumulative cases in a given time period is no doubt a important index for effectiveness of local disease control. In the construction of the objective function, we may weight the cost of each cases by its severity rankings (asymptomatic, mild, severe, ICU, death).

**Objective Function**:

$$f = \sum \text{Weighted Costs of all accumulative cases in a given time period} \tag{50}$$

$$= \sum_{i=1}^{n} \sum_{j=0}^{3} \int_0^T g(E_{ij}, I_{ij}, A_{ij}, Q_{ij}, parameters) \tag{51}$$

where $g(\cdot)$ denote the weight function; $i$ denote the $i$-th age group; $j$ denote group of those finished $j$-th doses; $E_{ij}$, $I_{ij}$, $A_{ij}$, $Q_{ij}$ denotes the solution of the systems of ODEs; parameters denotes the constant parameters in the systems of ODEs.

The objective function $f$ is implicit since there is no explicit solutions of $E_{ij}$, $I_{ij}$, $A_{ij}$, $Q_{ij}$. Therefore, as a good approximation of the original objective function $f$, a numerical solution is adopted with a given method and a given step size choosing method. That is, we use the following objective function $\tilde{f}$ instead:

$$\tilde{f} = \sum_{i=1}^{n} \sum_{j=0}^{3} \int_0^T g(\tilde{E}_{ij}, \tilde{I}_{ij}, \tilde{A}_{ij}, \tilde{Q}_{ij}, parameters, method, stepSize) \tag{52}$$

here $\int$ denotes the numerical integration; $\tilde{E}_{ij}, \tilde{I}_{ij}, \tilde{A}_{ij}, \tilde{Q}_{ij}$ denotes the numerical solution of the systems of ODEs using given numerical method and stepSize choosing method. For the sake of convenience, we use symbol $f$ represent $\tilde{f}$ in following sections.

Note: We treated those compartment as continuous variable, and use 4-order Runge-Kuatt method with fixed step size to perform numerical integrals to construct the objective function. The use of numerical integration adds no difficulties, but makes the objective function more preciser than [1], where a simply summation of daily values obtained via Euler Method with 1-day step size are adopted.

**Decision Variable**:

$$nVC = \begin{pmatrix} \mathrm{nVC}_{10} & \mathrm{nVC}_{11} & \mathrm{nVC}_{12} & \mathrm{nVC}_{13} \\ \mathrm{nVC}_{20} & \mathrm{nVC}_{21} & \mathrm{nVC}_{22} & \mathrm{nVC}_{23} \\ \mathrm{nVC}_{30} & \mathrm{nVC}_{31} & \mathrm{nVC}_{32} & \mathrm{nVC}_{33} \\ \mathrm{nVC}_{40} & \mathrm{nVC}_{41} & \mathrm{nVC}_{42} & \mathrm{nVC}_{43} \\ \mathrm{nVC}_{50} & \mathrm{nVC}_{51} & \mathrm{nVC}_{52} & \mathrm{nVC}_{53} \\ \mathrm{nVC}_{60} & \mathrm{nVC}_{61} & \mathrm{nVC}_{62} & \mathrm{nVC}_{63} \\ \mathrm{nVC}_{70} & \mathrm{nVC}_{71} & \mathrm{nVC}_{72} & \mathrm{nVC}_{73} \\ \mathrm{nVC}_{80} & \mathrm{nVC}_{81} & \mathrm{nVC}_{82} & \mathrm{nVC}_{83} \end{pmatrix} \tag{53}$$

where $nVC_{ij}$ denotes the population size of group $ij$, which is formed by those in age group $i$ and already finished $j$ doses.

Note that $f$ is a function of $nVC$, that is, $f(nVC)$.

**Feasible Set**:

We may normalize the vaccine convergence $nVC$ using population size of each age groups, and obtain a vaccine convergence rate $VC$.

The feasible set is a set of all point $VC$ that is feasible. Those points satisfies: 1) each entry of $VC$ is non-negative, 2) summation of each row of $VC$ equals to 1 (for $nVC$, this summation equals to the population size of the corresponding age group). Such feasible set is indeed a probabilistic simplex with linear constrains (i.e. intersection of a probabilistic simplex and a hyperplane). Therefore, one may consider the optimization on manifolds, use the Riemann Gradient instead of Euclidean Gradient for updating, or to consider the alternative projection between the simplex and the hyperplane.

Unfortunately, the objective function $f(VC)$ is unavailable in this problem, and so it is the gradient. This is because $f$ is defined via numerical solution of systems of ODEs with 1) a given step size choosing method and 2) a given update method and 3) other parameters.

#### 4.2 Directional Derivatives of all possible update direction

This optimizing is a difficult problem, since only the computation of the objective function itself is available, without the gradient or hessian or any other information that how $f$ behaves on the feasible set. And one may consider the Intelligent Optimization Algorithms (Genetic Algorithm, Simulated annealing algorithm, Evolutionary computation, Ant colony optimization algorithms, Immune Algorithm, Tabu Search Algorithm), Nelder-Mead simplex algorithm, and other algorithms that only computations of objective function are required.

Unlike those global search on the entire feasible set, we found that the information of gradient is sufficient to answer the question "Who to vaccinate first?", and use numerical directional derivatives (the action of gradient on specific doses assignments) as criteria for choosing a direction for update.

Assume that the current vaccine convergence is describe by $nVC$, then for each newly vaccinated dose, the convergence $nVC$ is added with a one-dose perturbation $\Delta VC_{ij}$, a $8 \times 4$ matrix with its $(i, j)$-th entry equals to $-1$, and $(i, j+1)$ entry equals to 1, and otherwise 0. Those $\Delta VC_{ij}$ constitute all possible direction for updating.

For example, when one individual of age group 7 who has already finished 1 dose is further vaccinated 1 dose, then the following perturbation is added to the current vaccine convergence $nVC$:

$$\Delta VC_{71} = \begin{pmatrix} 0 & 0 & 0 & 0 \\ 0 & 0 & 0 & 0 \\ 0 & 0 & 0 & 0 \\ 0 & 0 & 0 & 0 \\ 0 & 0 & 0 & 0 \\ 0 & 0 & 0 & 0 \\ 0 & -1 & 1 & 0 \\ 0 & 0 & 0 & 0 \end{pmatrix} \tag{54}$$

The differences of objective function:

$$\Delta f_{ij} = f(VC) - f(VC + \Delta VC_{ij}) \tag{55}$$

represent the effectiveness of this dose distributed to a individual of age group $i$ who has already finished $j$ doses. Hence, by computing the maximum $\Delta f_{ij}$ in each step, a series of vaccinating decision is obtained.

##### 4.3 Greedy Algorithm for Optimization of Vaccinate Process

###### 4.3.1 Pseudo-Code

**Algorithm 1** Greedy Alogrithm for Vaccinating Process

**Input:** Objective function $f(nVC)$ for minimization; Current vaccine convergence $nVC$; Number of available doses $m$; Parameters of the VEFIAQR dynamical system; Days for simulation.

**Output:** A decision matrix $\Delta nVC$ indicating best order of vaccinating process (in the sense of minimizing accumulative cases) (rows of $\Delta nVC$ indicates decision of each doses, and columns of $\Delta nVC$ indicate different decisions for updating)

```
for i = 1 to m do
    for all possible decision ΔnVC of the i-th dose distributing do
        Compute and save Δf = f(nVC) − f(nVC + ΔnVC), the decrease of the specific decision
    end for
    Find and save the decision which produce maximum ΔnVC_i
    Update the current convergence nVC by nVC = nVC + ΔnVC_i
end for
Output decision matrix ΔnVC
```

### 5 Non-pharmacological interventions (NPIs)

#### 5.1 Contact Matrix Decomposition

We categorize the contact data into following categories:

- Public Transports
- Amusement
- School
- Household
- Hospital
- Working Area
- Service Industry

- Community
- Catering industry
- Uncategorized

And for each category, we trained the corresponding contact matrix via the maximum likelihood estimation proposed in Chapter 1. Those matrices are given as follows:

```
clear all; close all; clc;
contactMatrixDecomposition;
```

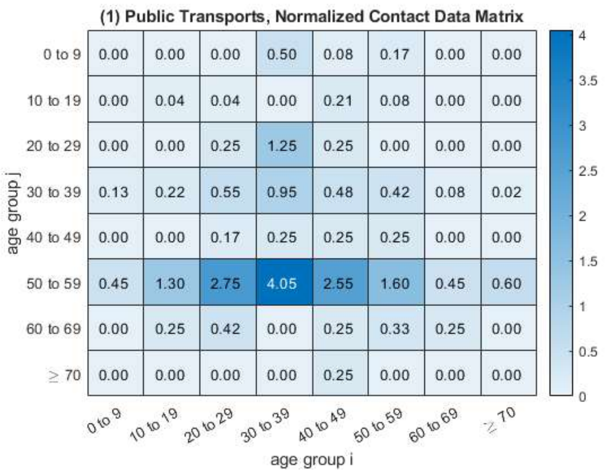


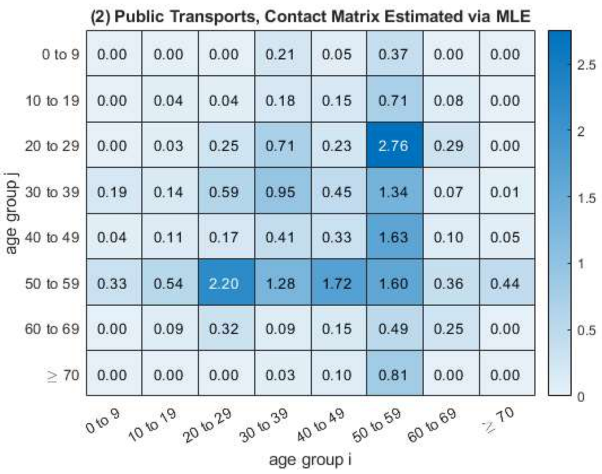


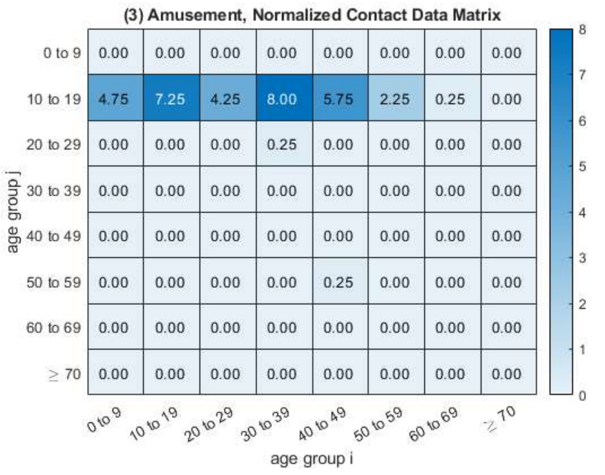


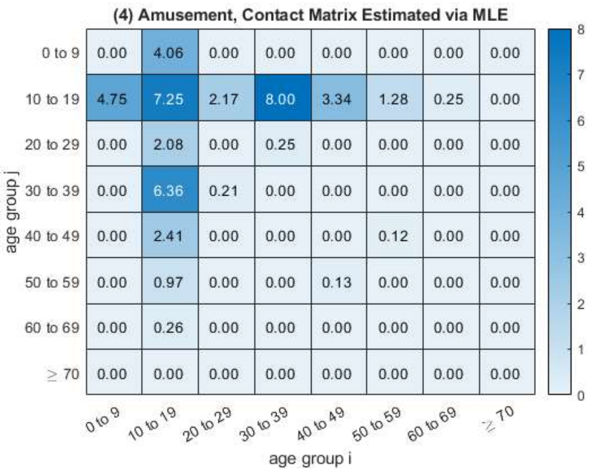


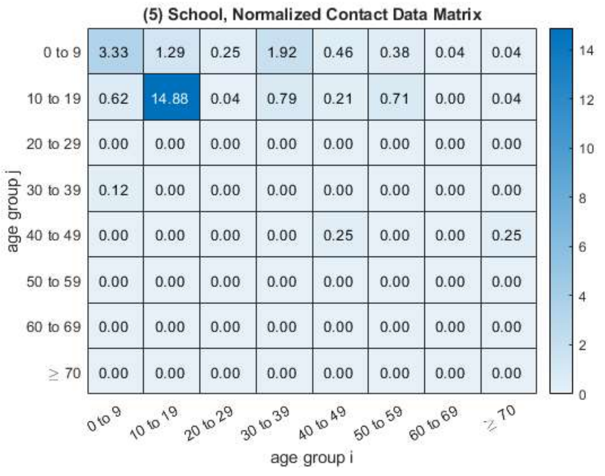


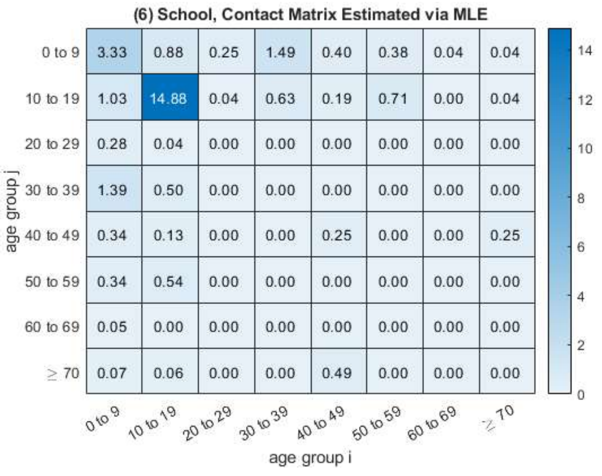

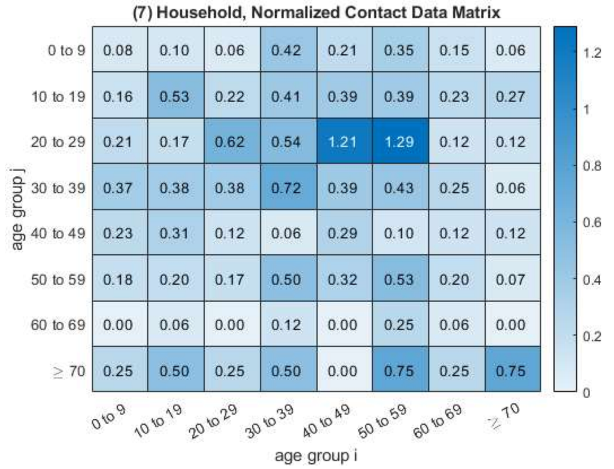
(7) Household, Normalized Contact Data Matrix
0 to 9 0.08 0.10 0.06 0.42 0.21 0.35 0.15 0.06
10 to 19 0.16 0.53 0.22 0.41 0.39 0.39 0.23 0.27
20 to 29 0.21 0.17 0.62 0.54 1.21 1.29 0.12 0.12
30 to 39 0.37 0.38 0.38 0.72 0.39 0.43 0.25 0.06
40 to 49 0.23 0.31 0.12 0.06 0.29 0.10 0.12 0.12
50 to 59 0.18 0.20 0.17 0.50 0.32 0.53 0.20 0.07
60 to 69 0.00 0.06 0.00 0.12 0.00 0.25 0.06 0.00
≥ 70 0.25 0.50 0.25 0.50 0.00 0.75 0.25 0.75
age group j
0 to 9 10 to 19 20 to 29 30 to 39 40 to 49 50 to 59 60 to 69 ≥ 70
age group i

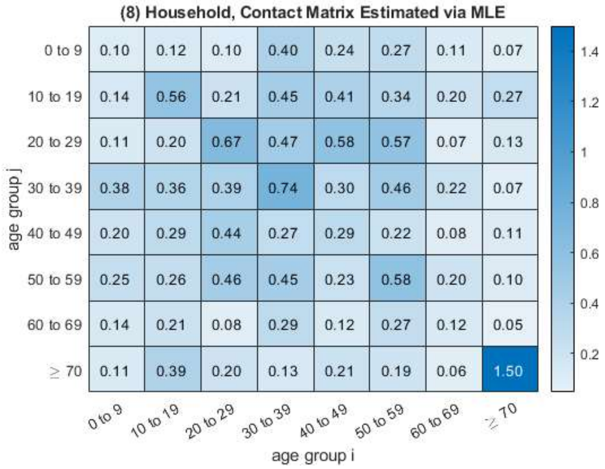
(8) Household, Contact Matrix Estimated via MLE
0 to 9 0.10 0.12 0.10 0.40 0.24 0.27 0.11 0.07
10 to 19 0.14 0.56 0.21 0.45 0.41 0.34 0.20 0.27
20 to 29 0.11 0.20 0.67 0.47 0.58 0.57 0.07 0.13
30 to 39 0.38 0.36 0.39 0.74 0.30 0.46 0.22 0.07
40 to 49 0.20 0.29 0.44 0.27 0.29 0.22 0.08 0.11
50 to 59 0.25 0.26 0.46 0.45 0.23 0.58 0.20 0.10
60 to 69 0.14 0.21 0.08 0.29 0.12 0.27 0.12 0.05
≥ 70 0.11 0.39 0.20 0.13 0.21 0.19 0.06 1.50
age group j
0 to 9 10 to 19 20 to 29 30 to 39 40 to 49 50 to 59 60 to 69 ≥ 70
age group i

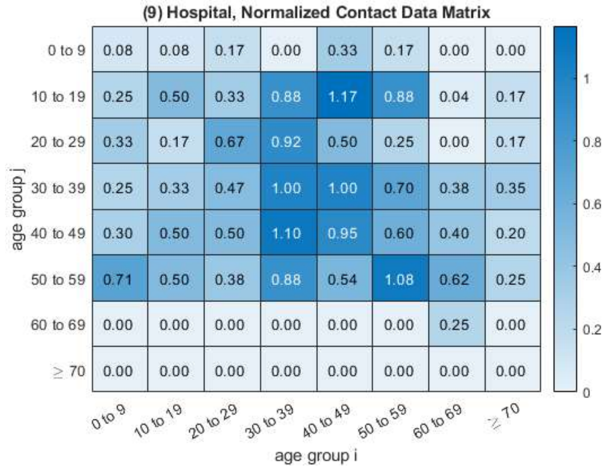
(9) Hospital, Normalized Contact Data Matrix
0 to 9 0.08 0.08 0.17 0.00 0.33 0.17 0.00 0.00
10 to 19 0.25 0.50 0.33 0.88 1.17 0.88 0.04 0.17
20 to 29 0.33 0.17 0.67 0.92 0.50 0.25 0.00 0.17
30 to 39 0.25 0.33 0.47 1.00 1.00 0.70 0.38 0.35
40 to 49 0.30 0.50 0.50 1.10 0.95 0.60 0.40 0.20
50 to 59 0.71 0.50 0.38 0.88 0.54 1.08 0.62 0.25
60 to 69 0.00 0.00 0.00 0.00 0.00 0.00 0.25 0.00
≥ 70 0.00 0.00 0.00 0.00 0.00 0.00 0.00 0.00
age group j
0 to 9 10 to 19 20 to 29 30 to 39 40 to 49 50 to 59 60 to 69 ≥ 70
age group i

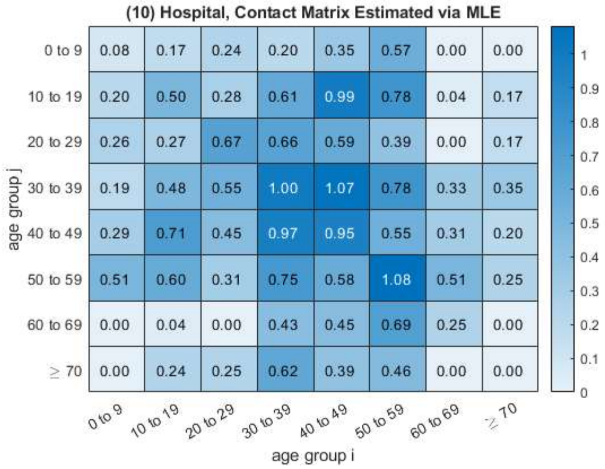
(10) Hospital, Contact Matrix Estimated via MLE
0 to 9 0.08 0.17 0.24 0.20 0.35 0.57 0.00 0.00
10 to 19 0.20 0.50 0.28 0.61 0.99 0.78 0.04 0.17
20 to 29 0.26 0.27 0.67 0.66 0.59 0.39 0.00 0.17
30 to 39 0.19 0.48 0.55 1.00 1.07 0.78 0.33 0.35
40 to 49 0.29 0.71 0.45 0.97 0.95 0.55 0.31 0.20
50 to 59 0.51 0.60 0.31 0.75 0.58 1.08 0.51 0.25
60 to 69 0.00 0.04 0.00 0.43 0.45 0.69 0.25 0.00
≥ 70 0.00 0.24 0.25 0.62 0.39 0.46 0.00 0.00
age group j
0 to 9 10 to 19 20 to 29 30 to 39 40 to 49 50 to 59 60 to 69 ≥ 70
age group i

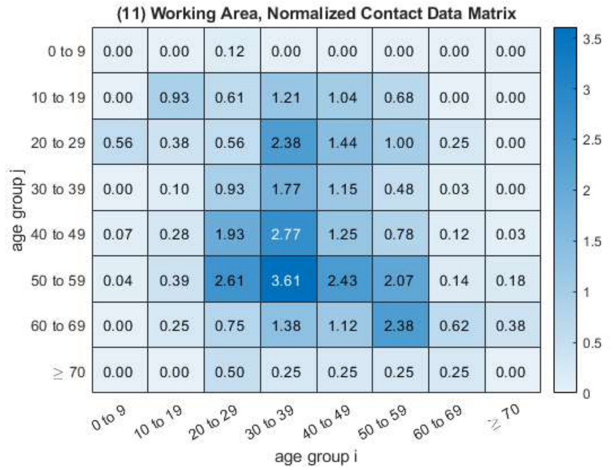
(11) Working Area, Normalized Contact Data Matrix
0 to 9 0.00 0.00 0.12 0.00 0.00 0.00 0.00 0.00
10 to 19 0.00 0.93 0.61 1.21 1.04 0.68 0.00 0.00
20 to 29 0.56 0.38 0.56 2.38 1.44 1.00 0.25 0.00
30 to 39 0.00 0.10 0.93 1.77 1.15 0.48 0.03 0.00
40 to 49 0.07 0.28 1.93 2.77 1.25 0.78 0.12 0.03
50 to 59 0.04 0.39 2.61 3.61 2.43 2.07 0.14 0.18
60 to 69 0.00 0.25 0.75 1.38 1.12 2.38 0.62 0.38
≥ 70 0.00 0.00 0.50 0.25 0.25 0.25 0.25 0.00
age group j
0 to 9 10 to 19 20 to 29 30 to 39 40 to 49 50 to 59 60 to 69 ≥ 70
age group i

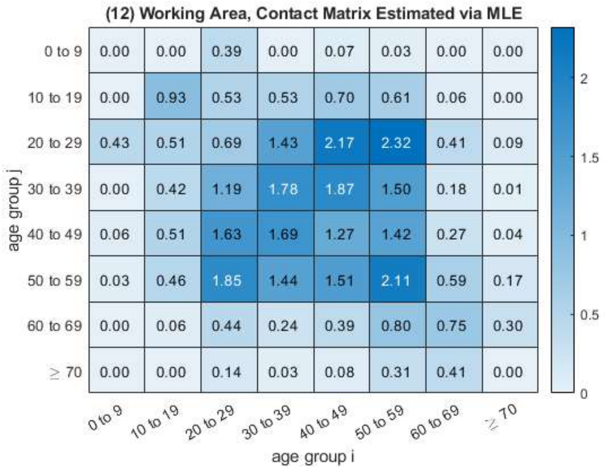
(12) Working Area, Contact Matrix Estimated via MLE
0 to 9 0.00 0.00 0.39 0.00 0.07 0.03 0.00 0.00
10 to 19 0.00 0.93 0.53 0.53 0.70 0.61 0.06 0.00
20 to 29 0.43 0.51 0.69 1.43 2.17 2.32 0.41 0.09
30 to 39 0.00 0.42 1.19 1.78 1.87 1.50 0.18 0.01
40 to 49 0.06 0.51 1.63 1.69 1.27 1.42 0.27 0.04
50 to 59 0.03 0.46 1.85 1.44 1.51 2.11 0.59 0.17
60 to 69 0.00 0.06 0.44 0.24 0.39 0.80 0.75 0.30
≥ 70 0.00 0.00 0.14 0.03 0.08 0.31 0.41 0.00
age group j
0 to 9 10 to 19 20 to 29 30 to 39 40 to 49 50 to 59 60 to 69 ≥ 70
age group i

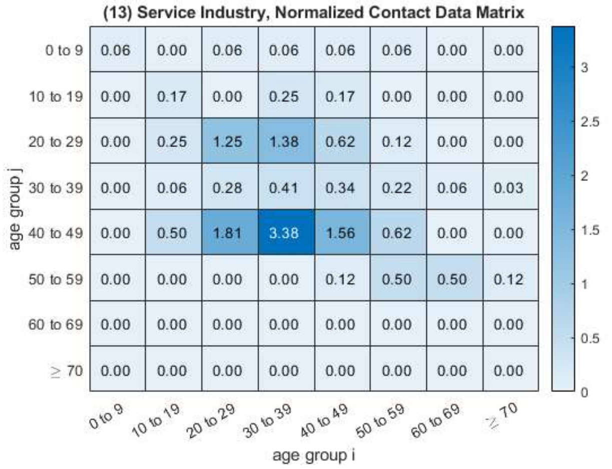
(13) Service Industry, Normalized Contact Data Matrix
age group j
age group i
0 to 9: 0.06 0.00 0.06 0.06 0.06 0.06 0.00 0.00
10 to 19: 0.00 0.17 0.00 0.25 0.17 0.00 0.00 0.00
20 to 29: 0.00 0.25 1.25 1.38 0.62 0.12 0.00 0.00
30 to 39: 0.00 0.06 0.28 0.41 0.34 0.22 0.06 0.03
40 to 49: 0.00 0.50 1.81 3.38 1.56 0.62 0.00 0.00
50 to 59: 0.00 0.00 0.00 0.00 0.12 0.50 0.50 0.12
60 to 69: 0.00 0.00 0.00 0.00 0.00 0.00 0.00 0.00
≥ 70: 0.00 0.00 0.00 0.00 0.00 0.00 0.00 0.00
0 to 9
10 to 19
20 to 29
30 to 39
40 to 49
50 to 59
60 to 69
≥ 70
0
0.5
1
1.5
2
2.5
3
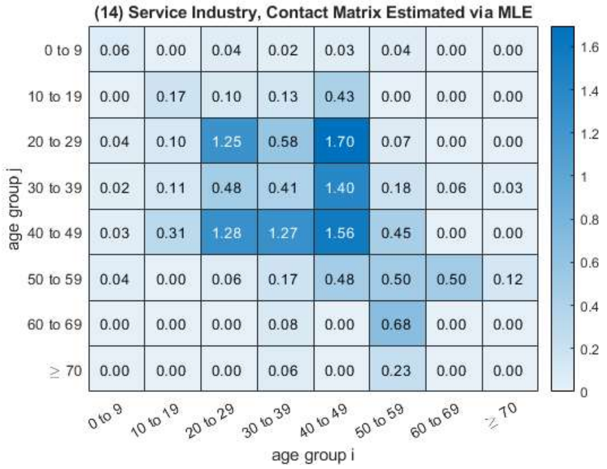
(14) Service Industry, Contact Matrix Estimated via MLE
age group j
age group i
0 to 9: 0.06 0.00 0.04 0.02 0.03 0.04 0.00 0.00
10 to 19: 0.00 0.17 0.10 0.13 0.43 0.00 0.00 0.00
20 to 29: 0.04 0.10 1.25 0.58 1.70 0.07 0.00 0.00
30 to 39: 0.02 0.11 0.48 0.41 1.40 0.18 0.06 0.03
40 to 49: 0.03 0.31 1.28 1.27 1.56 0.45 0.00 0.00
50 to 59: 0.04 0.00 0.06 0.17 0.48 0.50 0.50 0.12
60 to 69: 0.00 0.00 0.00 0.08 0.00 0.68 0.00 0.00
≥ 70: 0.00 0.00 0.00 0.06 0.00 0.23 0.00 0.00
0 to 9
10 to 19
20 to 29
30 to 39
40 to 49
50 to 59
60 to 69
≥ 70
0
0.2
0.4
0.6
0.8
1
1.2
1.4
1.6
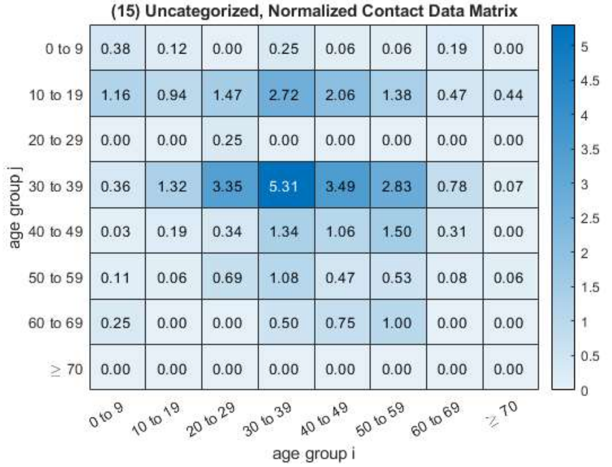
(15) Uncategorized, Normalized Contact Data Matrix
age group j
age group i
0 to 9: 0.38 0.12 0.00 0.25 0.06 0.06 0.19 0.00
10 to 19: 1.16 0.94 1.47 2.72 2.06 1.38 0.47 0.44
20 to 29: 0.00 0.00 0.25 0.00 0.00 0.00 0.00 0.00
30 to 39: 0.36 1.32 3.35 5.31 3.49 2.83 0.78 0.07
40 to 49: 0.03 0.19 0.34 1.34 1.06 1.50 0.31 0.00
50 to 59: 0.11 0.06 0.69 1.08 0.47 0.53 0.08 0.06
60 to 69: 0.25 0.00 0.00 0.50 0.75 1.00 0.00 0.00
≥ 70: 0.00 0.00 0.00 0.00 0.00 0.00 0.00 0.00
0 to 9
10 to 19
20 to 29
30 to 39
40 to 49
50 to 59
60 to 69
≥ 70
0
0.5
1
1.5
2
2.5
3
3.5
4
4.5
5
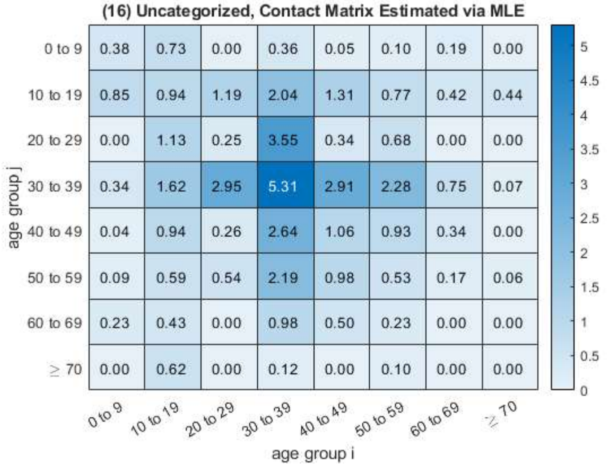
(16) Uncategorized, Contact Matrix Estimated via MLE
age group j
age group i
0 to 9: 0.38 0.73 0.00 0.36 0.05 0.10 0.19 0.00
10 to 19: 0.85 0.94 1.19 2.04 1.31 0.77 0.42 0.44
20 to 29: 0.00 1.13 0.25 3.55 0.34 0.68 0.00 0.00
30 to 39: 0.34 1.62 2.95 5.31 2.91 2.28 0.75 0.07
40 to 49: 0.04 0.94 0.26 2.64 1.06 0.93 0.34 0.00
50 to 59: 0.09 0.59 0.54 2.19 0.98 0.53 0.17 0.06
60 to 69: 0.23 0.43 0.00 0.98 0.50 0.23 0.00 0.00
≥ 70: 0.00 0.62 0.00 0.12 0.00 0.10 0.00 0.00
0 to 9
10 to 19
20 to 29
30 to 39
40 to 49
50 to 59
60 to 69
≥ 70
0
0.5
1
1.5
2
2.5
3
3.5
4
4.5
5

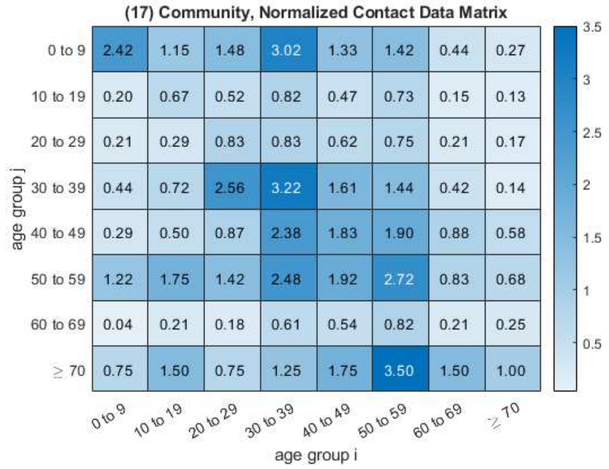


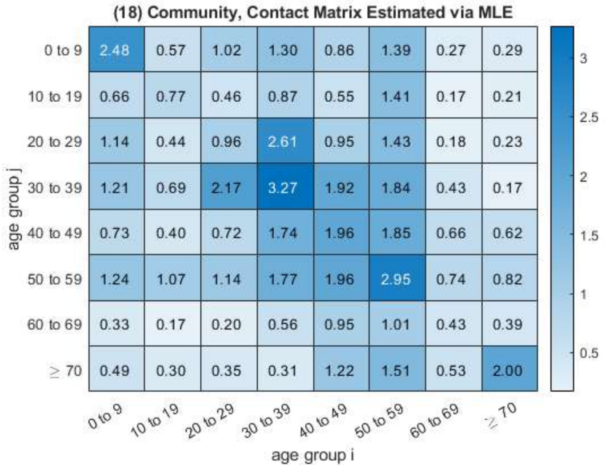


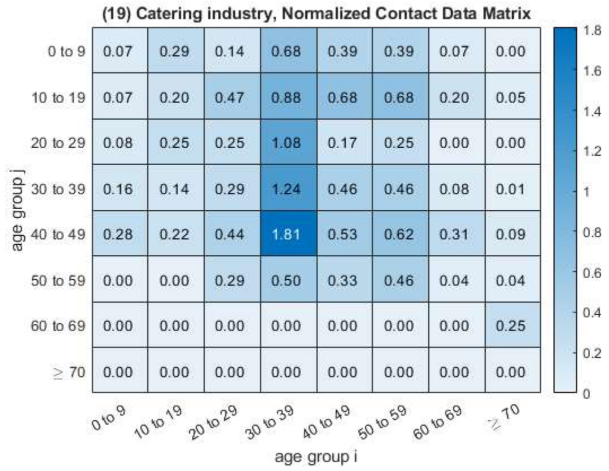


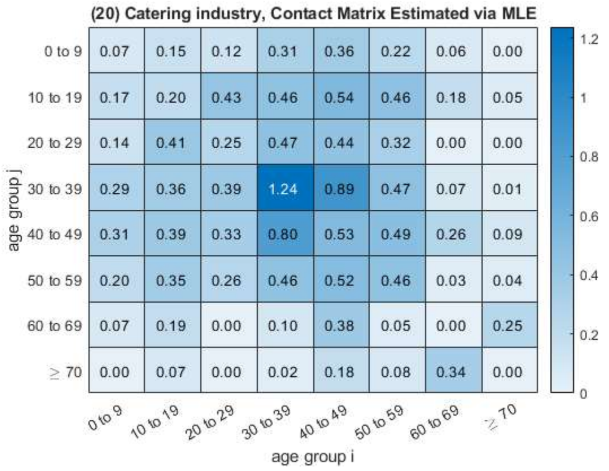


*Published with MATLAB® R2021b*

#### 5.2 NPI Simulations

We consider the contact matrix under certain NPI condition as a linear combination of those 10 matrices. For example, school closure means the coefficient of the school component equals 0.

### 6 Source Codes

#### 6.1 Construction of Contact Matrix

##### 6.1.1 Script: testContactMatrix_WholeTimeSegment

```
clear; close all; clc;

opts = detectImportOptions('caseContactData.xlsx');
opts.VariableTypes(1,[7,8]) = repmat({'datetime'},1,2);
data = readtable('caseContactData.xlsx',opts);


%% relevent Data Vectors
% caseAge, contactAge, lastContact, illnessOnset, diagnosedDate, caseNames

% caseNames
caseNames = data.caseNameIncaseData;
[uniqueCaseNames,uniqueCaseIndex] = unique(caseNames);

emptyID = cellfun(@isempty,caseNames);
data(emptyID,:) = [];
%data(isnan(data.contactAge),:) = [];


caseNames = data.caseNameIncaseData;
[uniqueCaseNames,uniqueCaseIndex] = unique(caseNames);


% caseAge, contactAge
caseAge = data{:,3};

contactAge = data{:,15};
n = numel(caseAge);
```

```
% lastContact, illnessOnset, diagnosedDate
lastContact = data{:,20};
illnessOnset = data{:,7};
diagnosedDate = data{:,8};


% duration between illnessOnset and been diagnosed
sum(diagnosedDate == illnessOnset)
sum(isnat(diagnosedDate))
sum(isnat(illnessOnset))

D = diagnosedDate - illnessOnset;
durationInDays = day(D) + 1;

%% Construct A, the Contact Data Matrix
% contactDataMatrix(i,j) denotes the daily average number of close contact in group
% j, will produced by a individual in group i.

% Assumption 1:  the contact are uniformly distributed
% Assumption 2:  contactMatrix is symmetric
% Assumption 3:  contacts in data meets the case only once
% Assumption 4: all contacts of one particular case are recorded for a four-day period before ...
    the date fo been diagnosed

% age partition
agePartition = [0,10,20,30,40,50,60,70,200]; %linspace(0,100,11); % [0,10,20,...,100]
[contactDataMatrix, caseRepeatCount] = computeContactDataMatrix(caseNames, caseAge, contactAge, ...
    agePartition);

% heatmap of un-normalized contactDataMatrix
figure;
format shorte;
xvalue = {'0 to 9','10 to 19','20 to 29', '30 to 39', '40 to 49', '50 to 59', '60 to 69', '\geq ...
    70'};
yvalue = {'0 to 9','10 to 19','20 to 29', '30 to 39', '40 to 49', '50 to 59', '60 to 69', '\geq ...
    70'};
h_DEF = heatmap(xvalue, yvalue, contactDataMatrix);
h_DEF.Title = 'un-normalized contactDataMatrix';
h_DEF.XLabel = 'age group i';
h_DEF.YLabel = 'age group j';
```

```
h_DEF.CellLabelFormat = '%.2f ';

% normalize the contactDataMatrix by caseRepeatCount
normalizedContactDataMatrix = contactDataMatrix ./ caseRepeatCount;

figure;
format shorte;
xvalue = {'0 to 9','10 to 19','20 to 29', '30 to 39', '40 to 49', '50 to 59', '60 to 69', '\geq ...
    70'};
yvalue = {'0 to 9','10 to 19','20 to 29', '30 to 39', '40 to 49', '50 to 59', '60 to 69', '\geq ...
    70'};
h_DEF = heatmap(xvalue, yvalue, normalizedContactDataMatrix);
h_DEF.Title = 'contactDataMatrix for age groups';
h_DEF.XLabel = 'age group i';
h_DEF.YLabel = 'age group j';
h_DEF.CellLabelFormat = '%.2f ';
exportgraphics(gca,'contactDataMatrixGroupedByAges.jpg','Resolution',300);


%% read population Data
populationData = readtable('populationData.xlsx');
head(populationData)
ageGroup = populationData.ageGroup;
population = populationData.all;

agePopulation = zeros(numel(agePartition)-1,1);
for i = 1:numel(agePartition)-1
    agePopulation(i) = sum(population(ageGroup≥agePartition(i) & ageGroup < ...
        agePartition(i+1)),'omitnan');
end
writematrix(agePopulation,'agePopulationVector.xlsx');

%% least square estimation of contactMatrix C
C_LSE = estimateContactMatrix_LSE(normalizedContactDataMatrix, caseRepeatCount, agePopulation);
exportgraphics(gca,'contactMatrixGroupedByAges_LSE.jpg','Resolution',300);

%% weighted least square estimation of contactMatrix C
C_WLSE = estimateContactMatrix_WLSE(normalizedContactDataMatrix, caseRepeatCount, agePopulation);
exportgraphics(gca,'contactMatrixGroupedByAges_WLSE.jpg','Resolution',300);

%% maximum likelihood estimation of contact matrix C
```

```
C_MLE = estimateContactMatrix_MLE(contactDataMatrix,caseRepeatCount, agePopulation);
exportgraphics(gca,'contactMatrixGroupedByAges_MLE.jpg','Resolution',300);

%% entrywise 95% confidential intervals (based MLE)
C_lower = icdf('Poisson',0.025,C_MLE);
C_upper = icdf('Poisson',0.975,C_MLE);

figure;
format shorte;
xvalue = {'0 to 9','10 to 19','20 to 29', '30 to 39', '40 to 49', '50 to 59', '60 to 69', '\geq ...
    70'};
yvalue = {'0 to 9','10 to 19','20 to 29', '30 to 39', '40 to 49', '50 to 59', '60 to 69', '\geq ...
    70'};
h_lower = heatmap(xvalue, yvalue, C_lower);
h_lower.Title = 'the lower bounds of the entrywise 95% CI of the contactMatrix';
h_lower.XLabel = 'age group i';
h_lower.YLabel = 'age group j';
h_lower.CellLabelFormat = '%.2f ';

figure;
format shorte;
xvalue = {'0 to 9','10 to 19','20 to 29', '30 to 39', '40 to 49', '50 to 59', '60 to 69', '\geq ...
    70'};
yvalue = {'0 to 9','10 to 19','20 to 29', '30 to 39', '40 to 49', '50 to 59', '60 to 69', '\geq ...
    70'};
h_upper = heatmap(xvalue, yvalue, C_upper);
h_upper.Title = 'the upper bounds of the entrywise 95% CI of the contactMatrix';
h_upper.XLabel = 'age group i';
h_upper.YLabel = 'age group j';
h_upper.CellLabelFormat = '%.2f ';


%% write MLE C, C_lower, C_upper
cd('H:\xmuph\0 HunanCovid-19 paper\Codes\step3_SEIAR_and_VEFIAQR_Simulation');
writematrix(C_MLE,'contactMatrix.xlsx');
writematrix(C_lower,'contactMatrixLowerBound.xlsx');
writematrix(C_upper,'contactMatrixUpperBound.xlsx');
cd('H:\xmuph\0 HunanCovid-19 paper\Codes\step2_Construction of ContactMatrix');

%% export figures
exportgraphics(h_upper,'contactMatrixGroupedByAges_MLE_upperCI.jpg','Resolution',300);
```

```
exportgraphics(h_lower,'contactMatrixGroupedByAges_MLE_lowerCI.jpg','Resolution',300);
```

##### 6.1.2 Script: testContactMatrix_TwoTimeSegment

```
clear; close all; clc;

opts = detectImportOptions('caseContactData.xlsx');
opts.VariableTypes(1,[7,8]) = repmat({'datetime'},1,2);
data = readtable('caseContactData.xlsx',opts);


% caseNames
caseNames = data.caseNameIncaseData;
emptyID = cellfun(@isempty,caseNames);
data(emptyID,:) = [];

caseNames = data.caseNameIncaseData;
[uniqueCaseNames,uniqueCaseIndex] = unique(caseNames);

% diagnoseDate
diagnoseDate = data.diagnoseDate(uniqueCaseIndex,1);

minDate = dateshift(min(diagnoseDate),'start','day')
maxDate = dateshift(max(diagnoseDate),'end','day')
edges = minDate:maxDate;
histogram(diagnoseDate, edges);
%set(gca,'xticklabel',['', sortedNames(id)]);

plot(diagnoseDate, data.caseAge(uniqueCaseIndex), 'rp');

% datetime partition threshold
 tau = datetime(2021,08,04);
%tau = datetime(2021,08,07);
% tau = datetime(2021,08,01);
% tau = datetime(2021,08,14);
data1 = data(data.diagnoseDate ≤ tau,:);
data2 = data(data.diagnoseDate > tau,:);

% age partition
```

```
agePartition = [0,10,20,30,40,50,60,70,200]; %linspace(0,100,11); % [0,10,20,...,100]

[contactDataMatrix, caseRepeatCount] = computeContactDataMatrix(data.caseNameIncaseData, ...
    data.caseAge, data.contactAge, agePartition);
[contactDataMatrix_2, caseRepeatCount_2] = computeContactDataMatrix(data2.caseNameIncaseData, ...
    data2.caseAge, data2.contactAge, agePartition);
[contactDataMatrix_1, caseRepeatCount_1] = computeContactDataMatrix(data1.caseNameIncaseData, ...
    data1.caseAge, data1.contactAge, agePartition);


%% read agePopulation
agePopulation = readmatrix('agePopulationVector.xlsx');


%% maximum likelihood estimation of contact matrix C_1, C_2
C1_MLE = estimateContactMatrix_MLE(contactDataMatrix_1, caseRepeatCount_1, agePopulation, 0);
C2_MLE = estimateContactMatrix_MLE(contactDataMatrix_2, caseRepeatCount_2, agePopulation, 0);

figure;
heatmap(double(C1_MLE>C2_MLE));
sum(C1_MLE>C2_MLE,'all')

C1_MLE(isnan(C1_MLE)) = 0;
lambda_max_C1 = max(eig(C1_MLE))
lambda_max_C2 = max(eig(C2_MLE))

%%

figure;
format shorte;
xvalue = {'0 to 9','10 to 19','20 to 29', '30 to 39', '40 to 49', '50 to 59', '60 to 69', '\geq ...
    70'};
yvalue = {'0 to 9','10 to 19','20 to 29', '30 to 39', '40 to 49', '50 to 59', '60 to 69', '\geq ...
    70'};
h_DEF = heatmap(xvalue, yvalue, C1_MLE);
h_DEF.Title = 'contactMatrix of first segment, estimated by MLE';
h_DEF.XLabel = 'age group i';
h_DEF.YLabel = 'age group j';
h_DEF.CellLabelFormat = '%.2f ';

figure;
```

```
h_DEF = heatmap(xvalue, yvalue, C2_MLE);
h_DEF.Title = 'contactMatrix of second segment, estimated by MLE';
h_DEF.XLabel = 'age group i';
h_DEF.YLabel = 'age group j';
h_DEF.CellLabelFormat = '%.2f ';

figure;
plot(C1_MLE(:),C2_MLE(:),'rp'); hold on;
t = linspace(0,12,1e3);

% linearEq = fit(C1_MLE(:), C2_MLE(:), 'linear');
% k = predict(linearEq,t);

plot(t,t,'b-');
```

##### 6.1.3 Function: estimateContactMatrix_LSE

```
function C = estimateContactMatrix_LSE(normalizedContactDataMatrix, caseRepeatCount, ...
    agePopulation, varargin)
%%% least square estimation of contactMatrix C

if nargin ≤ 3
    plotFlag = 1; % plot the heatmap as a default
else
    plotFlag = varargin{1};
end




A = normalizedContactDataMatrix;
N = agePopulation;
groupCount = size(A,1);
C = zeros(groupCount);

for i = 1 : groupCount-1
    for j = i+1 : groupCount
        C(i,j) = ( A(i,j) + N(i)/N(j) * A(j,i) ) / (1 + N(i)^2/N(j)^2);
        C(j,i) = N(i)/N(j) * C(i,j);
```

```
        end
    end

    for i = 1 : groupCount
        C(i,i) = A(i,i);
    end

    figure;
    format shorte;
    xvalue = {'0 to 9','10 to 19','20 to 29', '30 to 39', '40 to 49', '50 to 59', '60 to 69', '\geq ...
        70'};
    yvalue = {'0 to 9','10 to 19','20 to 29', '30 to 39', '40 to 49', '50 to 59', '60 to 69', '\geq ...
        70'};
    h_DEF = heatmap(xvalue, yvalue, C);
    h_DEF.Title = 'contactMatrix for age groups, estimated by LS';
    h_DEF.XLabel = 'age group i';
    h_DEF.YLabel = 'age group j';
    h_DEF.CellLabelFormat = '%.2f ';
    end
```

##### 6.1.4 Function: estimateContactMatrix_WLSE

```
function C = estimateContactMatrix_LSE(normalizedContactDataMatrix, caseRepeatCount, ...
    agePopulation, varargin)
%%% weighted least square estimation of contactMatrix C

if nargin ≤ 3
    plotFlag = 1; % plot the heatmap as a default
else
    plotFlag = varargin{1};
end


A = normalizedContactDataMatrix;
N = agePopulation;
n = caseRepeatCount;

groupCount = size(A,1);
C = zeros(groupCount);
```

```

for i = 1 : groupCount-1
    for j = i+1 : groupCount
       C(i,j) = ( n(i)*A(i,j) + n(j)*N(i)/N(j) * A(j,i) ) / (n(i) + N(i)^2/N(j)^2*n(j));
       C(j,i) = N(i)/N(j) * C(i,j);
    end
end

for i = 1 : groupCount
    C(i,i) = A(i,i);
end

figure;
format shorte;
xvalue = {'0 to 9','10 to 19','20 to 29', '30 to 39', '40 to 49', '50 to 59', '60 to 69', '\geq ...
    70'};
yvalue = {'0 to 9','10 to 19','20 to 29', '30 to 39', '40 to 49', '50 to 59', '60 to 69', '\geq ...
    70'};
h_DEF = heatmap(xvalue, yvalue, C);
h_DEF.Title = 'contactMatrix for age groups, estimated by WLS';
h_DEF.XLabel = 'age group i';
h_DEF.YLabel = 'age group j';
h_DEF.CellLabelFormat = '%.2f ';

end
```

##### 6.1.5 Function: estimateContactMatrix_MLE

```
function contactMatrix = estimateContactMatrix_MLE(contactDataMatrix,caseRepeatCount, ...
    agePopulation, varargin)
%%% maximum likelihood estimation of contact matrix C

if nargin ≤ 3
    plotFlag = 1; % plot the heatmap as a default
else
    plotFlag = varargin{1};
end


```

```
N = agePopulation;
groupCount = size(contactDataMatrix,1);

C = zeros(groupCount);

for i = 1:groupCount-1
    for j = i+1:groupCount
        Ki = caseRepeatCount(i);
        Kj = caseRepeatCount(j);
        C(i,j) = (contactDataMatrix(i,j) + contactDataMatrix(j,i)) / (Ki+N(i)/N(j)*Kj);
        C(j,i) = N(i)/N(j) * C(i,j);
    end
end

for i = 1:groupCount
    Ki = caseRepeatCount(i);
    C(i,i) = (contactDataMatrix(i,i) + contactDataMatrix(j,i)) / Ki;
end

contactMatrix = C;

if plotFlag == 1
    figure;
    format shorte;
    xvalue = {'0 to 9','10 to 19','20 to 29', '30 to 39', '40 to 49', '50 to 59', '60 to 69', ...
        '\geq 70'};
    yvalue = {'0 to 9','10 to 19','20 to 29', '30 to 39', '40 to 49', '50 to 59', '60 to 69', ...
        '\geq 70'};
    h_DEF = heatmap(xvalue, yvalue, C);
    h_DEF.Title = 'contactMatrix for age groups, estimated by MLE';
    h_DEF.XLabel = 'age group i';
    h_DEF.YLabel = 'age group j';
    h_DEF.CellLabelFormat = '%.2f ';
end
end
```

##### 6.1.6 Function: whichGroup

```
function group = whichGroup(age, agePartition)
```

```
% given a partition like: agePartition = [0,10,20,30,40,50,60,70,200];
% input a scalar age
% ouput the group that such age belongs to.

groupCount = numel(agePartition) - 1;

group = 0;
for k = 1:groupCount
    if age ≥ agePartition(k) && age < agePartition(k+1)
        group = k;
        continue;
    end
end


end
```

#### 6.2 $SEIAR$ Simulation

##### 6.2.1 Script: ImportParametersSEIAR

```
n = 8;  % number of subgroups
params.kappa  = 0.7 * ones(n,1);
params.p      = 0.3 * ones(n,1);
params.omega  = 1/3 * ones(n,1);
params.omegap = 1/5 * ones(n,1);
params.gamma  = 1/5 * ones(n,1);
params.gammap = 1/10 * ones(n,1);
params.f      = 0 * ones(n,1);
params.br     = 0 * ones(n,1);
params.dr     = 0 * ones(n,1);
kappa = params.kappa;
p = params.p;


omega = params.omega;
omegap = params.omegap;
gamma = params.gamma;
gammap = params.gammap;
```

```
f = params.f;
br = params.br;
dr = params.dr;
kappa = params.kappa;
p = params.p;
```

##### 6.2.2 Script: testSimulationSEIAR

```
clear all; close all; clc;

ImportParametersSEIAR;   % import parameters omega, gamma, p, kappa,...
ImportFigureLegends; % import the following cell array for figure legend:
                     % groupLegend; ageLegend;
N = readmatrix('agePopulationVector.xlsx'); % the population vector (stratified by age)
C = readmatrix('contactMatrix.xlsx');   % the contact matrix

%% steup simulation

% initial value
xInit = [N, zeros(n,4)];
xInit(4,3) = 1;

% time span
tInit = 0;
tFinal = 14;

% step size
stepSize = 0.01;

% a series R0 for simulation
R0 = [0.2, 0.5:6.5];     R0 = R0';



%% probability vector q (DBM and NGM are adopted respectively), for infection by a one-time contact
q_DBM = zeros(numel(R0),1);
q_NGM = zeros(numel(R0),1);

for i = 1:n
```

```
    for j = 1:n
        R(i,j) = C(j,i) / (dr(i) + p(i)*omegap(i) + (1-p(i))*omega(i)) * ...
            (kappa(i)*p(i)*omegap(i)/(gammap(i)+dr(i)) + (1-p(i))*omega(i)/(gamma(i)+dr(i)+f(i)));
    end
end


for i = 1:numel(R0)
    temp1 = R .* (N.') ./ sum(N);
    temp2 = sum(temp1,1);
    q_DBM(i) = R0(i) / sum(temp2);

    temp3 = 1 / (dr(i) + p(i)*omegap(i) + (1-p(i))*omega(i)) * ...
        (kappa(i)*p(i)*omegap(i)/(gammap(i)+dr(i)) + (1-p(i))*omega(i)/(gamma(i)+dr(i)+f(i)));
    q_NGM(i) = R0(i) / (max(eig(C)) * temp3);
end
fprintf('[R0, q_DBM, q_NGM] = \n');
disp([R0, q_DBM, q_NGM]);



%% simulate for each self-defined base-line R0
for i = 1:numel(R0)
    params.Beta   = C * q_DBM(i) ./ (N');
%     params.Beta   = C * q_NGM(i) ./ (N');

    fun = @(x,t)dxdt_SEIAR(x,t,params);
    [x,t] = odeSolveRK4(fun, tInit, xInit, tFinal, stepSize);
    [S,E,I,A,R] = extractVariableFromCells_forSEIAR(x);

    % currentSymptomaticCases
    figure;
    plot(t,I);
    legend(ageLegend);
    title(['current symptomatic cases \it{I}, with R_0 = ', num2str(R0(i))]);
    xlabel('time (in days)');
    ylabel('population size (in person)')

    % dailySymptomaticNewCases
    figure;
    dailySymptomaticNewCases = ((1-p).*omega)' .* E;
```

```
    plot(t,dailySymptomaticNewCases);
    legend(ageLegend);
    title(['daily new symptomatic cases \it{(1-p) \omega E}, with R_0 = ', num2str(R0(i))]);
    xlabel('time (in days)');
    ylabel('population size (in person)')

    % allCurrentCases
    figure;
    allCurrentCases = I + A;
    plot(t,allCurrentCases);
    legend(ageLegend);
    title(['all current cases \it{I + A}, with R_0 = ', num2str(R0(i))]);
    xlabel('time (in days)');
    ylabel('population size (in person)')

    % accumulativeNumberOfCases
    figure;
    dailyNewCases = (p.*omegap)' .* E + ((1-p).*omega)' .* E;
    accumulativeNumberOfCases = zeros(size(dailyNewCases,1), n);
    for k = 1:n
        accumulativeNumberOfCases(:,k) = cumtrapz(t, dailyNewCases(:,k));
    end
    plot(t,accumulativeNumberOfCases);
    legend(ageLegend);
    title(['accumulative number of cases, with R_0 = ', num2str(R0(i))]);
    xlabel('time (in days)');
    ylabel('population size (in person)')


    % how to define and compute the accumulativeNumberOfCases from model directly
    % instead of a extra numerical integration, it is still an very interesting question.
    Nt = sum(S + E + I + A + R, 2) / sum(N);
end
```

##### 6.2.3 Script: ImportFigureLegends

```
%%% groupLegend
groupLegend = cell(n,1);
for i = 1:n
```

```
    groupLegend{i,1} = ['group ',num2str(i)];
end

%%% ageLegends
ageLegend = {'0 to 9','10 to 19','20 to 29', '30 to 39',...
             '40 to 49', '50 to 59', '60 to 69', '\geq 70' };



%%% doseLegends
doseLegend = {'none', 'unfully vaccinated', 'fully vaccinated', 'booster vaccinated'};
```

##### 6.2.4 Function: odeSolveRK4

```
function [varargout] = odeSolveRK4(fun, tInit, xInit, tFinal, stepSize)
%%% Solve the Initial Value Problem using Rungue Kutta 4th Order Method
%   INPUT:
%       fun           : function handle describe the differencial equation dx/dt = ...
    @(x,t)f(x,t,params)
%       tInit         : scalar, time instance of the initial point
%       xInit         : row vector, values of the initial point
%       tFinal        : solve the ode until t = tFinal
%       stepSize      : fixed stepsize for update
%
%   OUTPUT:
%       x             : m*5 matrix, the solution of the ODE
%       t             : time instances corresponding to x
%       I             : m*n matrix, I_i for each group
%
% by Guo Xiaohao, 2021/09/23
%

tSpan = tInit : stepSize : tFinal;
m = numel(tSpan);
[n,¬] = size(xInit);

x = cell(m,1);
x{1} = xInit;

```

```
for i = 1:m
    x{i+1} = updateRK4(fun, tSpan(i), x{i}, stepSize);
end

t = [tSpan, tFinal+stepSize]';

varargout{1} = x;
varargout{2} = t;


end
```

###### 6.2.5 Function: updateRK4

```
function xNext = updateRK4(fun, tCurrent, xCurrent, stepSize)
%%% one-step iteration for solving the differencial equation
%%% dx/dt = @(x,t)f(x,t,params) by method of RK4
% INPUT
%        fun: a function handle describe f(x,t)
%   xCurrent: m*n matrix, current x, to be updated
%   tCurrent: a scalar, the current t
%   stepSize: stepSize for updating
%    params : all parameters for the f(x,t,params)
%
% by Guo Xiaohao, 2021/09/23
%

K1 = stepSize * fun(xCurrent, tCurrent);  % m*n matrix
K2 = stepSize * fun(xCurrent + K1/2, tCurrent + stepSize/2);
K3 = stepSize * fun(xCurrent + K2/2, tCurrent + stepSize/2);
K4 = stepSize * fun(xCurrent + K3, tCurrent + stepSize);

xNext = xCurrent + (K1 + 2*K2 + 2*K3 + K4) / 6;

end
```

###### 6.2.6 Function: extractVariableFromCells_forSEIAR

```
function [S,E,I,A,R] = extractVariableFromCells_forSEIAR(x)



m = numel(x);
[n,¬] = size(x{1});

% save S
if nargout ≥ 1
    S = zeros(m,n);
    for i = 1:m
        S(i,:) = (x{i}(:,1)).';
    end
    varargout{1} = S;
end

% save E
if nargout ≥ 2
    E = zeros(m,n);
    for i = 1:m
        E(i,:) = (x{i}(:,2)).';
    end
    varargout{2} = E;
end

% save I
if nargout ≥ 3
    I = zeros(m,n);
    for i = 1:m
        I(i,:) = (x{i}(:,3)).';
    end
    varargout{3} = I;
end


% save A
if nargout ≥ 4
    A = zeros(m,n);
    for i = 1:m
        A(i,:) = (x{i}(:,4)).';
```

```
    end
    varargout{4} = A;
end


% save R
if nargout ≥ 5
    R = zeros(m,n);
    for i = 1:m
        R(i,:) = (x{i}(:,5)).';
    end
    varargout{5} = R;
end
```

##### 6.2.7 Function: dxdt_SEIAR

```
function dxdt = dxdt_SEIAR(x,t,params)
%%% Consider the differencial equation: dx/dt = f(x,t,params), where x denotes a vector of
%%% m dimension. (Let n denotes the number of subgroups, and m = 5*n)
%%% This function evaluates the derivative f(x,t) at a given point (x,t)
% INPUT:
%  x = [S,E,I,A,R], a n*5 matrix, where:
%       S(t): a vector [S1(t), S2(t), ..., Sn(t)]';
%       E(t): a vector [E1(t), E2(t), ..., En(t)]';
%       I(t): a vector [I1(t), I2(t), ..., In(t)]';
%       A(t): a vector [A1(t), A2(t), ..., An(t)]';
%       R(t): a vector [R1(t), R2(t), ..., Rn(t)]';
%         t : time variable
%       params: a struct of model parameters (assume that all parameters are group-distincted)
%
%       params.Beta  : n*n matrix, whose entries are beta_ij (beta_ij denotes the transimission ...
    capacity from group i to group j)
%       params.kappa : describe the difference of Transmission capacity between I and A
%       params.omega : the inverse of average incubation period of I
%       params.omegap: (omegap is the short of omega_prime) the inverse of average incubation ...
    period of A
%       params.p     : proportion of asymptomatic cases
%       params.gamma : the inverse of infectious period of I
```

```
%       params.gammap: (gammap is the short of gamma_prime) the inverse of average infectious ... period of A
%       params.f     : a column vector of n dimension, with its i-th entry denotes the fatality ... rate of I_i
%       params.br    : a column vector of n dimension, with its i-th entry denotes the birth ... rate of group i
%       params.dr    : a column vector of n dimension, with its i-th entry denotes the natrual ... death rate of group i
%
%  NOTE:
%       matrices: Beta
%       vectors : kappa, omega, omegap, p, gamma, gammap, f, br, dr, S, E, I, A, R
%       scalars : none (assume that all parameters are group distincted)
%
%
% OUTPUT:
%       f = f(x,t,params), a vector of derivatives
%

S = x(:,1);
E = x(:,2);
I = x(:,3);
A = x(:,4);
R = x(:,5);
N = S + E + I + A + R;

Beta = params.Beta;
kappa = params.kappa;
omega = params.omega;
omegap = params.omegap;
p = params.p;
gamma = params.gamma;
gammap = params.gammap;
f = params.f;
br = params.br;
dr = params.dr;

newlyInfectionTerm = sum(Beta.*(I + kappa.*A))' .* S;

dSdt = br.*N - dr.*S - newlyInfectionTerm;
dEdt = newlyInfectionTerm - ((1-p).*omega + p.*omegap + dr) .* E;
```

```
dIdt = (1-p).*omega.*E - (dr + f + gamma) .* I;
dAdt = p.*omegap.*E - (dr + gammap) .* A;
dRdt = gamma.*I + gammap.*A - dr.*R;

dxdt = [dSdt, dEdt, dIdt, dAdt, dRdt];




end
```

#### 6.3 $VEFIAQR$ Simulation

##### 6.3.1 Script: ImportParametersVEFIAQR

```
n = 8;  % number of subgroups
params.kappa  = 0.7 * ones(n,1);
params.p      = 0.3 * ones(n,1);
params.omega  = 1/3 * ones(n,1);
params.omegap = 1/5 * ones(n,1);
params.gamma  = 1/5 * ones(n,1);
params.gammap = 1/10 * ones(n,1);
params.f      = 0 * ones(n,1);
params.br     = 0 * ones(n,1);
params.dr     = 0 * ones(n,1);
kappa = params.kappa;
p = params.p;


omega = params.omega;
omegap = params.omegap;
gamma = params.gamma;
gammap = params.gammap;
f = params.f;
br = params.br;
dr = params.dr;
kappa = params.kappa;
p = params.p;
```

##### 6.3.2 Script: testSimulationVEFIAQR

```
clear all; close all; clc;

ImportParametersVEFIAQR;   % import parameters omega, gamma, p, kappa,...
ImportFigureLegends; % import the following cell array for figure legend:
                     % groupLegend; ageLegend;
N = readmatrix('agePopulationVector.xlsx'); % the population vector (stratified by age)
C = readmatrix('contactMatrix.xlsx');   % the contact matrix

%% steup simulation

% initial point
nVC = vaccineCoverageViaDataByIsComplete();
feasibleVC = nVC ./ sum(nVC,2);
xInit = [feasibleVC .* N, zeros(n,24)];
doseCount = sum(xInit(:,1:4), 1);
xInit(6,16) = 1;        % first case in I_{6,3}
% xInit(4,13) = 1;        % first case in I_{4,0}

% time span
tInit = 0;
tFinal = 100;

% step size
stepSize = 0.01;

% a series R0 for simulation
%R0 = [0.2, 0.5:6.5];     R0 = R0';
R0 = [0.5, 1, 1.5, 2.5, 3, 3.5, 4, 7]';


%% probability vector q (DBM and NGM are adopted respectively), for infection by a one-time contact
q_DBM = zeros(numel(R0),1);
q_NGM = zeros(numel(R0),1);

for i = 1:n
    for j = 1:n
        R(i,j) = C(j,i) / (dr(i) + p(i)*omegap(i) + (1-p(i))*omega(i)) * ...
            (kappa(i)*p(i)*omegap(i)/(gammap(i)+dr(i)) + (1-p(i))*omega(i)/(gamma(i)+dr(i)+f(i)));
```

```
    end
end


for i = 1:numel(R0)
    temp1 = R .* (N.') ./ sum(N);
    temp2 = sum(temp1,1);
    q_DBM(i) = R0(i) / sum(temp2);

    temp3 = 1 / (dr(i) + p(i)*omegap(i) + (1-p(i))*omega(i)) * ...
        (kappa(i)*p(i)*omegap(i)/(gammap(i)+dr(i)) + (1-p(i))*omega(i)/(gamma(i)+dr(i)+f(i)));
    q_NGM(i) = R0(i) / (max(eig(C)) * temp3);
end
fprintf('[R0, q_DBM, q_NGM] = \n');
disp([R0, q_DBM, q_NGM]);



%% simulate for each self-defined base-line R0
% figure(1)
% for i = 1:numel(R0)
%     fig1(i) = subplot(2,ceil(numel(R0) / 2), i);
% end
%
% figure(2)
% for i = 1:numel(R0)
%     fig2(i) = subplot(2,ceil(numel(R0) / 2), i);
% end

for i = 1:numel(R0)
%     params.Beta   = C * q_DBM(i) ./ (N');
    params.Beta   = C * q_NGM(i) ./ (N');

    fun = @(x,t)dxdt_VEFIAQR(x,t,params);
    [x,t] = odeSolveRK4(fun, tInit, xInit, tFinal, stepSize);
    [V, E, F, I, A, Q, R] = extractVariableFromCells_forVEFIAQR(x);


%     % currentSymptomaticCases
%     figure;
%     I_by_doses = extractByVaccineDoses(I) ./ doseCount ;
```

```
%     plot(t,I_by_doses);
%     legend(doseLegend);
%     title(['current symptomatic cases \it{I}, with R_0 = ', num2str(R0(i))]);
%     xlabel('time (in days)');
%     ylabel('Ratio of current symptomatic cases by doses')

    % dailySymptomaticNewCases
    dailyNewSymptomaticCases_by_doses = extractByVaccineDoses((1-p(:)').*omegap(:)'.*F) ./ ...
        doseCount;
    figure(1);
    subplot(2,ceil(numel(R0) / 2), i);
    plot(t, dailyNewSymptomaticCases_by_doses);
    legend(doseLegend);
    title(['R_0 = ', num2str(R0(i))]);
    sgtitle('Daily Incidence Rate by Doses with Simulated R_0');
    xlabel('time (in days)');
    ylabel('Daily Incidence Rate (I) by Doses')

%     % allCurrentCases
%     figure;
%     allCurrentCases = extractByVaccineDoses(I+A) ./ doseCount;
%     plot(t,allCurrentCases);
%     legend(doseLegend);
%     title(['all current cases \it{I + A}, with R_0 = ', num2str(R0(i))]);
%     xlabel('time (in days)');
%     ylabel('Ratio of current cases by doses')
%


    % accumulativeNumberOfCases
    % reshapedDailyNewCases

    dailyNewCases = extractByVaccineDoses(p(:)'.*omegap(:)'.*E + (1-p(:)').*omega(:)'.*E);
    reshapedDailyNewCases = extractByDosesAndGroups(p(:)'.*omegap(:)'.*E + (1-p(:)').*omega(:)'.*E);
    accumulativeNumberOfCases = zeros(size(dailyNewCases,1), 4);
    TotalIncidenceRate = squeeze(trapz(t, reshapedDailyNewCases,1)) ./ (feasibleVC .* N);

    for k = 1:4
        accumulativeNumberOfCases(:,k) = cumtrapz(t, dailyNewCases(:,k));
    end

%     if R0(i) == 4
```

```
%         1;
%     end

    % plot(t,accumulativeNumberOfCases ./ doseCount);
    figure(2);
    subplot(2,ceil(numel(R0) / 2), i);
    TotalIncidenceRate(isnan(TotalIncidenceRate)) = 0;
    h_DEF = heatmap(doseLegend, ageLegend, TotalIncidenceRate);
    sgtitle('Total Attack Rate by Age and Dose Group with Simulated R_0');
    title(['R_0 = ', num2str(R0(i))]);
    h_DEF.CellLabelFormat = '%.2e ';

    % how to define and compute the accumulativeNumberOfCases from model directly
    % instead of a extra numerical integration, it is still an very interesting question.
    Nt = sum(V + E + F + I + A + Q + R) / sum(N);
end
```

##### 6.3.3 Function: dxdt_VEFIAQR

```
function dxdt = dxdt_VEFIAQR(x,t,params)
%%% Consider the differencial equation: dx/dt = f(x,t,params),
%%% This function evaluates the derivative f(x,t) at a given point (x,t)
% INPUT:
%  x = [V0, V1, V2, V3,...
%       E0, E1, E2, E3,...
%       F0, F1, F2, F3,...
%       I0, I1, I2, I3,...
%       A0, A1, A2, A3,...
%       Q0, Q1, Q2, Q3,...
%       R0, R1, R2, R3];   a n*28 matrix

%
%
%    n   : number of age groups
%    t   : time variable
%  params: a struct of model parameters (assume that all parameters are group-distincted)
%
%       params.Beta   : n*n matrix, whose entries are beta_ij (beta_ij denotes the transimission ...
    capacity from group i to group j)
```

```
%       params.VE     : vaccine effecacy
%       params.kappa  : describe the difference of Transmission capacity between Iij and Aij
%       params.omega  : transition rate from Eij to Fij
%       params.omegap : (omegap is the short of omega_prime) transition rate from Fij to Iij
%       params.omegapp: transition rate from Fij to Aij
%       params.p      : proportion of asymptomatic cases
%       params.gamma  : the inverse of infectious period of Iij
%       params.gammap : (gammap is the short of gamma_prime) the inverse of average infectious ...
    period of A
%       params.gammapp: remove rate of Qij
%       params.mu     : rate of quarantine
%       params.f      : fatality rate of Iij
%       params.br     : birth rate of group ij (INTERSECTION of age group i and vaccinated group j)
%       params.dr     : natrual death rate of group ij
%
%  NOTE:
%      n*n matrices: Beta
%      n*4 matrices: kappa, omega, omegap, p, gamma, gammap, f, br, dr
%          vectors : Vi, Ei, Fi, Ii, Ai, Qi, Ri, i = 1, 2,..., n.
%          scalars : mu
%
%
% OUTPUT:
%       f = f(x,t,params), a vector of derivatives
%
%
% by Guo Xiaohao, 2021/09/23
%

tempID = 1:4;
V = x(:, tempID + 4*0);
E = x(:, tempID + 4*1);
F = x(:, tempID + 4*2);
I = x(:, tempID + 4*3);
A = x(:, tempID + 4*4);
Q = x(:, tempID + 4*5);
R = x(:, tempID + 4*6);

% Population Size in Each Age Group
N = sum(x,2);

```

```
Beta = params.Beta;
VE = params.VE;
kappa = params.kappa;
omega = params.omega;
omegap = params.omegap;
omegapp = params.omegapp;
p = params.p;
gamma = params.gamma;
gammap = params.gammap;
gammapp = params.gammapp;
mu = params.mu;
f = params.f;
br = params.br;
dr = params.dr;


% infectiveVector = (I + kappa .* (F + A)) * [0.5, 0.4, 0.3, 0.2].'; ()
infectiveVector = sum(I + kappa .* (F + A), 2); % \sum_k  (I_jk + kappa(E_jk + Ajk))
infectionForce = Beta.' * infectiveVector;     % lambda, a n*1 vector


dVdt = -infectionForce.*(1-VE).*V;
dEdt =  infectionForce.*(1-VE).*V - (1-p).*omega.*E - p.*omegap.*E;
dFdt = (1-p).*omega.*E - omegapp.*F;
dIdt = omegapp.*F - mu.*I - gamma.*I;
dAdt = p.*omegap.*E - mu.*A - gammap.*A;
dQdt = mu.*A + mu.*I - gammapp.*Q;
dRdt = gamma.*I + gammap.*A + gammapp.*Q;


dxdt = [dVdt, dEdt, dFdt, dIdt, dAdt, dQdt, dRdt];
end
```

##### 6.3.4 Function: extractVariableFromCells_forVEFIAQR

```
function [V, E, F, I, A, Q, R] = extractVariableFromCells_forVEFIAQR(x)
% the dimension of temp told everything
%
% by Guo Xiaohao, 2021/09/23
```

```
%

m = numel(x);
[n,¬] = size(x{1});
id = 1:4;

% save V
if nargout ≥ 1
    V = zeros(m,n*4);
    for i = 1:m
        temp = x{i}(:,id + 4 * 0);
        V(i,:) = temp(:);
    end
    varargout{1} = V;
end

% save E
if nargout ≥ 2
    E = zeros(m,n*4);
    for i = 1:m
        temp = x{i}(:,id + 4 * 1);
        E(i,:) = temp(:);
    end
    varargout{2} = E;
end

% save F
if nargout ≥ 3
    F = zeros(m,n*4);
    for i = 1:m
        temp = x{i}(:,id + 4 * 2);
        F(i,:) = temp(:);
    end
    varargout{3} = F;
end


% save I
if nargout ≥ 4
    I = zeros(m,n*4);
    for i = 1:m
```

```
        temp = x{i}(:,id + 4 * 3);
        I(i,:) = temp(:);
    end
    varargout{4} = I;
end


% save A
if nargout ≥ 5
    A = zeros(m,n*4);
    for i = 1:m
        temp = x{i}(:,id + 4 * 4);
        A(i,:) = temp(:);
    end
    varargout{5} = A;
end

% save Q
if nargout ≥ 6
    Q = zeros(m,n*4);
    for i = 1:m
        temp = x{i}(:,id + 4 * 5);
        Q(i,:) = temp(:);
    end
    varargout{6} = Q;
end

% save R
if nargout ≥ 7
    R = zeros(m,n*4);
    for i = 1:m
        temp = x{i}(:,id + 4 * 6);
        R(i,:) = temp(:);
    end
    varargout{7} = R;
end
```

##### 6.3.5 Function: extractByVaccineDoses

```
function I_by_doses = extractByVaccineDoses(I)


[m,¬] = size(I);

I_by_doses = zeros(m,4);

for i = 1:4
    I_by_doses(:,i) = sum(I(:,(1:8)+(i-1)*8), 2);
end

end
```

#### 6.4 NPI Simulations

```
function [] = contactMatrixDecomposition()

ImportAgePartition;
caseContactData = readtable('caseContactDataForDecomposition.xlsx');

relation = caseContactData.relation;
setting = caseContactData.setting;

uniqueSetting = unique(setting);
uniqueNames = {'Empty', 'Public Transports', 'Amusement', 'School', 'Household', 'Hospital',...
    'Working Area', 'Service Industry', 'Uncategorized', 'Community', 'Catering industry'};

N1 = zeros(numel(uniqueSetting),1);
for i = 1:numel(uniqueSetting)
    N1(i) = sum(strcmp(setting, uniqueSetting{i}) & ¬isnan(caseContactData.contactAge));
end
sumN1 = sum(N1(2:end));
sumN2 = numel(unique(caseContactData.caseName));

N2 = zeros(size(N1));
C_recovered = zeros(numel(agePartition)-1);
figureCount = 0;
for i = 1:numel(uniqueSetting)

```

```
    % isempty
    if isempty(uniqueSetting{i})
        continue;
    end

    % extract rows corresponding to uniqueSetting{i}
    indices = strcmp(setting, uniqueSetting{i});
    caseContactDatai = caseContactData(indices,:);
    N2(i) = numel(unique(caseContactDatai.caseName));

    % for caseContactData corresponding to uniqueSetting{i}, estimate the contact matrix via MLE
    [Ci,normalizedContactDataMatrix] = estimateC_i(caseContactDatai, agePartition);

    % replace nan by 0
    Ci(isnan(Ci)) = 0;
    normalizedContactDataMatrix(isnan(normalizedContactDataMatrix)) = 0;

    % composition of Ci
    % C_recovered = C_recovered + Ci;% * N2(i) / sumN2;
    % Note that there is no explicit relation between the whole C and the
    % decomposited Ci. This is because one participant may comtribute to
    % contacts amnong many senerios.
    % By simply assume that all participants are equally contribute to all
    % senerios, the contact matrix recovered frome Ci will be exaggerate.

    % heatmap of un-normalized contactDataMatrix
    figure;     figureCount = figureCount + 1;
    format shorte;
    xvalue = {'0 to 9','10 to 19','20 to 29', '30 to 39', '40 to 49', '50 to 59', '60 to 69', ...
        '\geq 70'};
    yvalue = {'0 to 9','10 to 19','20 to 29', '30 to 39', '40 to 49', '50 to 59', '60 to 69', ...
        '\geq 70'};
    h_DEF = heatmap(xvalue, yvalue, normalizedContactDataMatrix);
    h_DEF.Title = ['(', num2str(figureCount), ') ', uniqueNames{i}, ', Normalized Contact Data ...
        Matrix'];
    h_DEF.XLabel = 'age group i';
    h_DEF.YLabel = 'age group j';
    h_DEF.CellLabelFormat = '%.2f ';

    % heatmap of un-normalized contactMatrix
    figure;     figureCount = figureCount + 1;
```

```
    format shorte;
    xvalue = {'0 to 9','10 to 19','20 to 29', '30 to 39', '40 to 49', '50 to 59', '60 to 69', ...
        '\geq 70'};
    yvalue = {'0 to 9','10 to 19','20 to 29', '30 to 39', '40 to 49', '50 to 59', '60 to 69', ...
        '\geq 70'};
    h_DEF = heatmap(xvalue, yvalue, Ci);
    h_DEF.Title = ['(', num2str(figureCount), ') ', uniqueNames{i}, ', Contact Matrix Estimated ...
        via MLE'];
    h_DEF.XLabel = 'age group i';
    h_DEF.YLabel = 'age group j';
    h_DEF.CellLabelFormat = '%.2f ';

end

%     % heatmap of recovered contactDataMatrix
%     figure;
%     format shorte;
%     xvalue = {'0 to 9','10 to 19','20 to 29', '30 to 39', '40 to 49', '50 to 59', '60 to 69', ...
    '\geq 70'};
%     yvalue = {'0 to 9','10 to 19','20 to 29', '30 to 39', '40 to 49', '50 to 59', '60 to 69', ...
    '\geq 70'};
%     h_DEF = heatmap(xvalue, yvalue, C_recovered);
%     h_DEF.Title = 'Recovered Contact Matrix';
%     h_DEF.XLabel = 'age group i';
%     h_DEF.YLabel = 'age group j';
%     h_DEF.CellLabelFormat = '%.2f ';
end


function [C_MLE, varargout] = estimateC_i(data, agePartition)

data(:,[17,18]) = [];

% caseNames
caseNames = data.caseNameIncaseData;
[uniqueCaseNames,uniqueCaseIndex] = unique(caseNames);

emptyID = cellfun(@isempty,caseNames);
data(emptyID,:) = [];
%data(isnan(data.contactAge),:) = [];

```

```
caseNames = data.caseNameIncaseData;
[uniqueCaseNames,uniqueCaseIndex] = unique(caseNames);


% caseAge, contactAge
caseAge = data{:,3};

contactAge = data{:,15};
n = numel(caseAge);

%%% Construct A, the Contact Data Matrix
% contactDataMatrix(i,j) denotes the daily average number of close contact in group
% j, will produced by a individual in group i.

% Assumption 1:  the contact are uniformly distributed
% Assumption 2:  contactMatrix is symmetric
% Assumption 3:  contacts in data meets the case only once
% Assumption 4: all contacts of one particular case are recorded for a four-day period before ...
    the date fo been diagnosed

% age partition
[contactDataMatrix, caseRepeatCount] = computeContactDataMatrix(caseNames, caseAge, contactAge, ...
    agePartition);

% normalize the contactDataMatrix by caseRepeatCount
normalizedContactDataMatrix = contactDataMatrix ./ caseRepeatCount;


%%% read population Data
populationData = readtable('populationData.xlsx');
ageGroup = populationData.ageGroup;
population = populationData.all;

agePopulation = zeros(numel(agePartition)-1,1);
for i = 1:numel(agePartition)-1
    agePopulation(i) = sum(population(ageGroup≥agePartition(i) & ageGroup < ...
        agePartition(i+1)),'omitnan');
end

%% maximum likelihood estimation of contact matrix C
```

```
C_MLE = estimateContactMatrix_MLE(contactDataMatrix,caseRepeatCount, agePopulation, 0);

%exportgraphics(gca,'contactMatrixGroupedByAges_MLE.jpg','Resolution',300);

% output contactDataMatrix as well
if nargout ≥ 2
    varargout{1} = normalizedContactDataMatrix;
end

end
```

#### 6.5 Vaccination Optimization

##### 6.5.1 Script: Visualize the vaccine coverage

```
clear; close all; clc;

n = 8;
ImportFigureLegends;

caseData129 = readtable('caseData129.xlsx');
nVC = readmatrix('nVC.xlsx'); % contactCount in (ageGroups, dosesGroups)
nVCC = readmatrix('nVCC.xlsx'); % caseCount in (ageGroups, dosesGroups)
nC = readmatrix('nC.xlsx'); % caseCount in ageGroups
caseAge = readmatrix('caseAge.xlsx'); % caseAge
N = readmatrix('agePopulationVector.xlsx');

%% Figure 0: visualize VC, the vaccine Coverage rate
VC = nVC ./ sum(nVC,2) * 100;
VC = [VC; sum(nVC,1) / sum(nVC,"all") * 100];
VC=VC(:,[4,3,2]); % 3, 2, 1-doses

figure;
ba = bar(VC,'stacked','FaceColor','flat');

c = colormap("summer");
% c = [102,194,165;...
%     141, 160, 203;...
%     252,141,98] /256;
```

```
for i = 1:size(VC,2)
    ba(i).CData = c(60*i+60,:);
    %ba(i).CData = c(i,:);
end


set(gca,'XTickLabel',[ageLegend,'Total']);
%legend('3-doses only','2-doses only','1-dose only');
legend('booster vaccinated', 'fully vaccinated', 'vaccinated but un-fully')
ylabel('Coverage Rate');


%%% add number for each bins of VC
[¬, ¬, ytips1] = ba.YEndPoints; ytips1 = ytips1 + 2;
[¬, ¬, xtips1] = ba.XEndPoints; xtips1 = xtips1 - 0.3;

for i=1:size(VC,1)
    % if this bin is too short
    if sum(VC(i,:)) < 5
        labels1 = num2str(sum(VC(i,:),2), '%.1f %%');
        text(xtips1(i),ytips1(i),labels1,'VerticalAlignment','middle');
        continue;
    end

    % otherwise, there are places to label the stacked bins
    for j=1:size(VC,2)
        if VC(i,j)>0
            labels_stacked=num2str(sum(VC(i,1:j)),'%.1f %%');
            hText = text(i, sum(VC(i,1:j),2), labels_stacked);
            set(hText, 'VerticalAlignment','top', 'HorizontalAlignment', 'center','FontSize',10, ...
                'Color','black');
        end
    end

end

%% Figure 1: Total Attack Rate with different doses of vaccination
P = nVCC ./ nVC;
contactInDoseGroup = sum(nVC, 1);
caseInDoseGroup = sum(nVCC, 1);
TAR = caseInDoseGroup ./ contactInDoseGroup;
```

```
figure;
b2 = bar(TAR);
set(b2, 'facecolor',[1,1,1]);

xlabel('Condition of Vaccination');
ylabel('Total attack rate (TAR)');
set(gca,'XTickLabel',{'none','unfully vaccinated','fully vaccinated','boster vaccinated'});

for i = 1:4
    labels_stacked=[num2str(sum(nVCC(:,i))),' cases'];
    hText = text(i, TAR(i)+3e-7, labels_stacked);
    set(hText, 'VerticalAlignment','top', 'HorizontalAlignment', 'center','FontSize',10, ...
        'Color','black');
end


%% Figure 2: Distribution of Number of Close Contact of those 129-th cases
numberOfCloseContact = caseData129.NumberOfCloseContact;
numberOfCloseContact(isnan(numberOfCloseContact)) = [];

XLABEL = 'Number of Close Contact';
YLABEL = 'density (frequency / binWidth)';
TITLE = '';
binWidth = 20;

fitDist(numberOfCloseContact, binWidth, XLABEL, YLABEL, TITLE);



%% Figure 3: Age distribution of cases
caseAge(isnan(caseAge)) = [];

XLABEL = 'Age';
YLABEL = 'density (frequency / binWidth)';
TITLE = '';
binWidth = 5;

fitDist(caseAge, binWidth, XLABEL, YLABEL, TITLE);
```

```
%% Figure 4: time interval from illness onset to positively tested
D1 = caseData129.DateOfIllnessOnset;
D2 = caseData129.DateOfPositiveTest;
dD21 = days(D2 - D1);
dD21(isnan(dD21)) = [];
dD21(dD21 == 0) = [];

XLABEL = 'Time Interval from Illness Onset to Diagnosed (in days)';
YLABEL = 'density (frequency / binWidth)';
TITLE = '';
binWidth = 0.5;

fitDist(dD21, binWidth, XLABEL, YLABEL, TITLE);

%% Figure 5: time interval from possible date of infection to illness onset
D0 = caseData129.PossibleDateOfInfection;
idx = ¬isnan(D0);
D0 = D0(idx);
D0 = datetime(D0,'ConvertFrom','excel');

XLABEL = 'Possible Date of Infection to Illness Onset';
YLABEL = 'density (frequency / binWidth)';
TITLE = '';
binWidth = 1;
dD10 = days(D1(idx) - D0);
dD10(dD10==0) = [];

fitDist(dD10, binWidth, XLABEL, YLABEL, TITLE);

%% Figure 6: time interval from possible date of infection to illness onset
%% GT distribution
data = readtable('GTdata.xlsx');
gt = data.generation_time;
maxGT = max(gt);
minGT = min(gt);
len = maxGT-minGT;
freqGT = zeros(len,1);
for i = 1:len+1
```

```
    freqGT(i) = sum(gt == minGT + i - 1);
end
freqGT(freqGT==0) = [];

XLABEL = 'Generation Time (in days)';
YLABEL = 'frequency';
TITLE = '';
binWidth = 1;
fitDist(gt,binWidth, XLABEL, YLABEL, TITLE);
```

##### 6.5.2 Script: testVaccinateOptimization

```
clear; close all; clc;

format shortE
ImportParametersVEFIAQR;   % import parameters omega, gamma, p, kappa,...
ImportFigureLegends; % import the following cell array for figure legend:
% groupLegend; ageLegend;
N = readmatrix('agePopulationVector.xlsx'); % the population vector (stratified by age)
C = readmatrix('contactMatrix.xlsx');   % the contact matrix
nVC = readmatrix('nVC.xlsx'); %vaccineConvergenceViaData();



% time span
tInit = 0;
tFinal = 100;

% step size
stepSize = 1;

%% Determine all parameters in ODEs, probability vector q (DBM and NGM are adopted ...
    respectively), for infection by a one-time contact
R0 = [0.5, 1, 1.5, 2.5, 3, 3.5, 4, 7]';


q_DBM = zeros(numel(R0),1);
q_NGM = zeros(numel(R0),1);

```

```
for i = 1:n
    for j = 1:n
        R(i,j) = C(j,i) / (dr(i) + p(i)*omegap(i) + (1-p(i))*omega(i)) * ...
            (kappa(i)*p(i)*omegap(i)/(gammap(i)+dr(i)) + (1-p(i))*omega(i)/(gamma(i)+dr(i)+f(i)));
    end
end


for i = 1:numel(R0)
    temp1 = R .* (N.') ./ sum(N);
    temp2 = sum(temp1,1);
    q_DBM(i) = R0(i) / sum(temp2);

    temp3 = 1 / (dr(i) + p(i)*omegap(i) + (1-p(i))*omega(i)) * ...
        (kappa(i)*p(i)*omegap(i)/(gammap(i)+dr(i)) + (1-p(i))*omega(i)/(gamma(i)+dr(i)+f(i)));
    q_NGM(i) = R0(i) / (max(eig(C)) * temp3);
end
% fprintf('[R0, q_DBM, q_NGM] = \n');
% disp([R0, q_DBM, q_NGM]);


%% for each R0, plot the reduction of accumulative cases
%% for R0 = 4, plot the vaccinating process
for r = 7 %1:numel(R0)


    %% Initial Values for VEFIAQR model
    % initial point
    feasibleVC = nVC ./ sum(nVC,2);
    xInit = [feasibleVC .* N, zeros(n,24)];
    doseCount = sum(xInit(:,1:4), 1);
    xInit(6,16) = 1;        % first case in I_{6,3}

    % Parameters
    params.Beta   = C * q_NGM(r) ./ (N');




    %% stepwise vaccinating process
    % OD equations of this model
```

```
    fun = @(x,t)dxdt_VEFIAQR(x,t,params);

    % Init ObjFun (accumulative number of cases under init vaccine coverage)
    fInit0 = AccumulativeCases(fun, params, tInit, xInit, tFinal, stepSize);

    M = 1e8; % number of doses available
    m = 5e4; % stepSize for Vaccination
    record = zeros(floor(M/m), 4);
    stateRecord = zeros(floor(M/m),n,4);
    fRecord = zeros(floor(M/m)+1,1);
    fRecord(1) = fInit0;

    % stepwise vaccinating
    for k = 1 : floor(M/m)
        df = zeros(n,3); % decreasement under different vaccinating strategies
        fInit = zeros(n,3); % save the object function under the previous vaccine convergence
        m_k = zeros(n,3); % feasible stepsize for dose vaccination
        for i = 1:n     % number of age groups: n = 8
            for j = 1:3 % 0, 1, 2 doses
                [df(i,j), fInit(i,j), m_k(i,j)] = decisionEffectiveness(i, j, m, fun, params, ...
                    tInit, xInit, fInit0, tFinal, stepSize);
            end
        end

        % prepare for next iteration
        [temp, idx] = min(df(:));
        j = ceil(idx / 8);
        i = idx - 8*(j-1);
        dVC = feasibleDirection(i,j);
        xInit(:,1:4) = xInit(:,1:4) + dVC * m_k(idx);
        fInit0 = fInit(idx);

        disp(xInit(:,1:4))


        % save record
        fRecord(k+1) = fRecord(k) + temp * m_k(idx);
        record(k,:) = [i, j, df(idx), fInit(idx)];

        stateRecord(k,:,:) = xInit(:,1:4);
    end
```

```


    %% visualizing the stepwise vaccinating process
    if r == 7
        figure;
        for i = 1:n
            for j = 1:4
                subplot(2,ceil(n/2),i);
                plot(m*(1:floor(M/m)),stateRecord(:,i,j)); hold on;
            end
            legend(doseLegend);

            xlabel('Process (in doses)');
            ylabel(['Population Size']);
            sgtitle('Optimized Vaccinating Process in Each Age group')
            subtitle(['Age Group of ', ageLegend{i}]);

        end
    end

    figure(2);
    subplot(2,ceil(numel(R0) / 2), r);
    plot((0:floor(M/m)) * m, fRecord);
    sgtitle('Effectiveness of vaccination: Accumulative cases within 14-days');
    title(['R_0 = ', num2str(R0(r))]);
    xlabel('Processing (in doses)');
    ylabel('number of accumulative cases');

end
```

##### 6.5.3 Script: testMyBootstrap

```
clear; close all; clc;

maxIter = 1e4; % max iteration for bootstrapping
N = 1e4;    % size of bootstrap sample set
agePartition = [0,10,20,30,40,50,60,70,200];
n = 8;
ImportFigureLegends;
```

```
ImportAgePartition;
caseContactData = readtable('caseContactDataForDecomposition.xlsx');
[n,¬] = size(caseContactData);
[caseNames, uniqueIndex] = unique(caseContactData.caseNameIncaseData); caseNames(1) = [];
caseAge = caseContactData.caseAge(uniqueIndex(2:end));
contactNames = caseContactData.contactName;
contactAge = caseContactData.contactAge;
vaccines = caseContactData(:,30:end);

data = zeros(n,3);
forDiscrimination = zeros(n,3);
for i = 1:n

    % skip the missing contactAges
    if isnan(contactAge(i))
        continue;
    end

    % how many doses this contact has taken
    dosei = ¬isempty(vaccines.time1{i}) + ¬isempty(vaccines.time2{i}) + ¬isempty(vaccines.time3{i});

    % save whether the contact is a case, for further discrimination analysis
    isCase = sum(strcmp(contactNames(i), caseNames)) ≥ 1;
    forDiscrimination(i,:) = [isCase, contactAge(i), dosei];

    % if the contact is a case, save to nCC
    if isCase
        data(i,1:3) = [dosei, 1, contactAge(i)]; % [contactDoses, isCase, age]

    else
        data(i,1:3) = [dosei, 0, contactAge(i)];
    end
end

temp1 = forDiscrimination(forDiscrimination(:,1) == 1,:);
temp0 = forDiscrimination(forDiscrimination(:,1) == 0,:);
idx = randi(size(temp1,1), [size(temp0,1) - size(temp1,1), 1]);
forDiscrimination_sampleSizeBalanced = [temp0; temp1; temp1(idx,:)];
writematrix(forDiscrimination_sampleSizeBalanced,'forDiscrimination_sampleSizeBalanced.csv');

%% bootstrapping
```

```
pBootstraped = zeros(maxIter, 4);
tarRecord = zeros(maxIter, 8, 4);
for k = 1:maxIter % repeating bootstraping
    [bootstrapData] = myBootstrap(data,N);
    bootstrapVC = zeros(8,4);
    bootstrapCC = zeros(8,4);
    for i = 1:N

        group = whichGroup(bootstrapData(i,3), agePartition);

        for j = 0:3 % doses
            if bootstrapData(i,1) == j
                bootstrapVC(group, j+1) = bootstrapVC(group, j+1) + 1;
                if bootstrapData(i,2) == 1
                    bootstrapCC(group, j+1) = bootstrapCC(group, j+1) + 1;
                end
            end
        end

    end

    bootstrapVC;
    bootstrapCC;
    tarRecord(k, :, :) = bootstrapCC ./ bootstrapVC;
    tarRecord(isnan(tarRecord)) = 0;
    pBootstraped(k,:) = sum(bootstrapCC, 1) ./ sum(bootstrapVC, 1);
end

writematrix(pBootstraped,'pBootstraped.xlsx');


%%

% reform the tarRecord to a suitable format for trainning
tarForTrainning = zeros(numel(tarRecord), 3);
count = 1;
for k = 1:maxIter
    for i = 1:8
        for j = 1:4
            tarForTrainning(count, :) = [tarRecord(k,i,j), i, j];  % [value, groupi, dosej];
            count = count + 1;
```

```
            end
        end
    end

    writematrix(tarForTrainning,'tarForTrainning.csv')
    writematrix(squeeze(sum(tarRecord,2)),'TARdistributionInDoseGroups.xlsx');
    writematrix(squeeze(sum(tarRecord,3)),'TARdistributionInAgeGroups.xlsx');

    figure;
    b1 = boxplot(squeeze(sum(tarRecord,2)),'Notch','on');
    xticklabels(doseLegend);
    title('Total Attack Rate In People Finished Different Doses');
    ylabel('TAR')

    figure;
    b2 = boxplot(squeeze(sum(tarRecord,3)),'Notch','on');
    ax = gca;
    ax.TickLabelInterpreter = 'tex';
    xticklabels(ageLegend);
    title('Total Attack Rate In Different Age Groups');
    ylabel('TAR')

    % quantDistribution = [0 0.25 0.5 0.75 1];
    % b3 = boxPlot3D(tarRecord);
    % xticks(1:8);
    % xticklabels(ageLegend);
    % yticklabels(doseLegend);

    figure;
    scatter3(tarForTrainning(:,2),tarForTrainning(:,3),tarForTrainning(:,1),'bo');
    xticks(1:8);
    xticklabels(ageLegend);
    yticklabels(doseLegend);
```

##### 6.5.4 Script: ImportFigureLegends

```
%%% groupLegend
groupLegend = cell(n,1);
for i = 1:n
```

```
    groupLegend{i,1} = ['group ',num2str(i)];
end

%%% ageLegends
ageLegend = {'0 to 9','10 to 19','20 to 29', '30 to 39',...
             '40 to 49', '50 to 59', '60 to 69', '\geq 70' };



%%% doseLegends
%doseLegend = {'none', '1 dose', '2 doses', '3 doses'};
doseLegend = {'none', 'unfully vaccinated', 'fully vaccinated', 'booster vaccinated'};
```

##### 6.5.5 Function: vaccineCoverageViaDataByIsComplete

```
function [nVC,nVCC] = vaccineCoverageViaDataByIsComplete()

ImportAgePartition;
caseContactData = ...
    readtable('caseContactDataForDecompositionWithGenderAndVaccineDoseCorrected.xlsx');
caseNames = unique(caseContactData.caseNameIncaseData);
caseNames(1) = []; % delete the empty
contactNames = caseContactData.contactName;
contactAge = caseContactData.contactAge;
vaccines = caseContactData(:,30:end);

nVC = zeros(numel(agePartition)-1, 4);
nVCC = zeros(numel(agePartition)-1, 4);
for i = 1:size(caseContactData,1)

    if i == 7882
        1;
    end

    % skip the missing contactAges
    if isnan(contactAge(i))
        continue;
    end

```

```
    % how many doses this contact has taken
    doseNum = ¬isempty(vaccines.time1{i}) + ¬isempty(vaccines.time2{i}) + ¬...
        isempty(vaccines.time3{i});
    doseDemand = max([vaccines.doseDemand1(i), vaccines.doseDemand2(i), vaccines.doseDemand3(i)]);

    if doseNum == 0
        dosei = 0;     % none
    elseif doseNum < doseDemand
        dosei = 1;     % unfully vaccinated
    elseif doseNum == doseDemand
        dosei = 2;     % fully vaccinated
    elseif doseNum > doseDemand
        dosei = 3;     % booster vaccinated
    end

    % which group this contact belongs to
    groupi = whichGroup(contactAge(i), agePartition);

    % add this contact to corresponding age-stratified coverage table
    nVC(groupi, dosei + 1) = nVC(groupi, dosei + 1) + 1;

    % if the contact is a case, save to nVCC
    if sum(strcmp(contactNames(i), caseNames)) ≥ 1
        nVCC(groupi, dosei + 1) = nVCC(groupi, dosei + 1) + 1 / ...
            sum(strcmp(contactNames,contactNames(i)));
    end
end

writematrix(nVC,'nVC.xlsx');
writematrix(nVCC,'nVCC.xlsx');
1;
end
```

##### 6.5.6 Function: myBootstrap

```
function [bootstrapData] = myBootstrap(data,N)

[m,¬] = size(data);
idx = randi(m,[N,1]);
```

```
bootstrapData = data(idx,:);
end
```

##### 6.5.7 Function: fitDist

```
function [] = fitDist(data, binWidth, XLABEL, YLABEL, TITLE)
pd1_Gamma = fitdist(data,'Gamma')
pd1_Weibull = fitdist(data,'Weibull')
pd1_Lognormal = fitdist(data,'Lognormal')
a = pd1_Gamma.a;
b = pd1_Gamma.b;
A = pd1_Weibull.A;
B = pd1_Weibull.B;
mu = pd1_Lognormal.mu;
sigma = pd1_Lognormal.sigma;

x = linspace(0, max(data), 1e4);
y1_Gamma = pdf(pd1_Gamma,x);
y1_Weibull = pdf(pd1_Weibull,x);
y1_Lognormal = pdf(pd1_Lognormal,x);

figure;
plot(x,y1_Gamma,'LineWidth',2); hold on;
plot(x,y1_Weibull,'LineWidth',2); hold on;
plot(x,y1_Lognormal,'LineWidth',2); hold on;
h1 = histogram(data,'Normalization','pdf','FaceColor',[0 0.4470 0.7410],'binWidth',binWidth);

LEGEND = {['Gamma,     scale = ', num2str(b),'  shape = ', num2str(a)],...
    ['Weibull,     scale = ', num2str(A), '  shape = ',num2str(B)],...
    ['Lognormal, log-Scale = ', num2str(mu), '  log-Location = ', num2str(sigma)]};

legend(LEGEND);
xlabel(XLABEL);
ylabel(YLABEL);
title(TITLE);

end
```

##### 6.5.8 Function: feasibleDirection

```
function dVC = feasibleDirection(i,j)

dVC = zeros(8,4);
dVC(i,j) = -1;
dVC(i,j+1) = 1;
end
```

##### 6.5.9 Function: decisionEffectiveness

```
function [df, fInit, dm] = decisionEffectiveness(i, j, dm, fun, params, tInit, xInit, fInit, ...
    tFinal, stepSize)
%%% evaluate the decision effectiveness of assign this dose to the ij-th
%%% group (those finished (j-1) doses in age group i)
%
%%% INPUT:
%   i,j          : the ij-th group
%   dm           : number of doses for one-time distributing
%   fun          : objective loss funtion
%   params       : model parameters
%   tInit        : time instance for simulation start
%   xInit        : current vaccine covorage at tInit
%   fInit        : the value of the objective function at current xInit
%   tFinal       : time instance for simulation end
%   stepSize     : stepSize for RK-4 solver
%
%%% OUTPUT:
%   df           : change of the objective function (the effectness of this dose)
%   fInit        : value of the objective function after update
%   dm           : output dm unchanged


% if population size of group ij equals to 0
if xInit(i,j) ≤ 0
    df = 0;
    dm = 0;
    return;
```

```
end

% if population size is greater 0 but lesser than xInit(i,j)
if xInit(i,j) - dm < 0
    dm = xInit(i,j);
end

xInit2 = xInit;
xInit2(:,1:4) = xInit2(:,1:4) + dm * feasibleDirection(i,j);
f1 = fInit; %f1 = AccumulativeCases(fun, params, tInit, xInit1, tFinal, stepSize);
f2 = AccumulativeCases(fun, params, tInit, xInit2, tFinal, stepSize);

df = (f2 - f1) / dm; % accumulative cases prevention of per doses
fInit = f2;

end
```

##### 6.5.10 Function: caseCountViaData

```
function nC = caseCountViaData()
ImportAgePartition;
caseContactData = readtable('caseContactDataForDecomposition.xlsx');
[uniqueCases, uniqueID] = unique(caseContactData.caseNameIncaseData);

caseAge = caseContactData.caseAge(uniqueID);
vaccines = caseContactData(uniqueID,30:end);

nC = zeros(numel(agePartition)-1, 1);
for i = 1:size(uniqueCases,1)

    % skip the missing contactAges
    if isnan(caseAge(i))
        continue;
    end

%     % how many doses this contact has taken
%     dosei = ¬isempty(vaccines.time1{i}) + ¬isempty(vaccines.time2{i}) + ¬...
    isempty(vaccines.time3{i});

```

```
    % which group this contact belongs to
    groupi = whichGroup(caseAge(i), agePartition);

    % add this contact to corresponding age-stratified convergence table
    nC(groupi, 1) = nC(groupi, 1) + 1;
end

writematrix(nC,'nC.xlsx');
end
```

##### 6.5.11 Function: AccumulativeCases

```
function accumulativeNumberOfCases = AccumulativeCases(fun, params, tInit, xInit, tFinal, stepSize)
%%% Compute all accumulative number of cases
%%% INPUT:
%   fun         : the derivative of the systems of ODEs
%   params      : parameters in ODEs, a struct
%   tInit       : time instance for simulation start
%   xInit       : initial value for the systems of ODEs
%   tFinal      : time instance for simulation end
%   stepSize    : step size for ode solvers
%
%%% OUTPUT:
% accumulativeNumberOfCases

% solve
[x,t] = odeSolveRK4(fun, tInit, xInit, tFinal, stepSize);
[V, E, F, I, A, Q, R] = extractVariableFromCells_forVEFIAQR(x);

% daily New Cases
p = params.p;
omegap = params.omegap;
omega = params.omega;
dailyNewCases = extractByVaccineDoses(p(:)'.*omegap(:)'.*E + (1-p(:)').*omega(:)'.*E);

% accumulative via integral
accumulativeNumberOfCases = zeros(4,1);
for k = 1:4
    accumulativeNumberOfCases(k) = trapz(t, dailyNewCases(:,k));
```

```
end

% output
accumulativeNumberOfCases = sum(accumulativeNumberOfCases);

end
```

### References

[1] ... Hongjie Yu. Time-varying optimization of covid-19 vaccine prioritization in the context of limited vaccination capacity. 2021.
